# Clinical Recognition of Frontotemporal Dementia with Right Temporal Predominance; Consensus Recommendations of the International Working Group

**DOI:** 10.1101/2024.10.18.24315786

**Authors:** Hulya Ulugut, Kyan Younes, Maxime Montembeault, Maxime Bertoux, Muireann Irish, Fiona Kumfor, Giorgio G. Fumagalli, Bedia Samanci, Ignacio Illán-Gala, Jennifer C. Thompson, Alexander F Santillo, Elisabet Englund, Maria Landqvist Waldö, Lina Riedl, Jan Van den Stock, Mathieu Vandenbulcke, Rik Vandenberghe, Robert Jr Laforce, Simon Ducharme, Peter S. Pressman, Paulo Caramelli, Leonardo Cruz de Souza, Leonel T. Takada, Hakan Gurvit, Janine Diehl-Schmid, Daniela Galimberti, Florence Pasquier, Sandra Weintraub, Bruce L. Miller, Virginia E. Sturm, Jennifer L. Whitwell, Bradley Boeve, Jonathan D. Rohrer, Olivier Piguet, Maria Luisa Gorno-Tempini, Keith A. Josephs, Julie Snowden, James B. Rowe, Jason D. Warren, Katherine P. Rankin, Yolande A.L. Pijnenburg, International rtvFTD working group

## Abstract

Accurate diagnosis of frontotemporal dementia (FTD) with right anterior temporal lobe (RATL) predominance remains challenging due to lack of clinical characterization, and standardized terminology. The recent research of the International Working Group (IWG) identified common symptoms but also unveiled broad terminologies lacking precision and operationalization, with risk of misdiagnoses, inappropriate referrals and poor clinical management. Based on the published evidence (91267 articles screened) and expert opinion (105 FTD specialists across 52 centers), the IWG delineates three primary domains of impairment causing behavioral, memory and language problems: (i) multimodal knowledge of non-verbal information including people, living beings, landmarks, flavors/odors, sounds, bodily sensations, emotions and social cues; (ii) socioemotional behavior encompassing emotion expression, social response and motivation; and (iii) prioritization for focus on specific interests, hedonic valuation and personal preferences. This study establishes a consensus on clinical profile, phenotypic nomenclature, and future directions to enhance diagnostic precision and therapeutic interventions.

## INTRODUCTION

Frontotemporal dementia (FTD) is one of the leading causes of dementia before age 65, and selectively impacts circuits in the frontal and anterior temporal lobes (ATL) that support behavior and language^1–3^. Despite the extensive study of semantic aphasia (left temporal dominant) and behavioral variant (frontal dominant) FTD syndromes, the syndrome of FTD related to pathology predominant in the right ATL (RATL) lacks standardized nomenclature and consensus diagnostic criteria^4^. The problem goes beyond the lack of a categorical term for the syndrome, with confusion or inconsistency in the very phenomenological characterization. Diverse and sometimes conflicting descriptions have been applied to the symptoms of FTD with RATL predominance. Hyper-religiosity, once regarded as virtually pathognomonic for this subtype, has been widely observed in numerous case reports, with influences from the “Geschwind Syndrome,” a concept derived from epilepsy literature that includes hyper-religiosity, hypergraphia, hyposexuality, and irritability^5^. Early publications with larger sample sizes highlighted prosopagnosia, parsimony, preoccupations, lack of empathy and compulsive behavior, while some emphasized loss of object knowledge and visual semantic deficits as core features^3,6–9^. Other studies pointed to memory deficits, mental rigidity, somatization, topographagnosia, atypical depression, slowness, hallucinations, and delusions^10–12^. Furthermore, a wide spectrum of singular case studies reported dysprosody, altered emotional expression, parosmia, gustatory agnosia, phonagnosia, musicophilia or amusia, visceral agnosia, and an array of emergent obsessions ranging from strong political or religious beliefs to artistic skills^13–21^. Most recently, some groups have proposed that socioemotional semantic deficits and altered hedonic valuation are the primary mechanisms underpinning the behavioral changes commonly seen in the RATL syndrome ^22–24^.

We established an international working group (IWG) in 2020 to elucidate the clinical characteristics of the syndrome, resolve contradictions in the field and promote consensus on terminology, for FTD presenting with RATL predominance. Our initial multi-cultural publication, which included 360 patients, showed that common symptoms included mental rigidity/ preoccupations (78%), disinhibition/socially inappropriate behavior (74%), naming/word-finding difficulties (70%), memory deficits (67%), apathy (65%), loss of empathy (65%), and face-recognition deficits (60%), as well as impairments regarding landmarks, smells, sounds, tastes, and bodily sensations (74%). Some of these were not specifically inquired about in many centers and others were interpreted heterogeneously using diverse terminologies^4^. Many symptoms in this study were described using broad terminologies such as ’disinhibition’ and ’word-finding difficulties’, which either lacked or misattributed underlying mechanisms. These descriptions were typically based on clinicians’ observations or informant-based surveys. The limited use of objective, face-to-face assessments for common and specific RATL symptoms led to heterogeneous interpretations. While available test results indicated deficits in the semantics of emotion, people, social interactions, and visual stimuli, we lacked objective assessments and operationalization for mental rigidity and preoccupations, despite the high prevalence^4^.

Lastly, our most recent multicultural cohort revealed that 80% of patients have no genetic variant or family history. Given the sporadic nature of the disease, early and accurate diagnosis is particularly challenging worldwide, often leading to undiagnosed or misdiagnosed cases as psychiatric disorders. Therefore, early and precise diagnoses, which are essential for improving patient care, advancing research, and facilitating clinical trial inclusion, primarily rely on thorough clinical assessments.

These results underscored the urgent need for consensus on terminologies that elucidate primary cortical dysfunctions and can be readily applied in everyday clinical practice globally. This is the first international initiation to address existing gaps and offer consensus recommendations by conducting a thorough systematic review and gathering expert opinions.

## METHODS

A comprehensive review in MEDLINE (PubMed) and Embase until February 2024 was performed. Thirteen separate systematic reviews were conducted to tackle each RATL relevant clinical symptom identified in our previous study and literature. Included symptoms were ‘person specific knowledge deficit’, ‘lack of empathy’, ‘disinhibition’, ‘apathy’, ‘mental rigidity’, ‘memory deficits’, ‘psychiatric symptoms’ and deficits regarding ‘smell’, ‘taste’, ‘sound’, ‘bodily sensations’, ‘visual’, ‘landmarks’. Search terms for each symptom were discussed and identified by the IWG consensus to increase the sensitivity of the search. (i.e., for person specific knowledge, search terms; “prosopagnosia” OR “associative prosopagnosia” OR “face recognition deficit” OR “person knowledge” OR “semantics for people” OR “face agnosia” OR “face blindness” OR “person identification” OR “person-specific” OR “person recognition” OR “face recognition”, [see the supplementary materials for the search-terms of other symptoms]). Subsequently each symptom was attached with the following terms “frontotemporal dementia” OR “semantic dementia” OR “semantic variant primary progressive aphasia” OR “behavioral variant frontotemporal dementia” OR “temporal variant frontotemporal dementia” OR “frontotemporal lobar degeneration” OR “Pick’s Disease” OR “right temporal” OR “right anterior*” OR “temporal pole”, “frontotemporal dementia AND right temporal”, “semantic dementia”, “semantic variant primary progressive aphasia AND right”, “behavioral variant frontotemporal dementia AND right”, “temporal variant frontotemporal dementia”, and “frontotemporal lobar degeneration AND right temporal” (see the supplementary materials for overview of the full electronic search strategy). Studies were excluded when no original data were reported (letters to the editor, meta-analyses, or review studies). Mendeley reference manager (v2.114.0) was used to register all citations. Duplicated studies were removed based on overlapping authorship, study description, year of publication, and journal. The titles and abstracts of the citations were then screened by 3 independent authors (KY, MM and HU) to determine their relevance for inclusion. Unlike other published systematic reviews on this topic, even if the prevalent RATL atrophy was not mentioned in the titles or abstracts, if there was a bvFTD, svPPA or SD cohort, full texts were screened to identify potential cases with predominant RATL atrophy (see the supplementary materials for case identification). Initial discordance between the three primary assessors were resolved through consensus or through the decision of a fourth author (YP). Consequently, all included articles were shared with the IWG for their review, and additional articles were included by the IWG if they were not in the initial search.

Full-text articles of the relevant citations were then assessed to determine whether the study met the predefined inclusion criteria. The study quality of all included articles was assessed using the IWG’s guidelines including a 6-point checklist assesses the rigor of inclusion criteria and subject selection, sample size, diversity (ethnicity/ country), biomarker, genetic and pathological confirmation availability, and measurement of symptoms (Table 1). Following this step, the IWG experts were requested to review the results before the meeting to provide their opinions on the symptoms during the meeting which was recorded to be further coded to capture the interpretations shared by the experts. In cases where members were unable to participate in the discussion or had additional comments, they were encouraged to watch the recorded videos and share their opinions via email. Subsequently, to determine the most appropriate terminology, a survey was conducted wherein each proposed term for each symptom was rated by the experts. Based on the survey results, a theoretical framework was developed for symptom categorization, reflecting the domains of impairment. These results were then shared with the IWG for further discussion. Experts deliberated on the terminology to be used, identified core features, and provided recommendations for the IWG’s guidance (see supplementary material for the establishment of the IWG, consensus approach, round table meetings).

## TERMINOLOGY

Table 2 presents real-life examples from the IWG’s dataset and illustrates how these examples are interpreted by caregivers in terms that belie the underlying cognitive deficit. For instance, knowledge loss for people may be perceived as a memory (e.g., “She doesn’t remember people…”), language (e.g., “He cannot name actors on TV…”), or a behavioral issue (e.g., “She behaves as if she is talking to strangers on the phone when she is actually talking to close friends and family”) (see examples for each symptom in Table 2). In the following section, the IWG meticulously reviewed each identified symptom from our previous work and the existing literature. The group conceptualized these symptoms by considering underlying neural mechanisms and provided recommended terminology and clinical guidelines to better capture these symptoms in daily clinical practice (Table 2, Box 1).

**Table 2.**
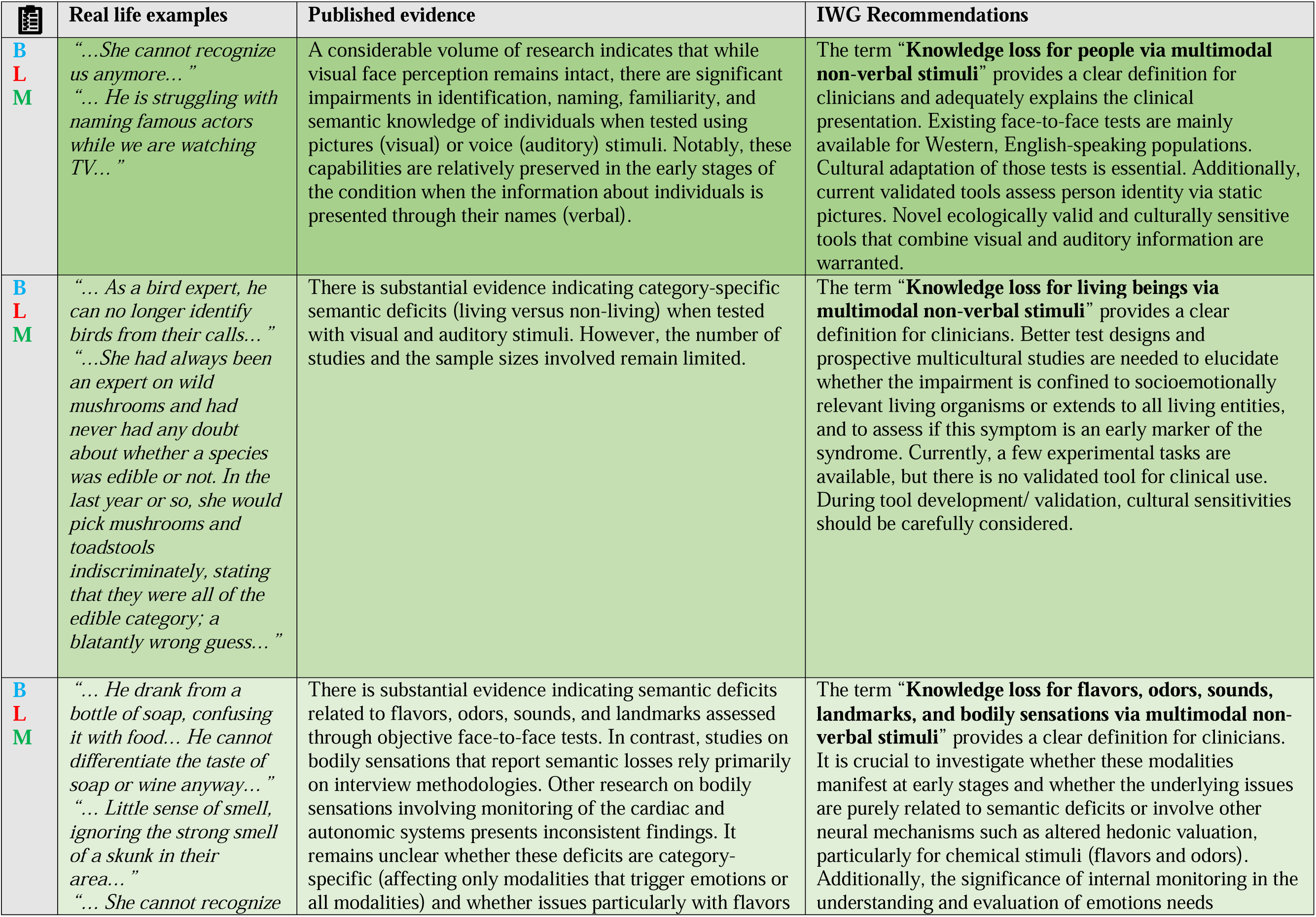
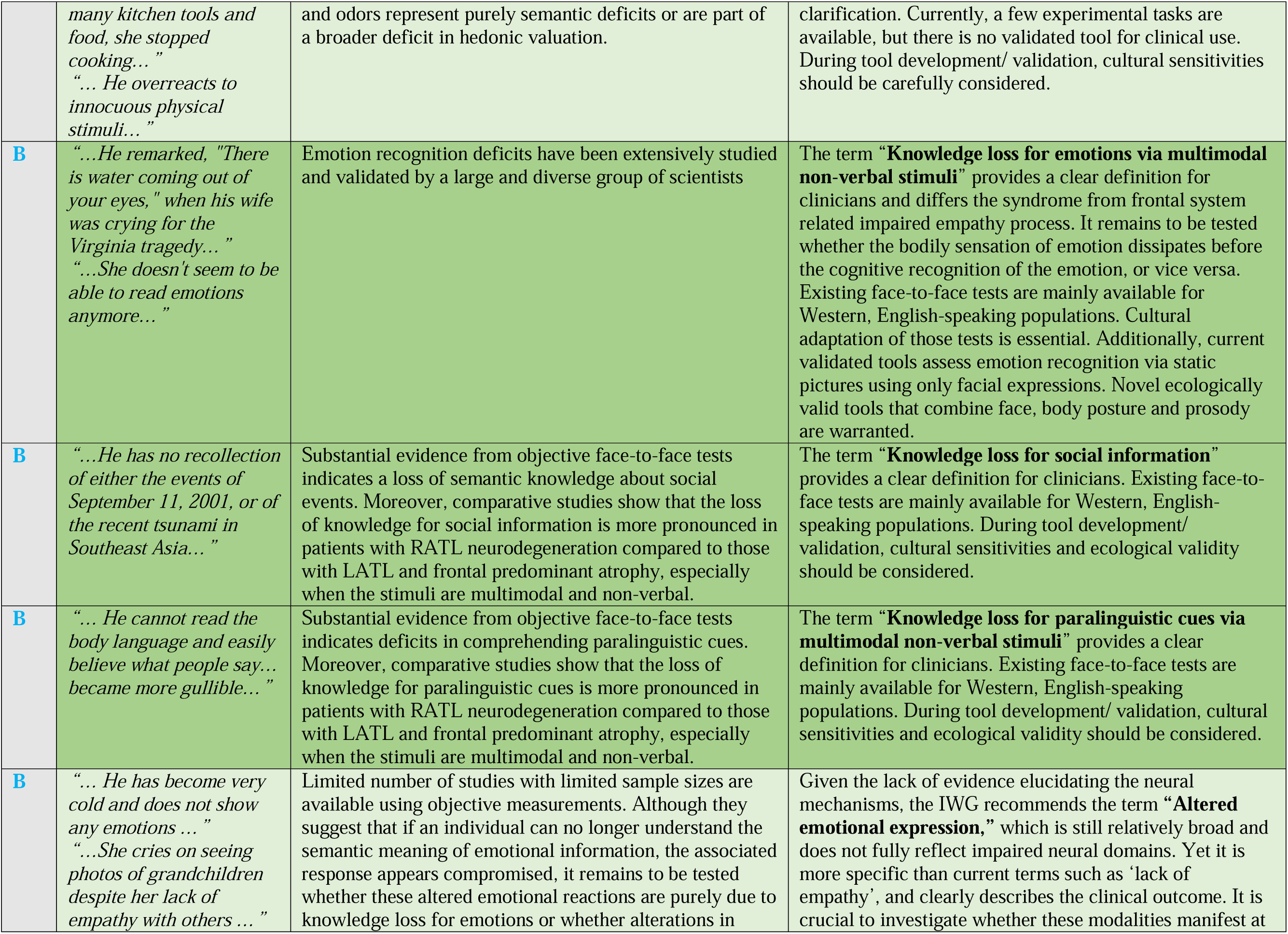
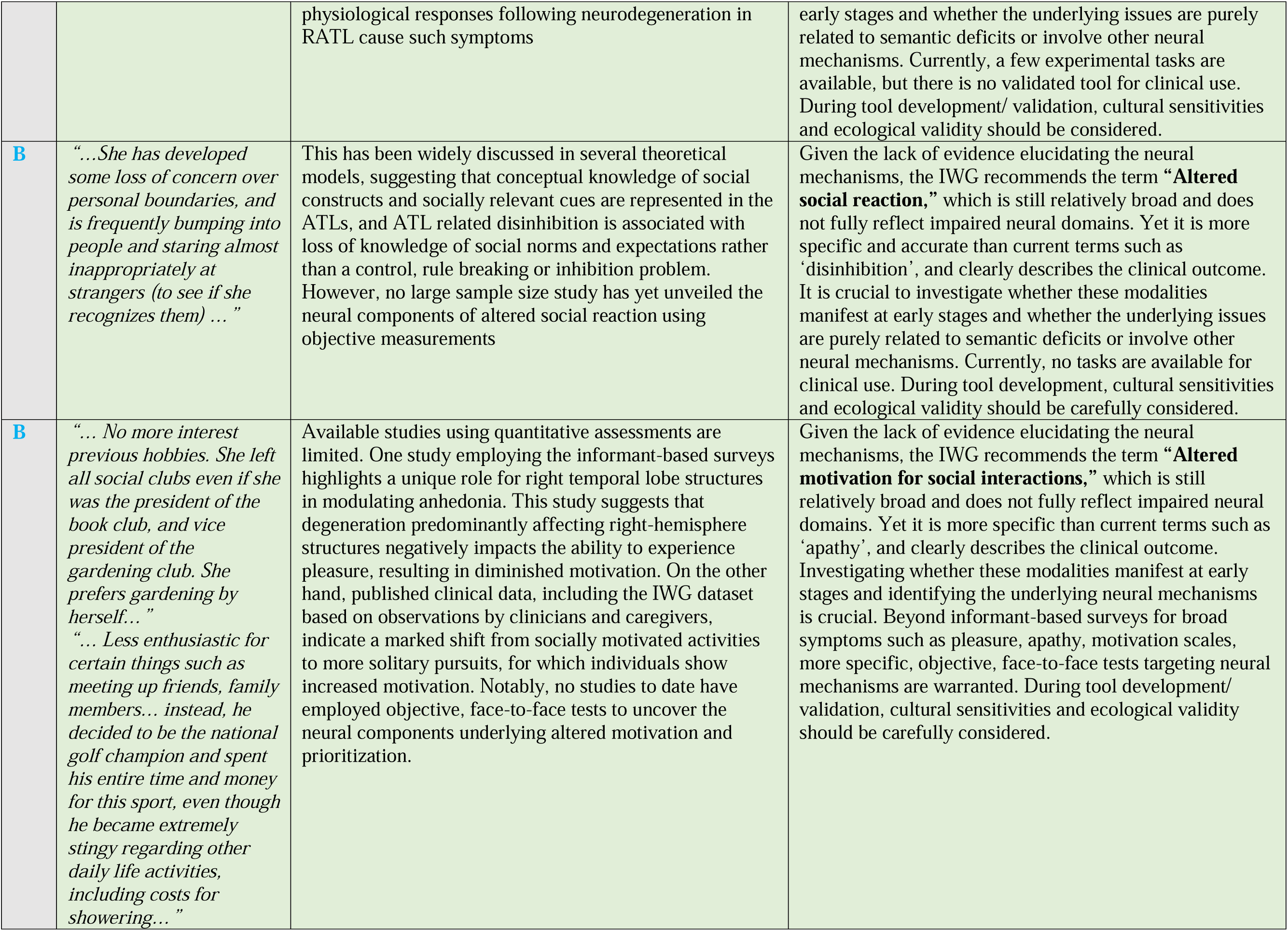
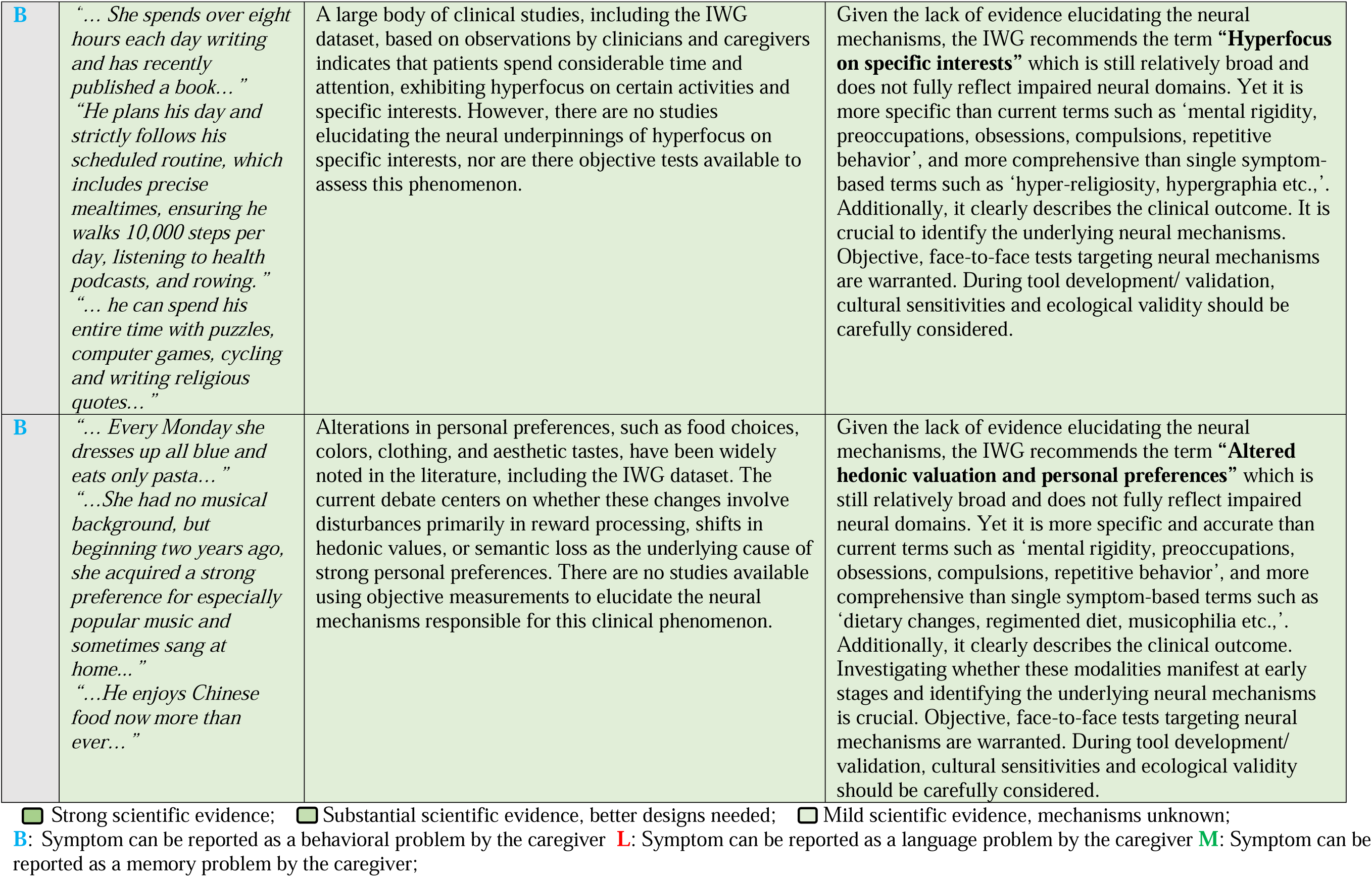
Interpretation of the reported symptoms and consensus recommendations.

|  | Real life examples | Published evidence | IWG Recommendations |
| --- | --- | --- | --- |
| B<br>L<br>M | *“...She cannot recognize us anymore...”*<br>*“... He is struggling with naming famous actors while we are watching TV...”* | A considerable volume of research indicates that while visual face perception remains intact, there are significant impairments in identification, naming, familiarity, and semantic knowledge of individuals when tested using pictures (visual) or voice (auditory) stimuli. Notably, these capabilities are relatively preserved in the early stages of the condition when the information about individuals is presented through their names (verbal). | The term “**Knowledge loss for people via multimodal non-verbal stimuli**” provides a clear definition for clinicians and adequately explains the clinical presentation. Existing face-to-face tests are mainly available for Western, English-speaking populations. Cultural adaptation of those tests is essential. Additionally, current validated tools assess person identity via static pictures. Novel ecologically valid and culturally sensitive tools that combine visual and auditory information are warranted. |
| B<br>L<br>M | *“... As a bird expert, he can no longer identify birds from their calls...”*<br>*“...She had always been an expert on wild mushrooms and had never had any doubt about whether a species was edible or not. In the last year or so, she would pick mushrooms and toadstools indiscriminately, stating that they were all of the edible category; a blatantly wrong guess...”* | There is substantial evidence indicating category-specific semantic deficits (living versus non-living) when tested with visual and auditory stimuli. However, the number of studies and the sample sizes involved remain limited. | The term “**Knowledge loss for living beings via multimodal non-verbal stimuli**” provides a clear definition for clinicians. Better test designs and prospective multicultural studies are needed to elucidate whether the impairment is confined to socioemotionally relevant living organisms or extends to all living entities, and to assess if this symptom is an early marker of the syndrome. Currently, a few experimental tasks are available, but there is no validated tool for clinical use. During tool development/ validation, cultural sensitivities should be carefully considered. |
| B<br>L<br>M | *“... He drank from a bottle of soap, confusing it with food... He cannot differentiate the taste of soap or wine anyway...”*<br>*“... Little sense of smell, ignoring the strong smell of a skunk in their area...”*<br>*“... She cannot recognize* | There is substantial evidence indicating semantic deficits related to flavors, odors, sounds, and landmarks assessed through objective face-to-face tests. In contrast, studies on bodily sensations that report semantic losses rely primarily on interview methodologies. Other research on bodily sensations involving monitoring of the cardiac and autonomic systems presents inconsistent findings. It remains unclear whether these deficits are category-specific (affecting only modalities that trigger emotions or all modalities) and whether issues particularly with flavors | The term “**Knowledge loss for flavors, odors, sounds, landmarks, and bodily sensations via multimodal non-verbal stimuli**” provides a clear definition for clinicians. It is crucial to investigate whether these modalities manifest at early stages and whether the underlying issues are purely related to semantic deficits or involve other neural mechanisms such as altered hedonic valuation, particularly for chemical stimuli (flavors and odors). Additionally, the significance of internal monitoring in the understanding and evaluation of emotions needs |
|  | *many kitchen tools and food, she stopped cooking...”*<br>*“... He overreacts to innocuous physical stimuli...”* | and odors represent purely semantic deficits or are part of a broader deficit in hedonic valuation. | clarification. Currently, a few experimental tasks are available, but there is no validated tool for clinical use. During tool development/ validation, cultural sensitivities should be carefully considered. |
| **B** | *“...He remarked, "There is water coming out of your eyes," when his wife was crying for the Virginia tragedy...”*<br>*“...She doesn't seem to be able to read emotions anymore...”* | Emotion recognition deficits have been extensively studied and validated by a large and diverse group of scientists | The term “**Knowledge loss for emotions via multimodal non-verbal stimuli**” provides a clear definition for clinicians and differs the syndrome from frontal system related impaired empathy process. It remains to be tested whether the bodily sensation of emotion dissipates before the cognitive recognition of the emotion, or vice versa. Existing face-to-face tests are mainly available for Western, English-speaking populations. Cultural adaptation of those tests is essential. Additionally, current validated tools assess emotion recognition via static pictures using only facial expressions. Novel ecologically valid tools that combine face, body posture and prosody are warranted. |
| **B** | *“...He has no recollection of either the events of September 11, 2001, or of the recent tsunami in Southeast Asia...”* | Substantial evidence from objective face-to-face tests indicates a loss of semantic knowledge about social events. Moreover, comparative studies show that the loss of knowledge for social information is more pronounced in patients with RATL neurodegeneration compared to those with LATL and frontal predominant atrophy, especially when the stimuli are multimodal and non-verbal. | The term “**Knowledge loss for social information**” provides a clear definition for clinicians. Existing face-to-face tests are mainly available for Western, English-speaking populations. During tool development/ validation, cultural sensitivities and ecological validity should be considered. |
| **B** | *“... He cannot read the body language and easily believe what people say... became more gullible...”* | Substantial evidence from objective face-to-face tests indicates deficits in comprehending paralinguistic cues. Moreover, comparative studies show that the loss of knowledge for paralinguistic cues is more pronounced in patients with RATL neurodegeneration compared to those with LATL and frontal predominant atrophy, especially when the stimuli are multimodal and non-verbal. | The term “**Knowledge loss for paralinguistic cues via multimodal non-verbal stimuli**” provides a clear definition for clinicians. Existing face-to-face tests are mainly available for Western, English-speaking populations. During tool development/ validation, cultural sensitivities and ecological validity should be considered. |
| **B** | *“... He has become very cold and does not show any emotions ...”*<br>*“...She cries on seeing photos of grandchildren despite her lack of empathy with others ...”* | Limited number of studies with limited sample sizes are available using objective measurements. Although they suggest that if an individual can no longer understand the semantic meaning of emotional information, the associated response appears compromised, it remains to be tested whether these altered emotional reactions are purely due to knowledge loss for emotions or whether alterations in | Given the lack of evidence elucidating the neural mechanisms, the IWG recommends the term “**Altered emotional expression,**” which is still relatively broad and does not fully reflect impaired neural domains. Yet it is more specific than current terms such as ‘lack of empathy’, and clearly describes the clinical outcome. It is crucial to investigate whether these modalities manifest at |
|  |  | physiological responses following neurodegeneration in RATL cause such symptoms | early stages and whether the underlying issues are purely related to semantic deficits or involve other neural mechanisms. Currently, a few experimental tasks are available, but there is no validated tool for clinical use. During tool development/ validation, cultural sensitivities and ecological validity should be considered. |
| **B** | *“...She has developed some loss of concern over personal boundaries, and is frequently bumping into people and staring almost inappropriately at strangers (to see if she recognizes them) ...”* | This has been widely discussed in several theoretical models, suggesting that conceptual knowledge of social constructs and socially relevant cues are represented in the ATLs, and ATL related disinhibition is associated with loss of knowledge of social norms and expectations rather than a control, rule breaking or inhibition problem. However, no large sample size study has yet unveiled the neural components of altered social reaction using objective measurements | Given the lack of evidence elucidating the neural mechanisms, the IWG recommends the term “**Altered social reaction,**” which is still relatively broad and does not fully reflect impaired neural domains. Yet it is more specific and accurate than current terms such as ‘disinhibition’, and clearly describes the clinical outcome. It is crucial to investigate whether these modalities manifest at early stages and whether the underlying issues are purely related to semantic deficits or involve other neural mechanisms. Currently, no tasks are available for clinical use. During tool development, cultural sensitivities and ecological validity should be carefully considered. |
| **B** | *“... No more interest previous hobbies. She left all social clubs even if she was the president of the book club, and vice president of the gardening club. She prefers gardening by herself...”*<br>*“... Less enthusiastic for certain things such as meeting up friends, family members... instead, he decided to be the national golf champion and spent his entire time and money for this sport, even though he became extremely stingy regarding other daily life activities, including costs for showering...”* | Available studies using quantitative assessments are limited. One study employing the informant-based surveys highlights a unique role for right temporal lobe structures in modulating anhedonia. This study suggests that degeneration predominantly affecting right-hemisphere structures negatively impacts the ability to experience pleasure, resulting in diminished motivation. On the other hand, published clinical data, including the IWG dataset based on observations by clinicians and caregivers, indicate a marked shift from socially motivated activities to more solitary pursuits, for which individuals show increased motivation. Notably, no studies to date have employed objective, face-to-face tests to uncover the neural components underlying altered motivation and prioritization. | Given the lack of evidence elucidating the neural mechanisms, the IWG recommends the term “**Altered motivation for social interactions,**” which is still relatively broad and does not fully reflect impaired neural domains. Yet it is more specific than current terms such as ‘apathy’, and clearly describes the clinical outcome. Investigating whether these modalities manifest at early stages and identifying the underlying neural mechanisms is crucial. Beyond informant-based surveys for broad symptoms such as pleasure, apathy, motivation scales, more specific, objective, face-to-face tests targeting neural mechanisms are warranted. During tool development/ validation, cultural sensitivities and ecological validity should be carefully considered. |
| **B** | *“... She spends over eight hours each day writing and has recently published a book...”*<br>*“He plans his day and strictly follows his scheduled routine, which includes precise mealtimes, ensuring he walks 10,000 steps per day, listening to health podcasts, and rowing.”*<br>*“... he can spend his entire time with puzzles, computer games, cycling and writing religious quotes...”* | A large body of clinical studies, including the IWG dataset, based on observations by clinicians and caregivers indicates that patients spend considerable time and attention, exhibiting hyperfocus on certain activities and specific interests. However, there are no studies elucidating the neural underpinnings of hyperfocus on specific interests, nor are there objective tests available to assess this phenomenon. | Given the lack of evidence elucidating the neural mechanisms, the IWG recommends the term **“Hyperfocus on specific interests”** which is still relatively broad and does not fully reflect impaired neural domains. Yet it is more specific than current terms such as ‘mental rigidity, preoccupations, obsessions, compulsions, repetitive behavior’, and more comprehensive than single symptom-based terms such as ‘hyper-religiosity, hypergraphia etc.,’. Additionally, it clearly describes the clinical outcome. It is crucial to identify the underlying neural mechanisms. Objective, face-to-face tests targeting neural mechanisms are warranted. During tool development/ validation, cultural sensitivities and ecological validity should be carefully considered. |
| **B** | *“... Every Monday she dresses up all blue and eats only pasta...”*<br>*“...She had no musical background, but beginning two years ago, she acquired a strong preference for especially popular music and sometimes sang at home...”*<br>*“...He enjoys Chinese food now more than ever...”* | Alterations in personal preferences, such as food choices, colors, clothing, and aesthetic tastes, have been widely noted in the literature, including the IWG dataset. The current debate centers on whether these changes involve disturbances primarily in reward processing, shifts in hedonic values, or semantic loss as the underlying cause of strong personal preferences. There are no studies available using objective measurements to elucidate the neural mechanisms responsible for this clinical phenomenon. | Given the lack of evidence elucidating the neural mechanisms, the IWG recommends the term **“Altered hedonic valuation and personal preferences”** which is still relatively broad and does not fully reflect impaired neural domains. Yet it is more specific and accurate than current terms such as ‘mental rigidity, preoccupations, obsessions, compulsions, repetitive behavior’, and more comprehensive than single symptom-based terms such as ‘dietary changes, regimented diet, musicophilia etc.,’. Additionally, it clearly describes the clinical outcome. Investigating whether these modalities manifest at early stages and identifying the underlying neural mechanisms is crucial. Objective, face-to-face tests targeting neural mechanisms are warranted. During tool development/ validation, cultural sensitivities and ecological validity should be carefully considered. |

☒ Strong scientific evidence; ☐ Substantial scientific evidence, better designs needed; ☐ Mild scientific evidence, mechanisms unknown;
**B**: Symptom can be reported as a behavioral problem by the caregiver **L**: Symptom can be reported as a language problem by the caregiver **M**: Symptom can be reported as a memory problem by the caregiver;

### Box 1.

Recommended symptom checklist for clinicians

1. MULTIMODAL KNOWLEDGE LOSS FOR NON-VERBAL INFORMATION*
  A. Knowledge loss for people and other living beings
  B. Knowledge loss for flavors, odors, sounds, landmarks, and bodily sensations
  C. Knowledge loss for emotions, social information and paralinguistic cues *These deficits may be interpreted as memory, language, executive or behavioral problems by care givers.
2. ALTERED SOCIOEMOTIONAL BEHAVIOR
  A. Altered emotional expression
  B. Altered social reaction
  C. Altered motivation for social interactions
3. ALTERED PRIORITIZATION
  A. Hyperfocus on specific interests
  B. Altered hedonic valuation and personal preferences
4. SPARED FUNCTIONS
  A. Spared visuospatial functions compared to healthy controls
  B. Relatively spared attention and executive functions compared to bvFTD
  C. Relatively spared episodic memory performances compared to AD
  D. Relatively spared verbal semantic skills compared to svPPA

## 1. MULTIMODAL KNOWLEDGE LOSS FOR NON-VERBAL INFORMATION

The IWG clustered the following symptoms under this broad category, pointing out that the main deficit is ’knowledge loss’ for many categories (each discussed in detail below) via multimodal (more than one modality, i.e., not only visual) ’non-verbal’ stimuli (without using language, i.e., visual, auditory, gustatory, or olfactory).

### 1.1. RATL and knowledge loss for people

Face recognition deficits are prominently observed in patients with predominant RATL atrophy, and extensively studied across various research groups (Table 1). While commonly labeled as “prosopagnosia,” recent literature employing rigorous face-to-face assessments indicates that RATL-related impairments differ significantly from those related to posterior cortical areas primarily involved in basic face perception. Instead, these deficits are associated with a multimodal loss of person-specific knowledge, encompassing recognition difficulties with voices, biographical details, and faces, collectively termed as “person-specific knowledge” or “person-based semantics”^8,17,25–40^. These studies have shown that patients with RATL atrophy often demonstrate intact basic face perception and discrimination skills, but struggle to identify people from photographs or recognize them by voice. They therefore also exhibit deficits in assessing familiarity, semantic associations and providing semantic information. Some studies have reported that knowledge of infrequent acquaintances or famous public figures deteriorates earlier than that of close family members, analogous to the early loss of less familiar/frequent words in the left hemisphere counterpart, svPPA^34^. Several authors have highlighted that semantic knowledge about people can be spared when individuals are provided with names or verbal definitions rather than pictures^29,32,34–36,41^. Moreover, comparative studies of RATL versus LATL atrophy in semantic dementia reveal distinct patterns of impairment. A strong anatomical correlation has been found between face-to-name and voice-to-name matching performance and the right temporal lobe but not the left^42^, and patients with predominant LATL atrophy identify faces from pictures (visual task) better than their names (verbal task), whereas those with RATL atrophy show the opposite pattern^28,30,31^, suggesting the crucial role of LATL in verbal semantics, and RATL in non-verbal processing. These laterality effects are likely to be a matter of degree, rather than binary distinctions^43^ and may dissipate as disease progression leads to bilateral ATL damage. In a detailed case study, the patient was also unable to describe her feelings about the ‘recognized’ person, as an aspect of semantic loss, beyond difficulties in providing biographical information. In addition, she used general semantic knowledge to identify people. For example, she would promptly recognize her grandson in his mechanic’s overall but not in ordinary clothes. Similarly, she would identify the local priest only when he wore his black cassock during church services^35^. These examples highlight the complexity of patients’ adaptive mechanisms, underscoring the difficulties in capturing such symptoms unless they are systematically inquired about and objectively tested. Existing face-to-face tests are mainly available for Western populations’ references images and delivered in English. Cultural adaptation of those tests is essential, both in language and in the cultural familiarity of test materials. Additionally, current validated tools assess person identity via static pictures. Novel ecologically valid, and culturally sensitive tools that combine visual and auditory information are warranted (Table 2).

### 1.2. RATL and knowledge loss for other living beings

Object recognition deficits, naming, and word-finding difficulties are other common symptoms reported by several groups^6,10,22,44^. However, studies using standardized tests indicate that these deficits are category-specific. Beyond person-based knowledge, research has shown that patients with RATL atrophy experience knowledge loss for living beings, leading to recognition, naming, and word-finding difficulties^35,45–48^. Comparative studies have demonstrated that while left-predominant SD affects both animate and inanimate words equally due to language involvement, right-predominant SD, with greater language sparing, continues to impair other semantic aspects related to animals^45^. Other studies using multi-modal stimuli reported poor knowledge of sensory attributes and consistently greater impairment for living things^48,49^. A more specialized study documented difficulties in recognizing birds by their calls in a bird expert with RATL atrophy^47^, while another reported a deficit in mushroom identification in an experienced mushroom gatherer with RATL atrophy^35^. This evidence highlights the need for more detailed cognitive assessments to identify such deficits in patients with RATL atrophy. Better test designs and prospective multicultural studies are needed to elucidate whether the impairment is confined to socioemotionally relevant living beings or extends to all living entities, and to assess if this symptom is an early marker of the syndrome. Additionally, given the reported similar deficits in patients with predominant LATL atrophy^50^, prospective comparative studies are warranted to elucidate whether these deficits are part of a broader conceptual system underpinned by a bilaterally-implemented, functionally-unitary semantic hub in the ATLs. Currently, a few experimental tasks are available, but there is no validated tool for clinical use. During tool development and/or validation, cultural sensitivities should be carefully considered (Table 2).

### 1.3. RATL and knowledge loss for landmarks, monuments and places

Deficits regarding landmarks and monuments have been reported by several groups, despite the use of heterogeneous terminologies for the ensuing phenomena, including ‘getting lost,’ ‘topographagnosia,’ ‘wayfinding difficulties,’ ‘knowledge loss for places’ or ‘semantic deficit for landmarks’^10–12,22,25,26,34,38^. Although most studies rely on clinical observations, some groups have assessed this deficit using quantitative tests, reflecting knowledge loss for landmarks and monuments^25,26,34,38^. One detailed case study provided deeper insights into the nature of the deficit in RATL degeneration^26^. In this study, the patient’s wayfinding abilities in a familiar environment (i.e., his hometown) were preserved despite an inability to recognize familiar and famous buildings, monuments, and landmarks. Wayfinding was achieved through heavy reliance on written indications (e.g., names of restaurants and streets), preservation of a pre-existing cognitive map of the familiar environment, normal executive functions necessary to plan the execution of a given trajectory, and an over-reliance on processing local features. This distinguishes the deficit from navigational impairment following damage to the medial temporal lobe. Naming (4/20) and identifying (6/20) famous monuments, as well as familiarity, were significantly impaired when presented with photographs. In contrast, upon verbal presentation of the names of these famous monuments, he correctly identified 17/20 of them. Although not systematically tested initially, the patient was also unable to recognize pictures of famous places he had visited and pictures of famous monuments in his hometown^26^. Considering the high prevalence of place orientation deficits and getting lost in Alzheimer’s disease (AD), improved terminologies and standardized assessments are crucial for differentiation and accurate diagnosis of FTD patients with RATL predominance. It is crucial to investigate in multicultural cohorts whether these modalities manifest at early stages. Although some experimental tasks are currently available, there is a lack of validated clinical tools. It is crucial to consider cultural sensitivities during the development and validation processes (Table 2).

### 1.4. RATL and knowledge loss for flavors and odors

Clinical studies based on retrospective analyses of medical records frequently identify this deficit^4,28,35,47,51,52^. Although this domain has not been as extensively studied by employing objective tests, as ‘person knowledge,’ substantial evidence indicates that patients with predominant RATL atrophy experience semantic knowledge loss for flavors and odors, tested with gustatory and olfactory stimuli, despite retaining intact taste and smell perception abilities^14,17,53^. One study reported impaired identification of food elements and the inability to associate them with semantically related content (e.g., edible *versus* inedible items), despite recognizing them when provided with their names^35^. Another study described a patient who noted his favorite food smelled strange, and the slightest food odor became intolerable, described as “foul,” “rotten,” or “like sewage,” leading to significant weight loss due to reduced food intake^14^. Several authors suggest that “the strong food preference” observed in RATL neurodegeneration may be due to semantic degradation for foodstuffs, narrowing their preferences^14,53^. However, it remains debated whether chemosensory alterations (taste, smell) represent a pure semantic deficit or a broader deficit in hedonic valuation^24^. It is necessary to objectively test whether deficits related to taste and smell stimuli constitute a multimodal non-verbal semantic deficit and if so, whether these deficits are category-specific (e.g., living beings, social context-related items) in larger samples. Furthermore, although such deficits have been reported as early symptoms of the syndrome, prospective multicultural comparative studies are needed to elucidate the contribution of LATL atrophy and to determine the impact of the culture in food appreciation, as well as the onset and distinctiveness of these symptoms. At present, a few experimental tasks exist, but no tools have been validated for clinical application. Attention to cultural nuances is essential throughout development and validation (Table 2).

### 1.5. RATL and knowledge loss for sounds

Studies focusing on RATL degeneration and sounds have primarily examined human voices, as discussed above (see section 1.1.). However, data using detailed assessments that include otorhinolaryngological examinations and tests for perception, identification, discrimination, familiarity, semantic association, and naming have revealed knowledge loss for accents, songs, vocal tones, and melodies as well as prosody, in patients with RATL neurodegeneration^39,54–58^. Neuroimaging studies have found significant correlations between famous music recognition deficits and right temporal pole atrophy^56^, and between atrophy in the right supra-marginal and superior temporal gyri and deficits in detecting violated sounds and melodies. Additionally, atrophy in the bilateral anterior temporal poles and left medial temporal structures was related to deficits in environmental sound recognition^55^. Furthermore, post-hoc analysis in this study showed that the right predominant SD group had significant impairments in both melody and environmental sound tasks, scoring lower for living superordinate categories (animals, humans), although this was not statistically significant^55^. However, given the limited amount of evidence, this domain still awaits better-designed studies to determine its category specificity, clinical prevalence in the early stages of the syndrome, and distinctiveness from LATL neurodegeneration. The field has some experimental tasks but lacks clinically validated tools. Cultural considerations should be integral to the development and validation phases (Table 2).

### 1.6. RATL and knowledge loss for bodily sensations

Another ubiquitous symptom in the RATL literature is altered responses of patients to sensations, mostly termed as “somatization”, “hypochondria”, or “alexisomia”^10,59^. The lack of knowledge related to interoceptive stimuli and their interpretation, leading to misidentification of normal bodily sensations, has been suggested as the underlying mechanism^18,59^. Patients exhibit a variety of complaints of behaviors related to this impairment, including unidentified pains, aches, numbness, itching, tinnitus, discomforts; feelings of warmth or cold; abnormal sensations in the bladder, bowel, thorax, abdomen, head and stomach; inappropriate reactions to own bodily odors, hunger, regurgitation, borborygmi, running nose, sweating, and fatigue. Normal interceptive signals may be considered as indicators of disease, which may be reported as hypochondriasis, illness anxiety disorder, or Cotard syndrome^10,18,59^.

Recent studies using cardiac monitoring during emotional stimuli have shown significantly impaired interoception in predominant RATL neurodegeneration compared to those with predominant LATL atrophy^60^. The authors suggested that impaired emotion recognition in the RATL syndrome is driven by inaccurate internal monitoring^60^. Conversely, another group found preserved cardiac reactivity during emotional stimuli in ATL syndromes, whereas cardiac reactivity was attenuated in groups with predominant fronto-insular atrophy (bvFTD and nonfluent PPA)^61^. Another group suggested that a decline in the parasympathetic nervous system may contribute to reductions in interpersonal engagement and gregariousness/extraversion, personality changes that are especially common in the RATL syndrome^62^. Nevertheless, these hypotheses need to be tested in larger cohorts to elucidate the neural mechanisms and determine whether they are early characteristic symptoms of the syndrome and whether the bodily sensation of emotion dissipates before the cognitive recognition of the emotion, or vice versa^63^. As discussed in the previous sections, a consistent theme emerges: while there are a few experimental tasks available, the absence of validated tools for clinical use remains a significant gap. Moving forward, it is imperative that cultural sensitivities are incorporated into every stage of tool development and validation (Table 2).

### 1.7. RATL and knowledge loss for emotions, social information and paralinguistic cues

Emotion recognition deficit is one of the most frequently reported symptoms in patients with RATL atrophy. Various emotion recognition tasks (see Table 1) have been used globally, confirming these deficits across different studies^22,60,64–67^. Comparative studies have shown that patients with right temporal predominance at early stages perform worse on facial emotion selection tasks compared to other FTD subtypes, including bvFTD and svPPA^22,67^. Recent work has shown that comprehending facial cues is not limited to emotions. A study using video-based dynamic stimuli (including facial expressions, body gestures, and vocal cues) demonstrated a sarcasm detection deficit in patients with predominant RATL atrophy, who considered the actors’ statements literally without reading paralinguistic cues. This deficit was worse in RATL patients than in their left temporal and frontal FTD counterparts^22^. A PET study indicated that FTLD patients with right superior ATL hypometabolism were significantly more impaired on social concepts (e.g., ‘polite,’ ‘stingy’) than on animal function concepts (e.g., ‘trainable,’ ‘nutritious’)^68^. A case report assessing knowledge of social information using a standardized test of general knowledge of public events and figures, found that the patient’s scores were significantly impaired in the visual identification part of the semantic test (a picture of a famous public event, such as the explosion of the atomic bomb, the destruction of the Berlin Wall, etc.,) while performance was within the normal range for the verbal part of the test^69^. Another study using a social interaction vocabulary test found a correlation between bilateral ATL atrophy and lower performances on this test. Notably, in this test, the definition of the social interaction was provided verbally, and participants were asked to match the word with a picture^70^. These studies support the argument that while the RATL is crucial for non-verbal comprehension of socioemotional concepts (paralinguistic, visual, vocal), the LATL supports verbal comprehension^71,72^. Existing face-to-face tests are mainly available for Western, English-speaking populations. During tool development and/or validation, cultural sensitivities and ecological validity should be considered (Table 2).

## 2. ALTERED SOCIOEMOTIONAL BEHAVIOR

A second domain affected in FTD with RATL predominance according to the IWG was altered socioemotional behaviors, characterized by either exaggerated responses or a lack of reaction, or altered motivation for social interactions.

### 2.1. RATL and altered emotional expression

Beyond emotion recognition deficits, several groups have reported other social cognition deficits such as inappropriate emotional expression (facial and prosodic) or theory of mind (ToM) (mentalizing) impairments in patients with RATL atrophy. This is often described as a ‘lack of empathy’ by caregivers and clinicians, ^13,16,20,22,73–76^. A recent study found a restricted prosodic range in this group compared to svPPA and healthy controls, which was associated with a reduction in empathy, as observed by caregivers^13^.

Another study showed that the subgroup with RATL atrophy exhibited a unique phenotype, characterized by globally reduced facial reactivity and aberrant coupling of muscle reactivity to facial expression identification. This group showed impaired emotion recognition, but their facial mimics were not correlated with emotion identification performances, unlike other FTD subgroups and healthy controls, and were described as ‘poker-faced (no reaction)’ or ‘caricatures’ of normal emotional reactions (over/inappropriate reaction)^16,74^. Similarly, another group reported that compared with healthy older adults, bvFTD patients showed an overall dampening of physiological responses, whereas SD patients exhibited abnormal facial expressiveness discordant with the emotional content of the stimuli^73^. The right fusiform gyrus has been implicated in the processes of facial and emotional recognition of laughter, explaining inconsistent shared laughter experiences in patients with RATL^75^. Additionally, ToM deficits on ToM tasks have been associated with RATL regions, a recent study assessing a large number of patients with focal RATL atrophy at early stages using dynamic face-to-face tests showed that patients with predominant RATL atrophy displayed lower scores in the emotional ToM task, unlike patients with predominant frontal atrophy who exhibited worse performance in the cognitive ToM task^22^. It has been suggested that if an individual can no longer understand the semantic meaning of emotional information, the associated response appears compromised^71,73^. However, it remains to be tested whether these altered emotional reactions are purely due to knowledge loss for emotions or whether alterations in other brain networks following neurodegeneration in RATL lead to the abnormal behavioral response. Given the lack of evidence elucidating the neural mechanisms, the IWG recommends the term “altered emotional expression”. Although it is still relatively broad and does not fully reflect impaired neural domains, it refers to the objective observation of the external expression of emotion, rather than attempting to infer the patient’s internal emotional state. Additionally, it is more specific than current terms such as ‘lack of empathy’, and clearly describes the clinical phenomenon. Next to neural mechanisms, it is crucial to investigate whether these modalities manifest at early stages. The same gap persists regarding the lack of culturally sensitive validated tests for clinical use (Table 2).

### 2.2. RATL and altered social reaction

Previous work has shown that knowledge loss for socioemotional information as well as people, living beings, landmarks, flavors, odors, sounds and bodily sensations was mainly reported as ‘disinhibition’ by many clinicians, as the clinical outcome was socially inappropriate behavior^4,71^ (i.e., drinking a bottle of soap, confusing it with food, see more real life examples in Table 2). This has been widely discussed in several theoretical models, suggesting that conceptual knowledge of social constructs and socially relevant cues are represented in the ATLs^65,71,72,77^, and an anatomical model for the umbrella term “disinhibition” was offered suggesting that ATL related disinhibition is associated with loss of knowledge of social norms and expectations rather than a problem with control or inhibition *per se*^78^. However, no large sample size study has yet unveiled the neural components of inappropriate social behavior in RATL using objective measurements. It remains unknown whether there are other components causing such reactions beyond semantic deficits. Given the lack of evidence elucidating the neural mechanisms, the IWG recommends the term “altered social reaction,” which is still relatively broad and does not fully reflect impaired neural domains. Yet it is more specific and accurate than current terms such as ‘disinhibition’, and clearly describes the clinical phenomenon without implying the unknown mechanism. Currently, no tasks are available for clinical use.

### 2.3. RATL and altered motivation for social interactions

Apathy is another highly reported symptom in RATL cohorts, mainly measured with the neuropsychiatric inventory apathy subscale^10,79–83^. However, the characteristics of ‘lack of motivation’ in the RATL syndrome differ from classic cognitive apathy, which typically involves losing motivation for almost all daily tasks and novelty seeking. Instead, patients exhibit a strong shift from socially motivated activities to narrowed solitary activities for which they show heightened motivation^4,23^ (see section 3). Moreover, in the early stages, patients maintain their non-social daily life activities such as showering, cooking, driving etc., but they may express inertia towards social activities^4^. Available studies using quantitative assessments are limited, however, one study using the Snaith-Hamilton Pleasure Scale and the motivation subscale of the Cambridge Behavior Inventory indicated a unique role for right temporal lobe structures in modulating anhedonia in SD. The study suggested that degeneration of predominantly right-hemisphere structures deleteriously impacts the capacity to experience pleasure in SD, leading to a lack of motivation^84^. However, no distinction regarding social vs solitary activities has been made in this study. Moreover, patients with semantic and behavioral deficits violate the assumptions implicit in self-report questionnaires and Likert scales^85^. Although there are no objective measurements to assess such behaviors, clinical studies indicate that the motivation for social interactions is often misplaced rather than completely absent. Given the lack of evidence elucidating the neural mechanisms, the IWG recommends the term “Altered motivation for social interactions,” which is still relatively broad and does not fully reflect impaired neural domains. Yet it is more specific than current complex terms such as ‘apathy’, and clearly describes the clinical phenomenon without implying a mechanism. Investigating whether these modalities manifest at early stages and identifying the underlying neural mechanisms is crucial. Beyond informant-based surveys for broad symptoms such as pleasure, apathy, motivation scales, more specific, objective, culturally sensitive face-to-face tests targeting neural mechanisms are warranted (Table 2).

## 3. ALTERED PRIORITIZATION

Another symptom group that is very prominent and even characteristic of patients with RATL is altered prioritization. This category includes symptoms such as hyper-religiosity (excessive preoccupation with religious activities, e.g., spending considerable time at church reading the Bible), developing strong appreciation for certain topics (e.g., musicophilia), rigidity around food (e.g., only eating spaghetti), color (e.g., only wearing blue), or scheduling (e.g., eating breakfast only at 8 a.m.). Instead of describing this symptom group based on individual examples, as has been done previously in the literature, the IWG conceptualized it under the title ‘altered prioritization’, categorizing it into two subgroups: hyperfocus on specific interests and altered hedonic valuation and personal preferences.

### 3.1. RATL and hyperfocus on specific interests

Perhaps the least understood symptom group of the syndrome is mental rigidity, preoccupations, ritualistic and obsessive-compulsive behavior, despite their high prevalence (78% in early stages) in the IWG dataset^4^. A total of 505 specific examples were reported, including time and schedule (21%), food (17%), puzzles/sudoku/computer games (12%), global warming/recycling/saving gas, water, electricity (8%), sports (6%), walking/cycling/driving (6%), hoarding/collecting (5%), health-related (4%), shopping/ordering (3%), colors (3%), clothes (3%), religion (3%), writing (2%), art (music/drawing/painting/sculpture) (2%), saving money/parsimony (2%), cleaning (1%), clock-watching (1%), checking/controlling (1%), gardening (1%), and other (4%). Similar examples have been described by several authors displayed in Table 1. These activities are part of patients’ daily lives, and they exhibit heightened motivation and attention spans toward such actions. There is no objective test available to assess such behavior; however, caregiver reports and clinical observations suggest a unique nature, indicating that patients spend considerable time and attention, exhibiting hyperfocus on certain activities and specific interests. Additionally, unlike individuals with psychiatrically diagnosed obsessive-compulsive disorder, patients with RATL atrophy exhibit less anxiety, self-criticism, or insight. Thus, the IWG advocates for improved terminologies instead of mental rigidity, preoccupations, ritualistic or obsessive-compulsive behavior to better phenotype the distinct characteristics of these symptoms. Future studies are warranted to identify the neural underpinnings of such deficits, to interrogate the thought process behind these typical behaviors, and objective tests are needed to examine these symptoms in daily clinical practice.

### 3.2. RATL and altered hedonic valuation and personal preferences

Alongside specific interests, alterations in personal preferences such as food choices, colors, clothes, and aesthetic tastes have been noted in international data^4^. A group of authors has suggested that the clinical syndrome associated with RATL atrophy may partly involve disturbances in reward processing, shifting hedonic values away from people towards inanimate objects^23,86^. Similar arguments have highlighted that semantic knowledge of social interactions is influenced by the hedonic evaluation system, emphasizing the close connection between ATLs and the medial orbitofrontal regions^70,77^. Several clinical scientists claimed semantic loss as the reason of strong personal preference, particularly for food^14,17,53^. However, to what extent semantic deficits contribute to personal preferences and whether valence processing (i.e., recognizing the pleasant or unpleasant nature of emotions) also depends on semantic knowledge remains to be determined through objective evaluations (Table 2).

## 4. OTHER SYMPTOMS

### 4.1. RATL and apparent memory deficits

To date, episodic, semantic and autobiographical memory deficits have been documented in RATL with discrepant frequencies by several groups^10,11,87–89^, and the occurrence of amnestic presentations (episodic memory deficits), remains a controversial topic in the field. Although episodic memory disturbances (i.e., forgetting appointments) have been reported in multiple studies^4,10,88,90–93^, these were objectified with episodic memory tests in relatively fewer studies^4,10,76,94^. The latter showed impairments in standard episodic memory tests, particularly in those using visual stimuli rather than verbal stimuli. When comparing dementia subtypes, worse semantic memory performances in FTD with RATL atrophy were found compared to AD and worse episodic memory performances in AD compared to FTD with RATL atrophy^10,76,94^. In IWG’s recent study, chart reviews showed that 67% of patients had reported memory problems, whereas objective abnormalities varied between 21% and 87% across eight different episodic memory tests^4^.

Studies using detailed memory tests have identified category specific memory deficits^25,95^. In those studies, memory for famous people and social events were selectively disturbed. Current neuroscientific evidence suggests that due to the categorization problem, patients with semantic deficits demonstrate a “over-generalization” tendency and exhibit learning difficulties^96^, and semantic processing may underlie forms of episodic and autobiographical memory by providing schemas and meaning for remembering the past, even for imagining the future^89^. However, those publications include patients with either predominant LATL or bilateral temporal atrophy. Given the advent of disease-modifying therapies for AD, the IWG calls for more focused studies that aim to disentangle the neural and molecular underpinnings of memory impairment in RATL syndrome.

### 4.2. RATL and psychiatric symptoms

Besides those core symptoms, affective dysregulation, anxiety/panic, delusions/ hallucinations, have also been reported with lower frequencies in previous publications^10,88^. Although, former literature suggested depression as a distinctive symptom^10,88^, our joint data also showed cases with mania and fluctuating mood^4^. A case with RATL atrophy whose severe claustrophobia had disappeared 7 years after onset of her first symptoms has been reported^97^. Another case with mania responded very well to symptomatic treatment ^98^. On the other hand, another group has drawn attention to increased potential risk for suicidal behavior in this disease group, as they found preoccupations around depressive thoughts and suicidal ideas^99^. All reported psychiatric symptoms in the literature rely on clinical observations and caregiver declarations, lacking objective assessments to better understand the neural mechanisms causing such problems. However, RATL neurodegeneration should be considered in cases with late-onset psychiatric problems, and other RATL-specific symptoms listed in this paper should be further assessed to detect potential underlying neurodegeneration.

### 4.3. RATL and language problems

As discussed in the previous sections and exemplified in Table 2, the loss of knowledge across several categories were reported as language problems by many caregivers and clinicians. In the IWG dataset, 70% of patients were reported to have naming and word-finding difficulties, although available cognitive test scores revealed that nearly all patients who underwent cognitive assessment exhibited severe visual and person-specific semantic deficits while performances on general naming were relatively better^4^.

Additionally, comparative studies have shown that patients with predominant RATL atrophy performed better on verbal semantics and fluency tests compared to svPPA, however worse than bvFTD and healthy controls^10,22^. Therefore, further work is required to understand the role of RATL in language functions, particularly in naming and elucidate the contributions of LATL atrophy which is commonly observed in patients with RATL predominant atrophy.

### 4.4. RATL and spared functions

It should be noted that almost all papers have reported no symptoms related to visuospatial and attention functions^4,6,8–10,22,88,100^. Patients have either performed within normal expectations or showed mild impairment on standardized tests. Visuospatial functions, in particular, have been highlighted as well-preserved, a finding confirmed by standardized test assessments^4,6,8–11,22,88^. Comparative studies showed although executive and attention functions may not be normal, they are less severely affected compared to AD, bvFTD, and svPPA^10,22^. And in RATL, there are better verbal semantic skills, less surface dyslexia in English; or fewer accent and tone regularization errors in other languages compared to svPPA^10,22,79^, and better episodic memory performances compared to AD^10,76,94^.

## CONCLUSIONS AND FUTURE DIRECTIONS

This consensus paper on FTD with RATL predominant neurodegeneration breaks new ground on four levels. First, it represents the first international initiative, employing a 4-year nominal group approach that includes neurologists, psychiatrists, psychologists, and dementia neuroimaging experts from around the world as equal partners to tackle discrepancies and resolve conflicts in the field.

Second, it implements a meticulous systematic review aimed at identifying patients with RATL atrophy harmonized in SD, svPPA, and bvFTD cohorts, resulting in the largest collection of cases to disentangle the nature of the symptoms in a neuroscientifically informed manner. Third, it provides transparent, evidence-based nomenclature that moves beyond subjective caregiver and clinician observations, clearly highlighting limitations and avoiding the imposition of personal opinions. Lastly, it identifies the lack of or limited evidence in many domains, indicating areas where physicians need more information, thus shaping future direction goals.

Our future goal is to perform cross-cultural validation of the clinician-faced symptom checklist identified in this study (Box 1) by utilizing culturally sensitive objective tests described in Table 2. This study will include direct comparisons with other diagnostic groups to differentiate the syndrome from AD and psychiatric disorders, and delineate the ambiguous boundaries with bvFTD, and particularly with left predominant SD/svPPA.

### Contributors

HU and YP established the international working group (IWG) and the study was designed by them with the contributions of the IWG. KY, MM and HU conducted the systematic review. HU collected and integrated the qualitative data (expert recommendations) generated by IWG. HU wrote the manuscript. HU designed the figures and tables. All authors contributed to the interpretation of the results and provided a critical review and approval of the manuscript.

### Role of the funding source

The funders of the study had no role in study design, data collection, data analysis, data interpretation, or writing of the report. The corresponding author had full access to all study data and had final responsibility for the decision to submit for publication.

### Declaration of interests

Authors declare no competing interest.

**\*Investigators of the IWG**

**Agustin Ibanez**^31^**, Alan Lerner**^32^**, Alexander Frizell Santillo**^12^**, Alexandre Morin**^33^**, Alma Ghirelli**^34^**, Andrea Arighi**^24^**, Arabella Bouzigues**^28^**, Bedia Samancı**^9^**, Bradley F. Boeve**^27^**, Bruce L. Miller**^2^**, Carmela Tartaglia**^35^**, Caroline Dallaire Theroux**^36^**, Christopher Kobylecki**^11^**, Daniel Ohm**^37^**, Daniela Galimberti**^24,25^**, David Foxe**^6^**, David Irwin**^37^**, David Perry**^2^**, Diana Matallana Eslava**^38^**, Edoardo Gioele Spinelli**^34^**, Elisa Canu**^34^**, Elisabet Englund**^13^**, Emily Rogalski**^25^**, Emma Devenney**^6^**, Emma Rhodes**^37^**, Eun Joo Kim**^39^**, Federica Agosta**^34^**, Fiona Kumfor**^6^**, Florence Pasquier**^5^**, Floor Duits**^1^**, Francesco Di Lorenzo**^40^**, Frederik Barkhof**^1^**, Gail Robinson**^41^**, Giorgio Fumagalli**^7,8^**, Giuseppe Piga**^42^**, Gregory Kuchcinski**^5^**, Hakan Gurvit**^9^**, Halle Quang**^6^**, Harro Seelaar**^43^**, Howie Rosen**^2^**, Hulya Ulugut**^1,2^**, Ignacio Illan Gala**^10^**, James Rowe**^30^**, Jan Van den Stock**^16^**, Janine Diehl Schmid**^15,22^**, Jason D. Warren**^28^**, Jennifer C. Thompson**^11^**, Jennifer L. Whitwell**^26^**, Jessica Hazelton**^6^**, Jonathan D. Rohrer**^28^**, Julie Fields**^25^**, Julie Snowden**^11^**, Julien Lagarde**^44^**, Jwala Narayanan**^45^**, Katherine P. Rankin**^2^**, Katya Rascovsky**^37^**, Keith A. Josephs**^28^**, Kristina Horne**^6^**, Kyan Younes**^2,3^**, Leonardo Cruz de Souza**^20^**, Leonel T. Takada**^21^**, Lina Riedl**^15^**, Lize Jiskoot**^43^**, Lucy Russell**^28^**, Luca Sacchi**^24^**, Manuela Pintus**^24^**, Maria Landqvist Waldo**^14^**, Marilu Gorno Tempini**^2,29^**, Mario F. Mendez**^46,47^**, Marsel Mesulam**^25^**, Massimo Filippi**^34^**, Mathieu Vandenbulcke**^16^**, Matthew Jones**^11^**, Matthew Rouse**^30^**, Masud Husain**^48^**, Matthias L. Schroeter**^49^**, Maud Tastevin**^18^**, Maxime Bertoux**^5^**, Maxime Montembeault**^4^**, Mira Didic**^50^**, Muireann Irish**^6^**, Murat Emre**^9^**, Na-Yeon Jung**^39^**, Oliver Piguet**^6^**, Oskar Hansson**^12^**, Paulo Caramelli**^20^**, Peter Pressman**^19^**, Raffaella Migliaccio**^44^**, Ratnavalli Ellajosyula**^45^**, Rik Ossenkoppele**^1^**, Rik Vandenberghe**^17^**, Robert Jr Laforce**^18^**, Samantha Loi**^51^**, Sandra Weintraub**^25^**, Shalom Henderson**^30^**, Sid Ramanan**^30^**, Sian Thompson**^30^**, Simon Ducharme**^4^**, So Young Moon**^52^**, Sun Min Lee**^52^**, Thibaud Lebouvier**^5^**, Toji Miyagawa**^27^**, Virginia E. Sturm**^2^**, Welmoed Krudop**^1^**, Yolande Pijnenburg**^1^

^1^Alzheimer Center Amsterdam, Department of Neurology, Amsterdam Neuroscience, Vrije Universiteit Amsterdam, Amsterdam UMC, Amsterdam, The Netherlands, ^2^Memory and Aging Center, Department of Neurology, UCSF Weill Institute for Neurosciences, University of California, San Francisco, USA, ^3^Stanford Neuroscience Health Center, Department of Neurology, Stanford University, Palo Alto, USA, ^4^Department of Psychiatry, Douglas Mental Health University Institute, McGill University Health Centre, McGill University, Montreal, Canada, ^5^Lille Neuroscience & Cognition U1172, Univ. Lille, Inserm, CHU Lille, LiCEND & Labex DistALZ, Lille, France., ^6^Brain and Mind Centre, University of Sydney, Sydney, Australia, ^7^Department of Neurology, University of Milan, Milan, Italy, ^8^Università degli Studi di Trento | UNITN · CIMEC - Center for Mind/Brain Sciences, ^9^Department of Neurology, Istanbul University, Istanbul, Turkey, ^10^Sant Pau Memory Unit, Department of Neurology, Hospital de la Santa Creu i Sant Pau, Biomedical Research Institute Sant Pau, Universitat Autònoma de Barcelona, Barcelona, Spain; Centro de Investigación en Red-Enfermedades Neurodegenerativas (CIBERNED) Madrid, Spain, ^11^Cerebral Function Unit, Greater Manchester Neuroscience Centre, Salford Royal NHS Foundation Trust, Salford, UK; Division of Neuroscience and Experimental Psychology, Faculty of Biology, Medicine and Health, University of Manchester, Manchester, UK, ^12^Clinical Memory Research Unit, Department of Clinical Sciences, Faculty of Medicine, Lund University, Lund/Malmö, Sweden, ^13^Division of Pathology, Department of Clinical Sciences, Lund University, Lund, SE-221 85, Sweden, ^14^Division of Clinical Sciences Helsingborg, Department of Clinical Sciences Lund, Lund University, Lund, Sweden, ^15^School of Medicine, Department of Psychiatry and Psychotherapy, Technical University of Munich, Munich, Germany, ^16^Neuropsychiatry, Department of Neurosciences, Leuven Brain Institute, KU Leuven, Leuven, Belgium, ^17^Neurology Service, University Hospitals Leuven, Leuven, Belgium; and Laboratory for Cognitive Neurology, Department of Neurosciences, KU Leuven, Leuven, Belgium, ^18^Clinique Interdisciplinaire de Mémoire (CIME), Département des Sciences Neurologiques, Laval University, Quebec City (QC), Canada, ^19^University of Colorado, Anschutz Medical Campus, Behavioral Neurology Section, Department of Neurology, USA, ^20^Behavioral and Cognitive Neurology Research Group, Department of Internal Medicine, Faculdade de Medicina, Universidade Federal de Minas Gerais, Belo Horizonte, Brazil, ^21^Cognitive and Behavioral Unit, Hospital das Clinicas, Department of Neurology, University of São Paulo Medical School, São Paulo, Brazil, ^22^Kbo-Inn-Salzach-Klinikum, Clinical Center for Psychiatry, Psychotherapy, Psychosomatic Medicine, Geriatrics and Neurology, Wasserburg/Inn, Germany, ^23^Department of Biomedical, Surgical and Dental Sciences, University of Milan, Milan, Italy, ^24^Fondazione IRCCS Ca’ Granda, Ospedale Maggiore Policlinico, Milan, Italy, ^25^Mesulam Center for Cognitive Neurology and Alzheimer’s Disease Northwestern University Feinberg School of Medicine, IL, Chicago, USA, ^26^Department of Radiology, Mayo Clinic, Rochester, Minnesota, USA, ^27^Department of Neurology, Mayo Clinic, Rochester, Minnesota, USA, ^28^Dementia Research Centre, UCL Queen Square Institute of Neurology, London, UK, ^29^Dyslexia Center, University of California, San Francisco, USA, ^30^Department of Clinical Neurosciences and Cambridge University Hospitals NHS Trust and Medical Research Council Cognition and Brain Sciences Unit, University of Cambridge, Cambridge, UK, ^31^Latin American Brain Health Institute (BrainLat) at Universidad Adolfo Ibáñez (UAI), ^32^Department of Neurology, Case Western Reserve University, Cleveland, OH,USA, ^33^ Department of Neurology Rouen University Hospital, France, ^34^Neurology Unit, IRCCS San Raffaele Scientific Institute, Milan, Italy, ^35^Department of Neurology, University of Toronto, Toronto, Canada, ^36^Clinique Interdisciplinaire de Mémoire (CIME), Laval University, Quebec, Canada, ^37^Penn Frontotemporal Degeneration Center, Department of Neurology, University of Pennsylvania, Philadelphia, PA, United States ^38^Department of Neurology, Javeriana University, Bogotá, Colombia, ^39^Pusan National University School of Medicine and Medical Research Institute 179 Gudeok-ro, Seo-gu, Busan, Korea, ^40^Non-invasive Brain Stimulation Unit, IRCCS Fondazione Santa Lucia, Rome, Italy, ^41^Brain & Mind Center, University of Sydney, Camperdown NSW 2050, Australia, ^42^ICM Paris Brain Institute, Paris France ^43^Department of Neurology, Erasmus MC - University Medical Centre Rotterdam, Rotterdam, The Netherlands, ^44^Department of Neurology of Memory and Language, GHU Paris Psychiatrie & Neurosciences, Hôpital Sainte Anne, Paris, France ^45^Department of Neurology, Manipal Hospital India, Bengaluru, Karnataka, India, ^46^Department of Neurology, University of California Los Angeles, Los Angeles, CA, USA; ^47^Department of Psychiatry, University of California Los Angeles, Los Angeles, CA, USA. ^48^Oxford University Hospitals NHS Foundation Trust, Oxford, UK, ^49^Max Planck Institute for Human Cognitive and Brain Sciences, Germany ^50^Department of Neurology, Aix-Marseille University, Marseille, France, ^51^Academic Unit for Psychiatry of Old Age, St. Vincent’s Health, Department of Psychiatry, University of Melbourne, Australia, Department of Neurology, School of Medicine, ^52^Ajou University, Suwon, 443–721, South Korea

## Data Availability

All data produced in the present work are contained in the manuscript

https://alz-journals.onlinelibrary.wiley.com/doi/10.1002/alz.14076

## Acknowledgements

We would like to thank to Alexandra Paizee for her help in technical details of the systematic review.

## Fundings

This project is supported by an Alzheimer’s Association Grant (AACSF-22-849085, PI: Ulugut). Josephs and Whitwell are supported by R01-AG37491, R01-DC010367, R21-NS94684. Van den Stock is supported by KU Leuven (IDN/21/010 and C24/18/095) and the Sequoia Fund for research on ageing and mental health. Santillo is supported by the Swedish federal government under the ALF agreement. Bertoux is supported by the Fondation Planiol, the Fondation France Alzheimer, and Fondation Vaincre Alzheimer. Laforce; Chaire de recherche sur les aphasies primaires progressives - Fondation de la Famille Lemaire. Warren is supported by the Alzheimer’s Society, Alzheimer’s Research UK, the Royal National Institute for Deaf People, the NIHR UCLH Biomedical Research Centre and a Frontotemporal Dementia Research Fellowship in Memory of David Blechner (funded through The National Brain Appeal). Rankin is supported by RF1-AG029577. Rowe is funded by the Medical Research Council (MC_UU_00030/14; MR/T033371/1); the NIHR Cambridge Biomedical Research Centre (NIHR203312) and Wellcome Trust (220258). Vandenberghe receives support from the Mady Browaeys Fonds voor Onderzoek naar Frontotemporale Degeneratie. For the purpose of open access, the authors have applied a CC BY public copyright license to any Author Accepted Manuscript version arising from this submission.

---

**Table 1.**
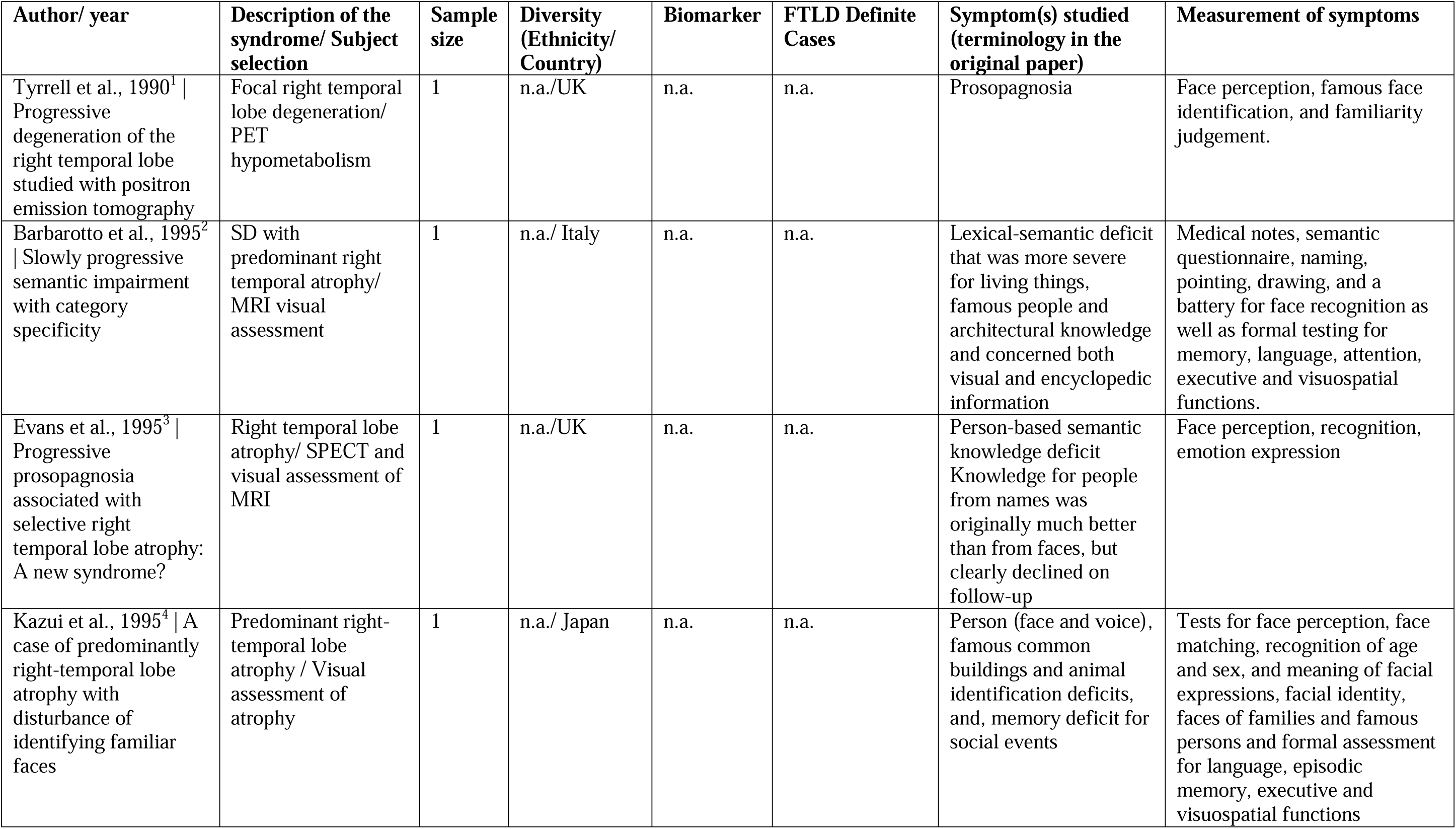
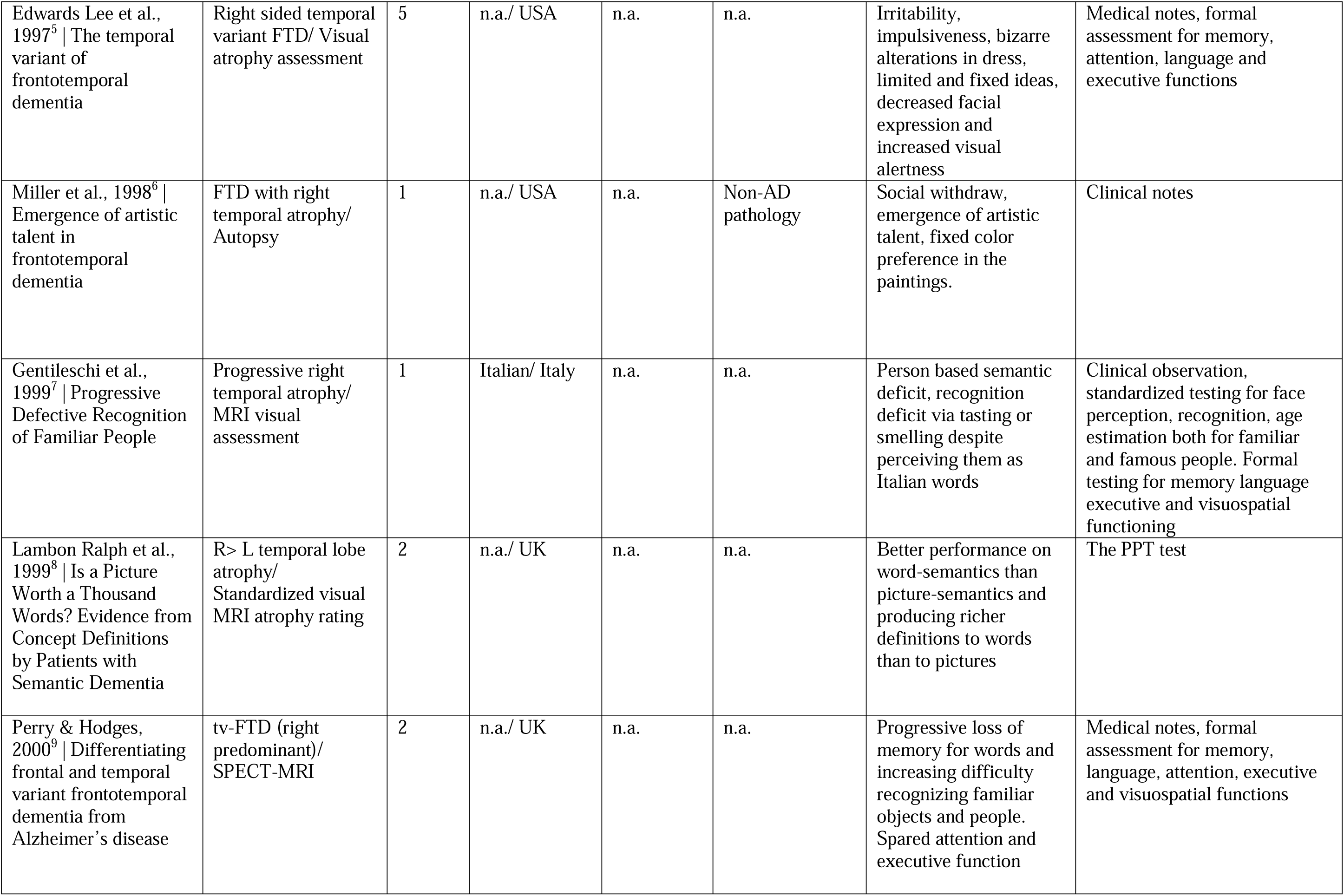
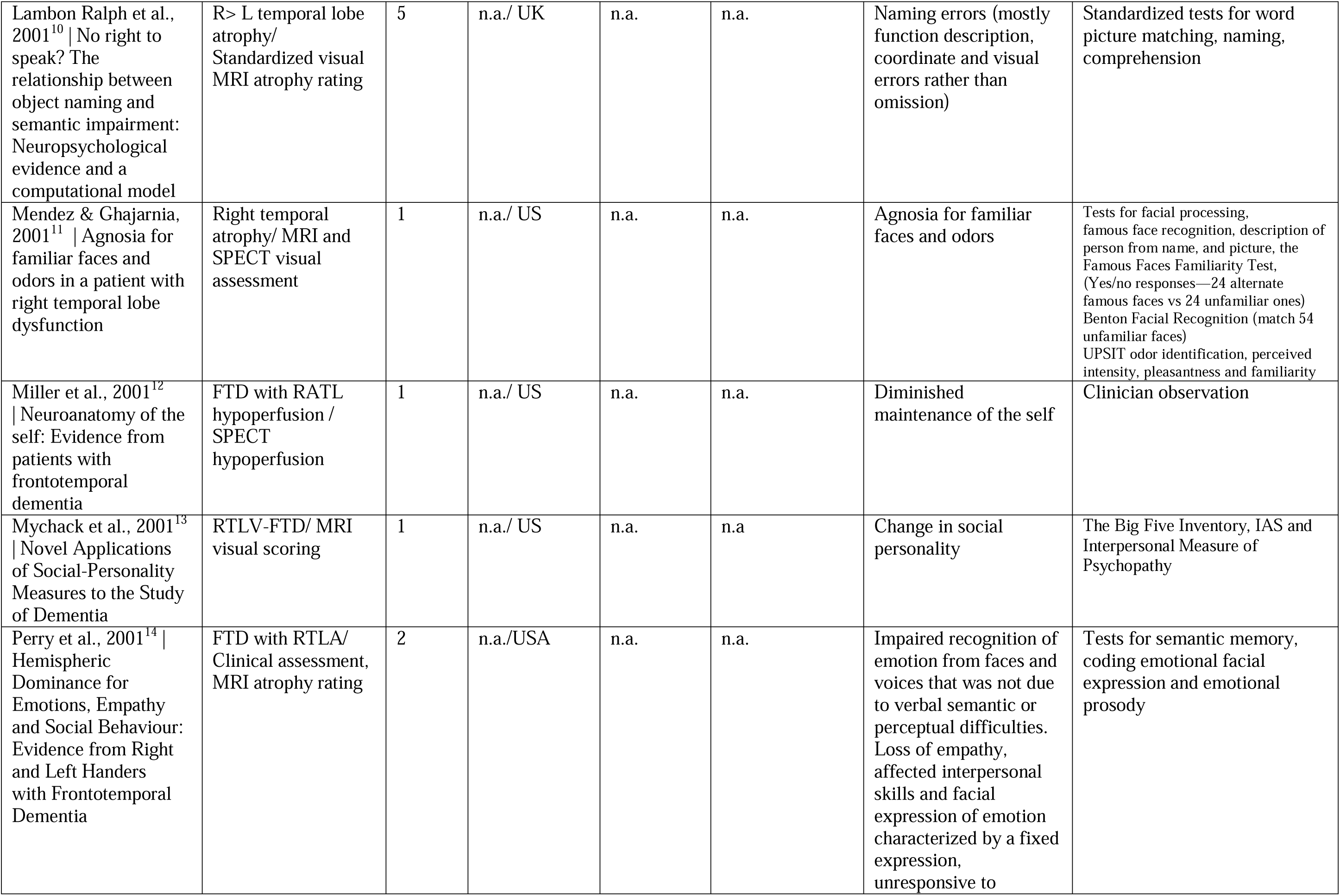
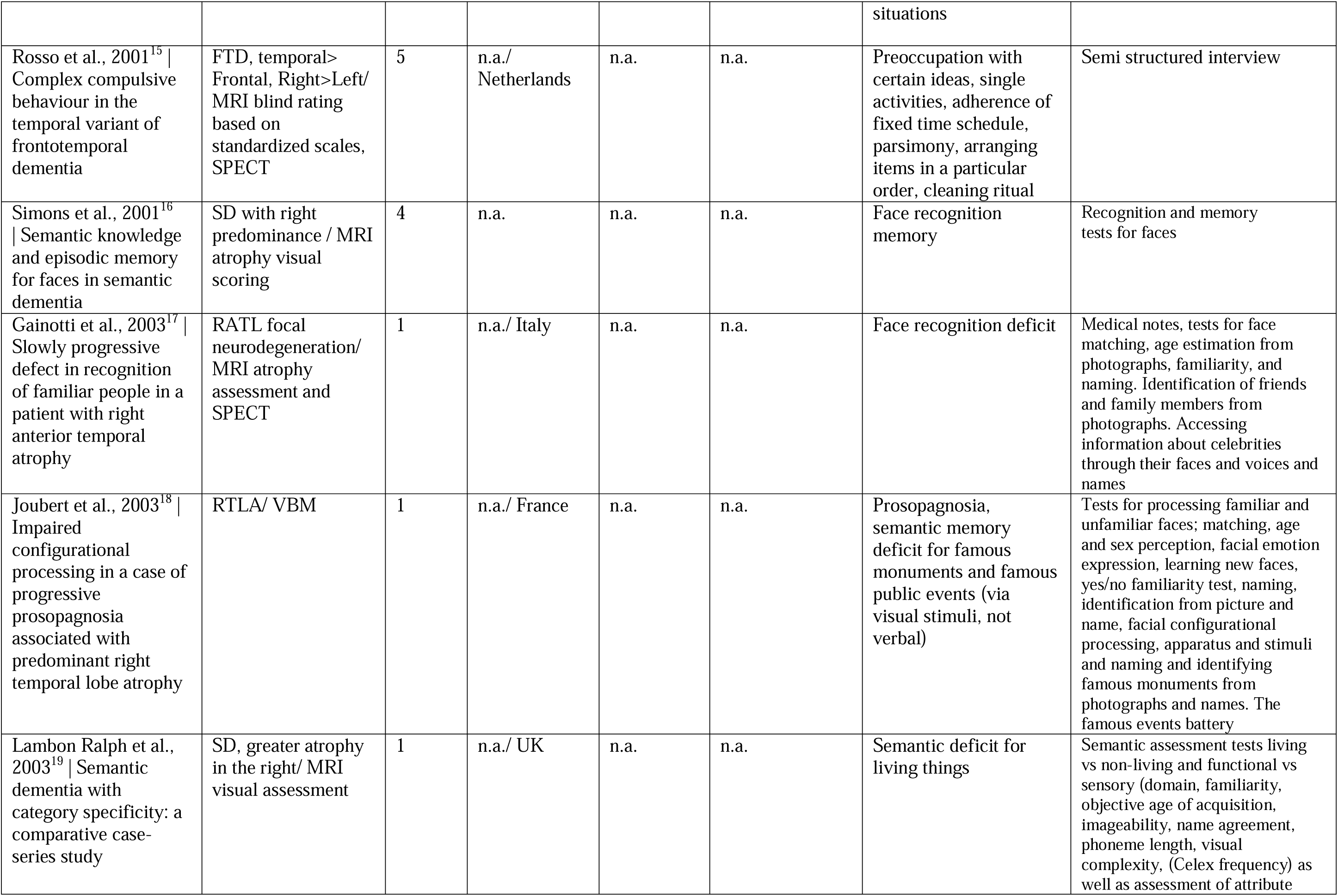
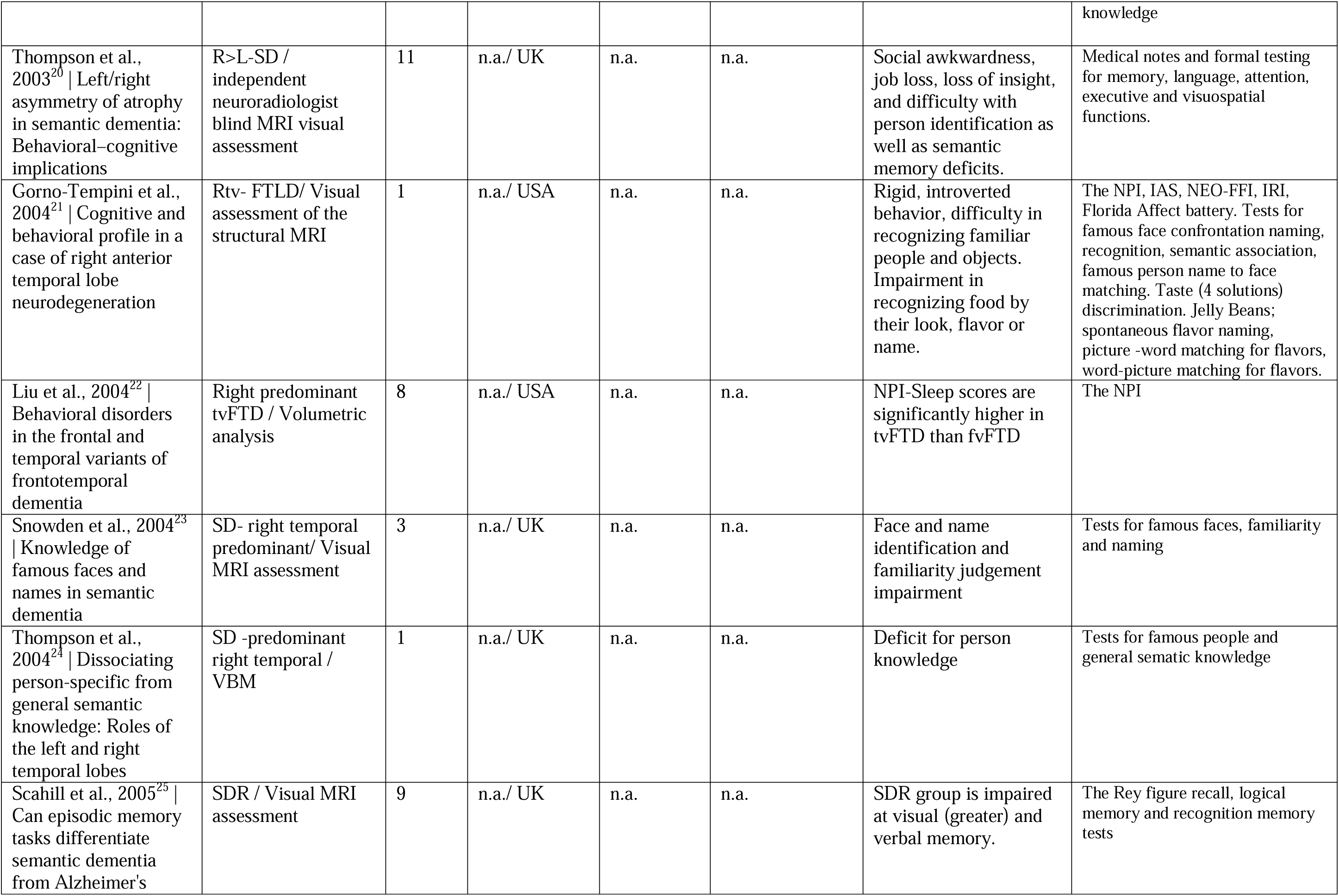
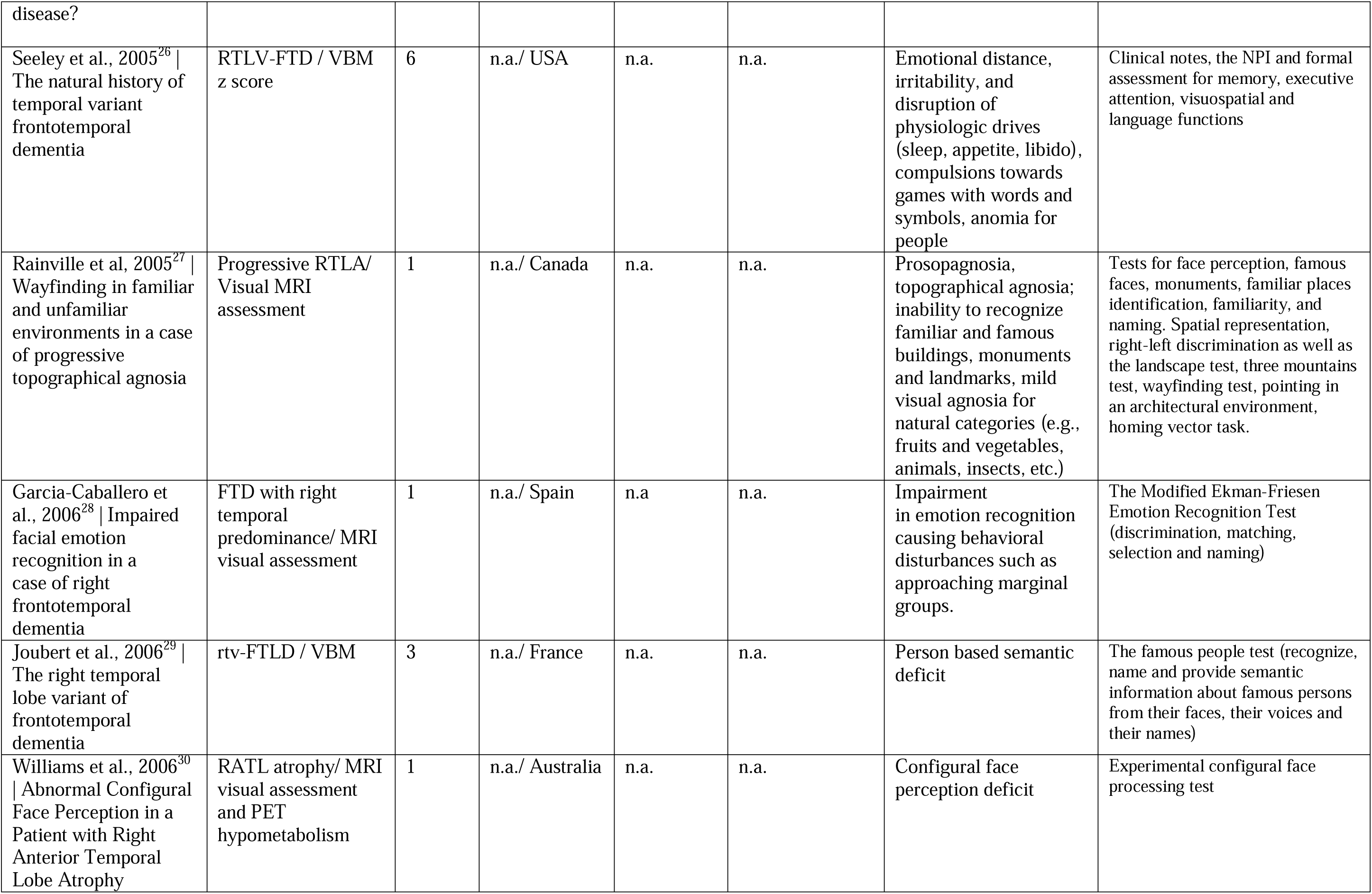
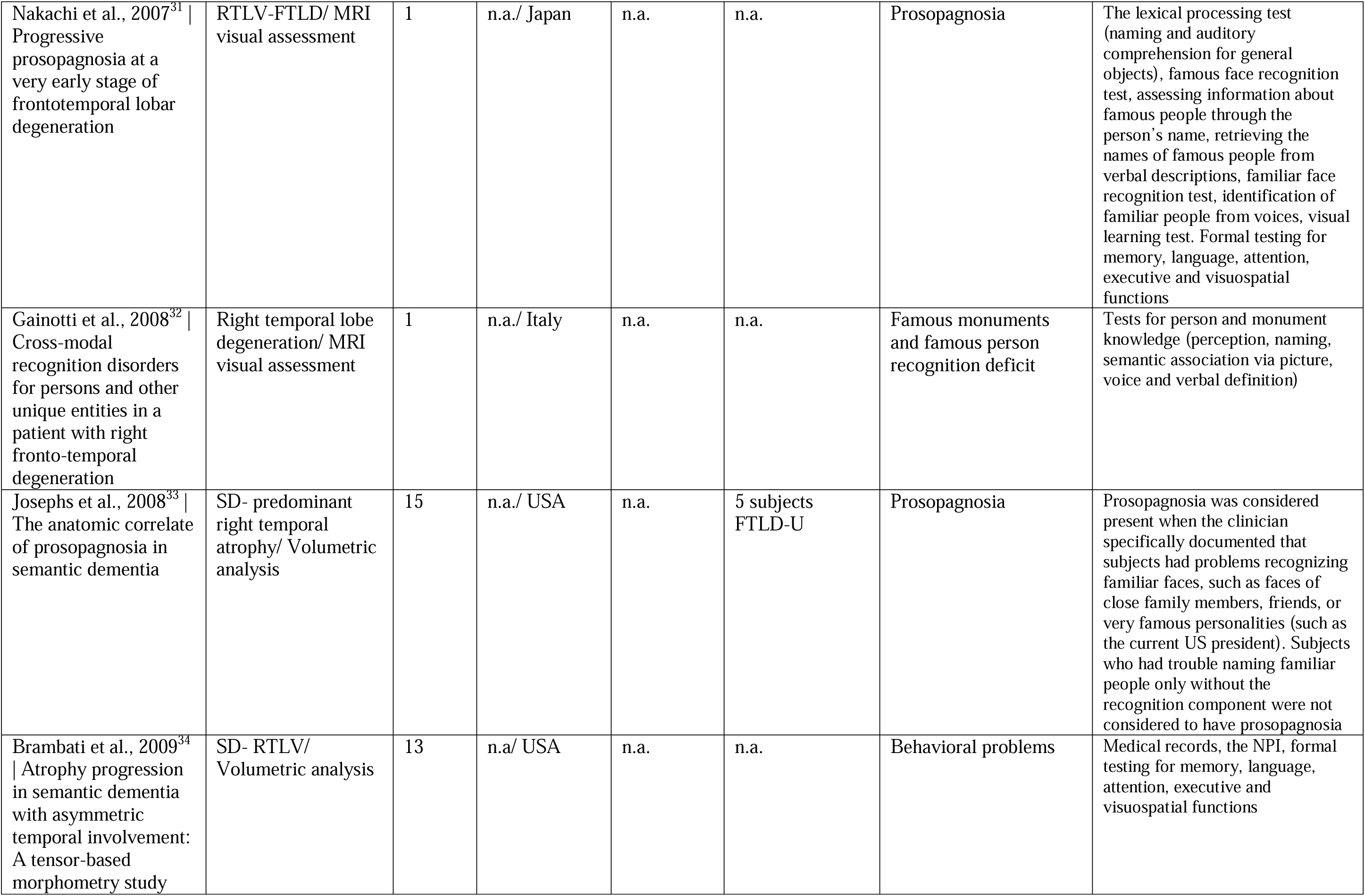
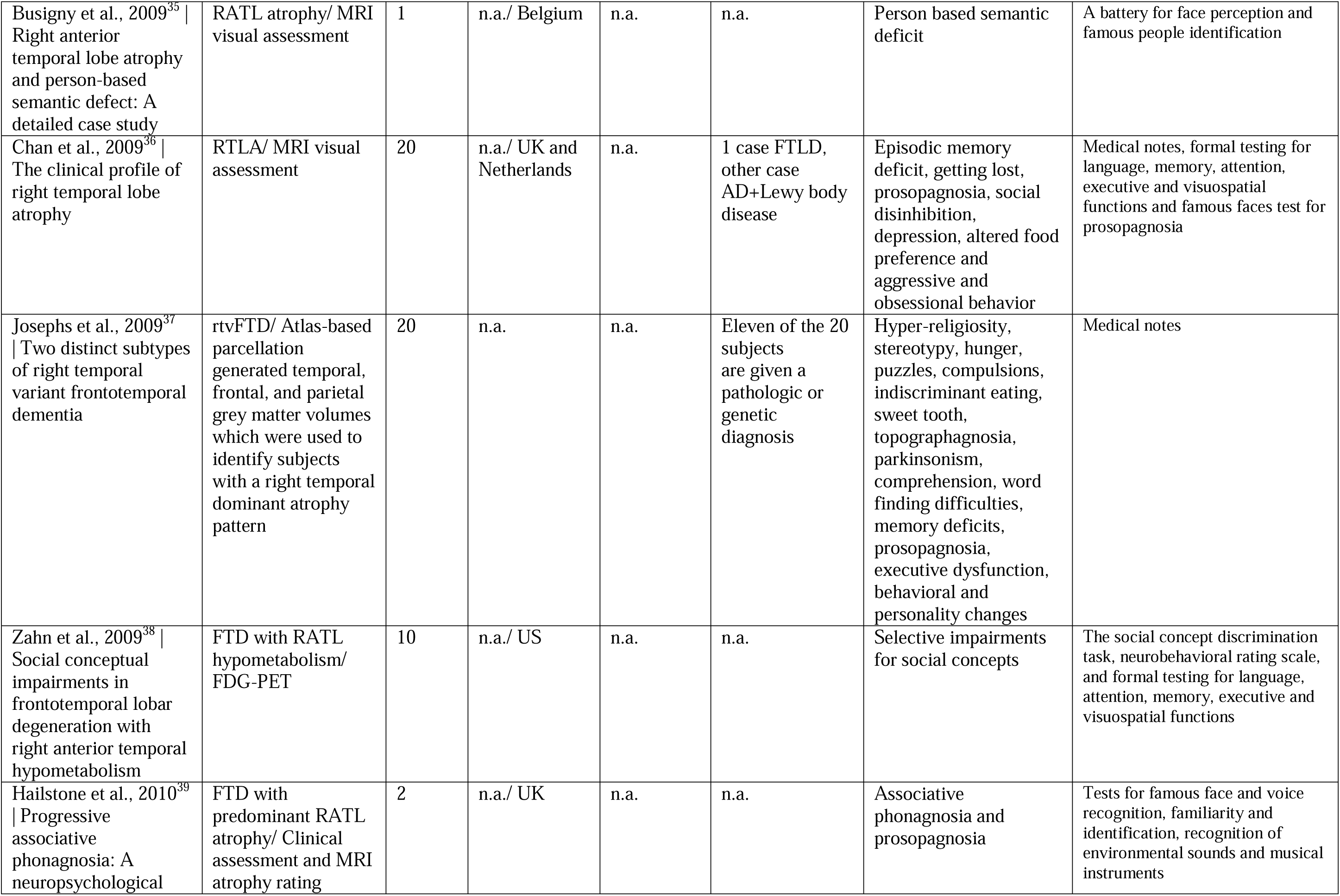
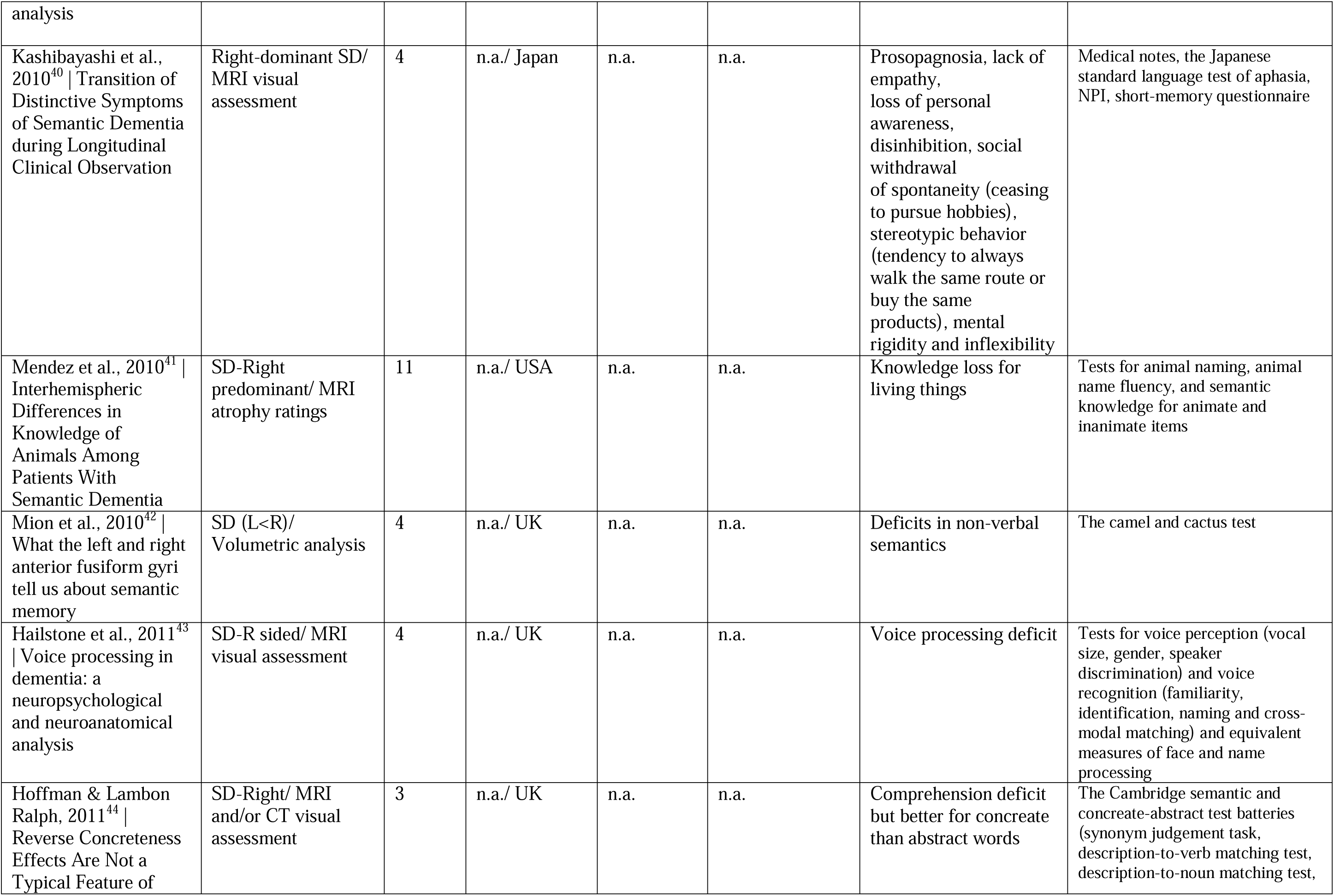
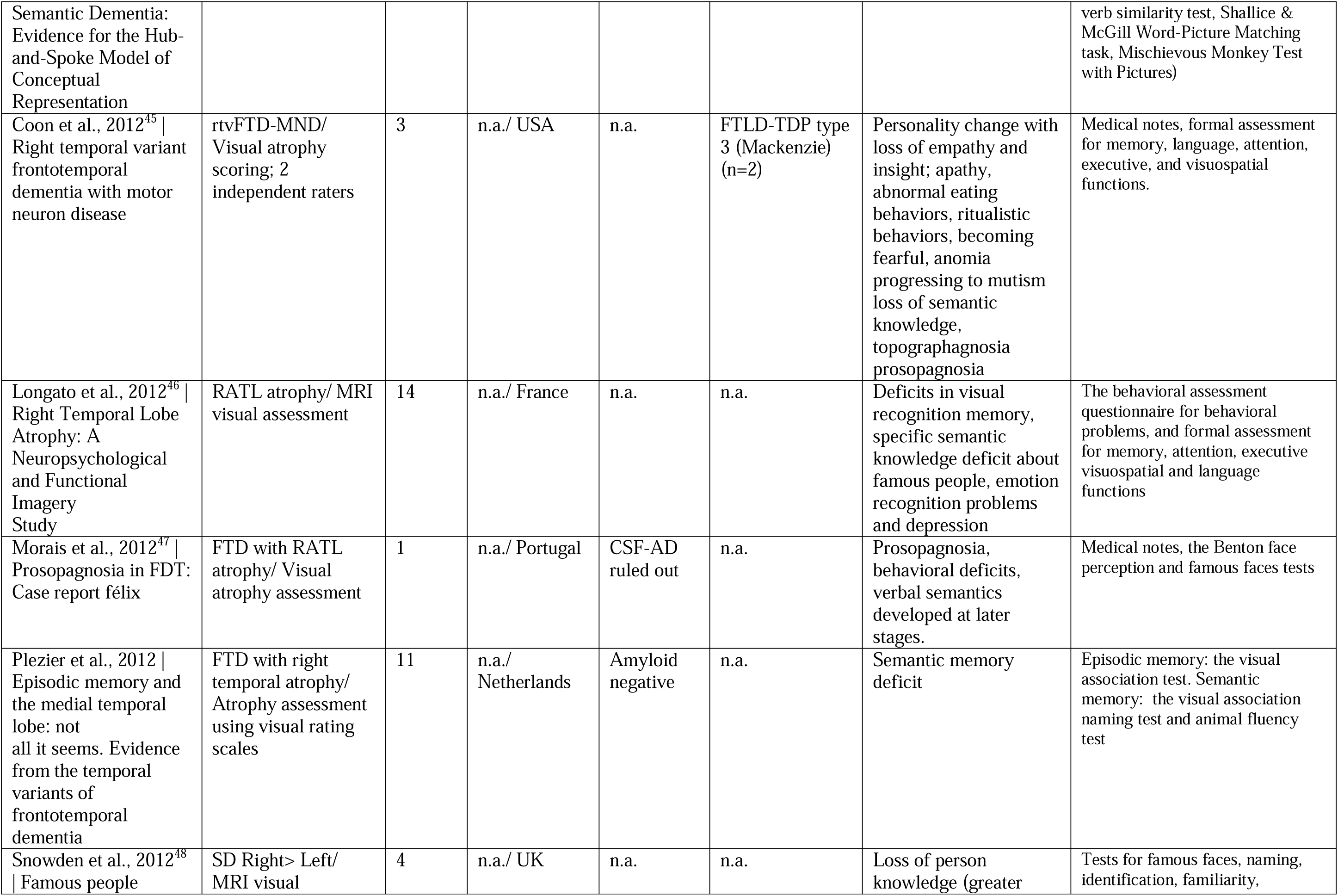
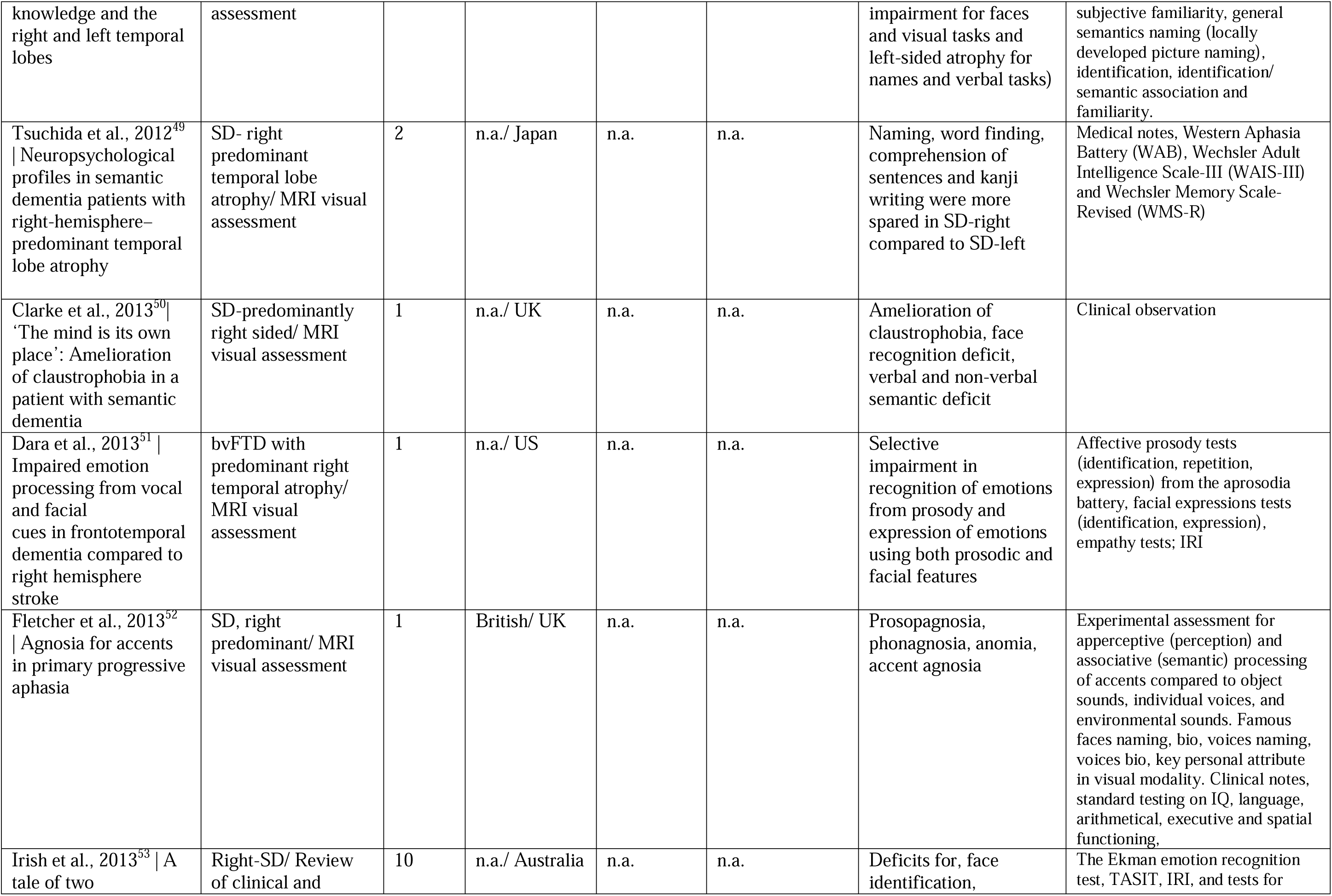
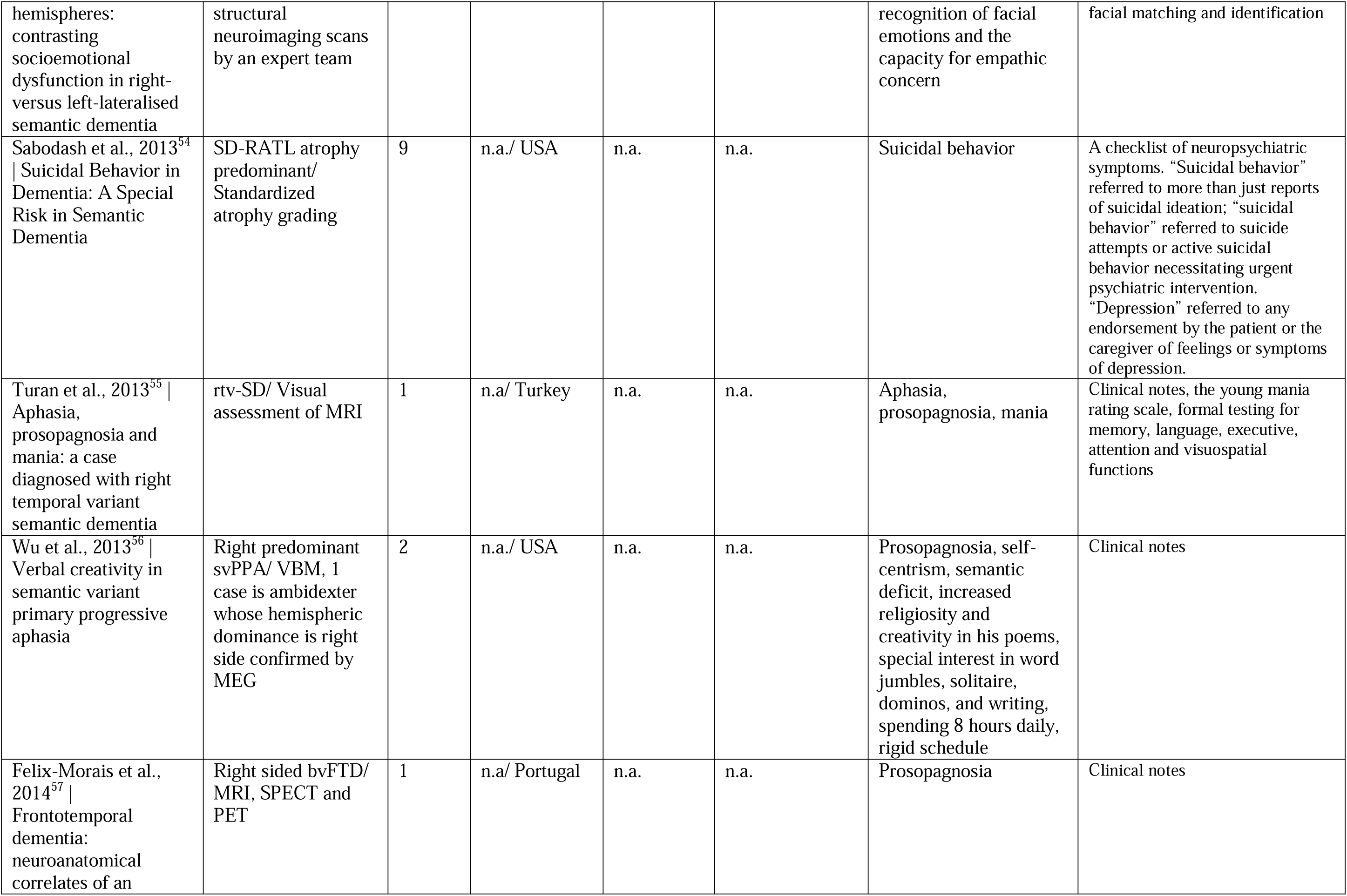
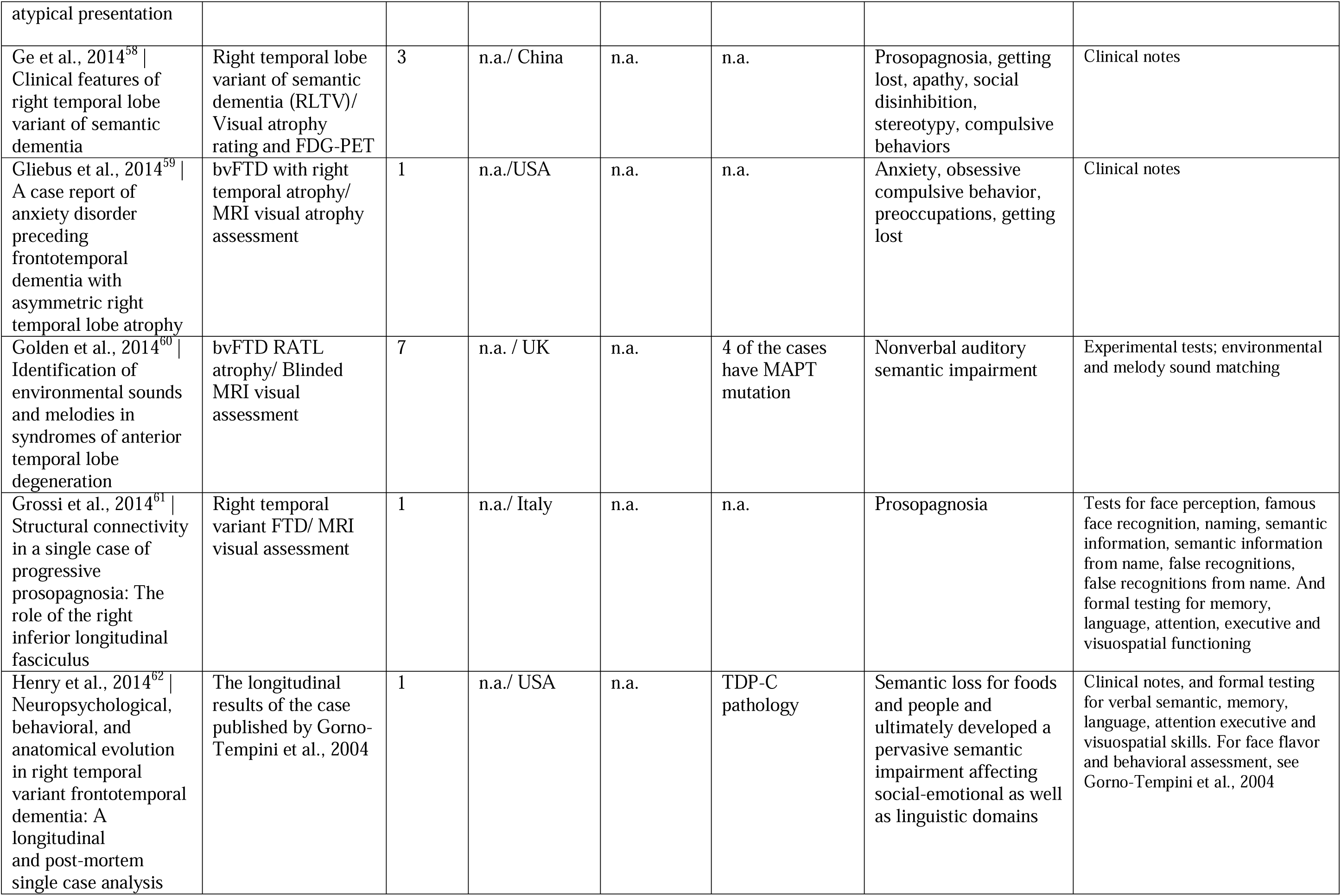
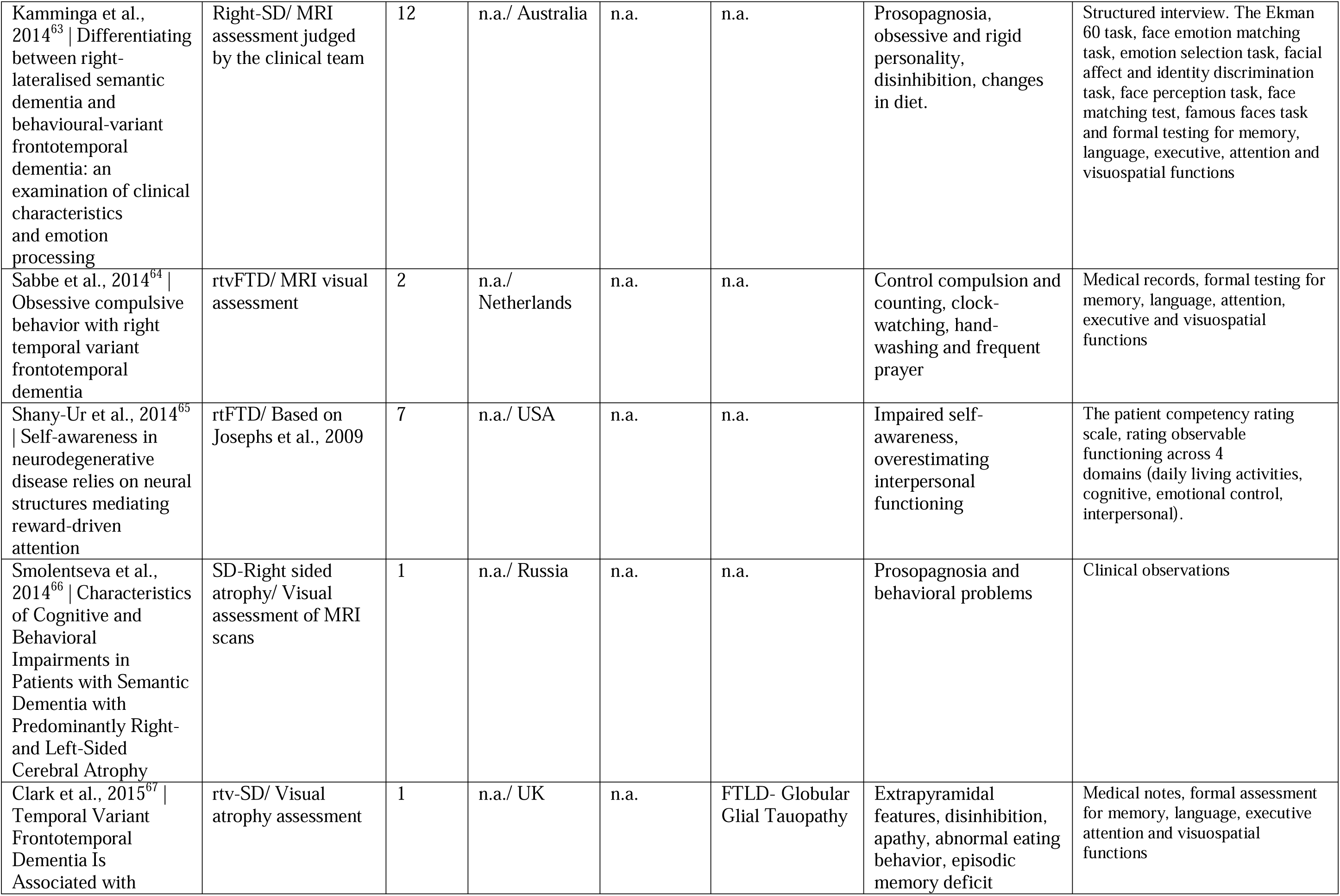
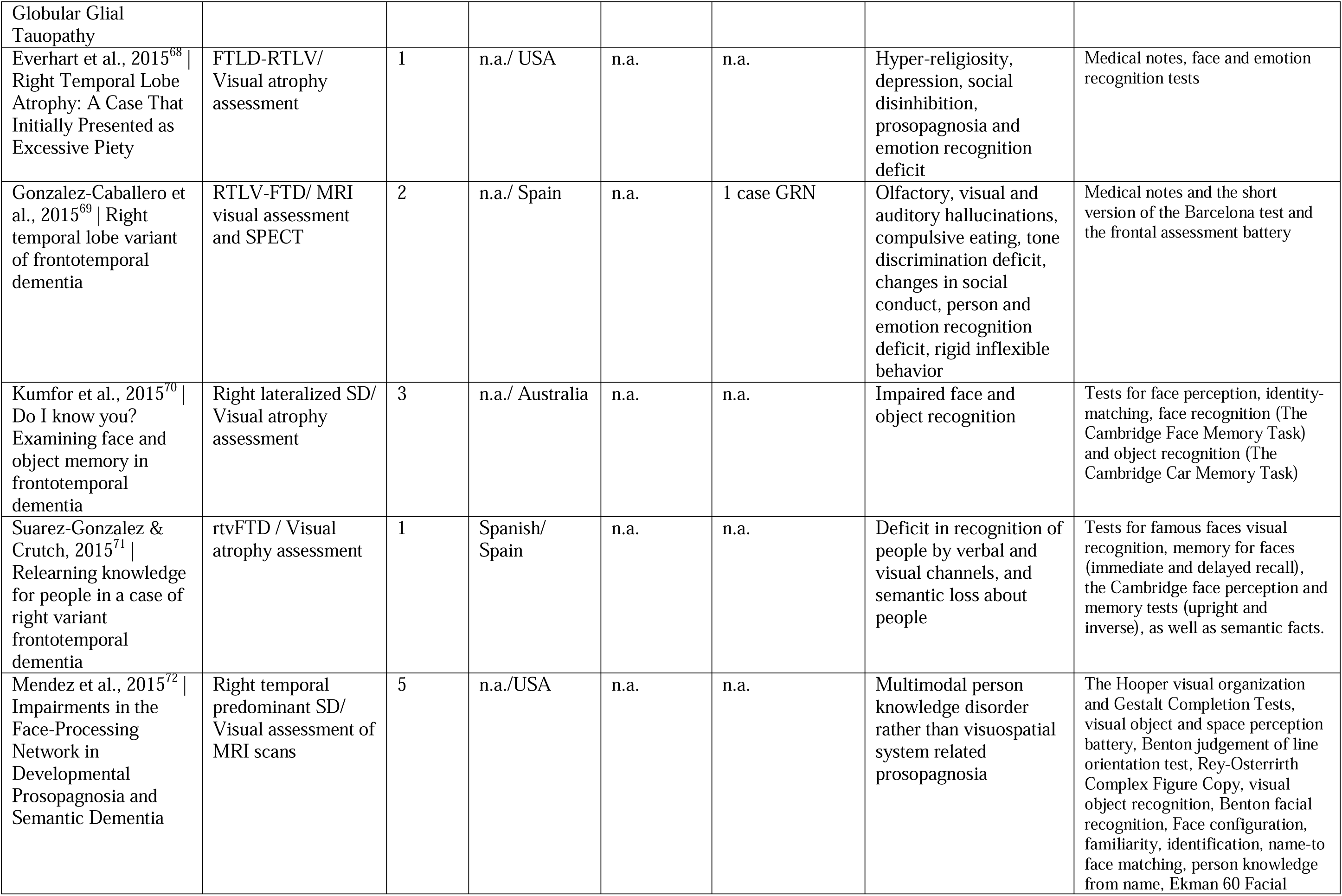
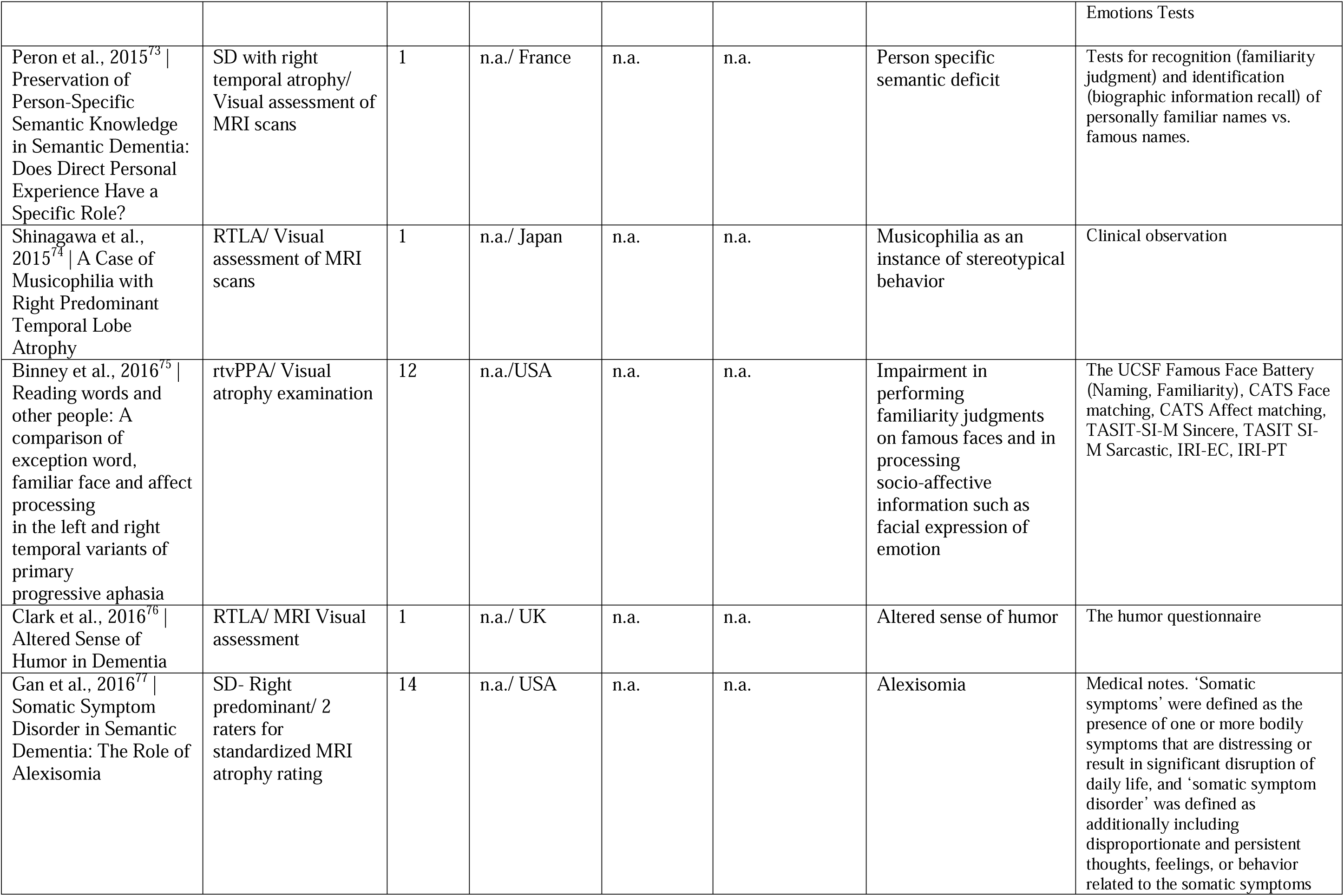
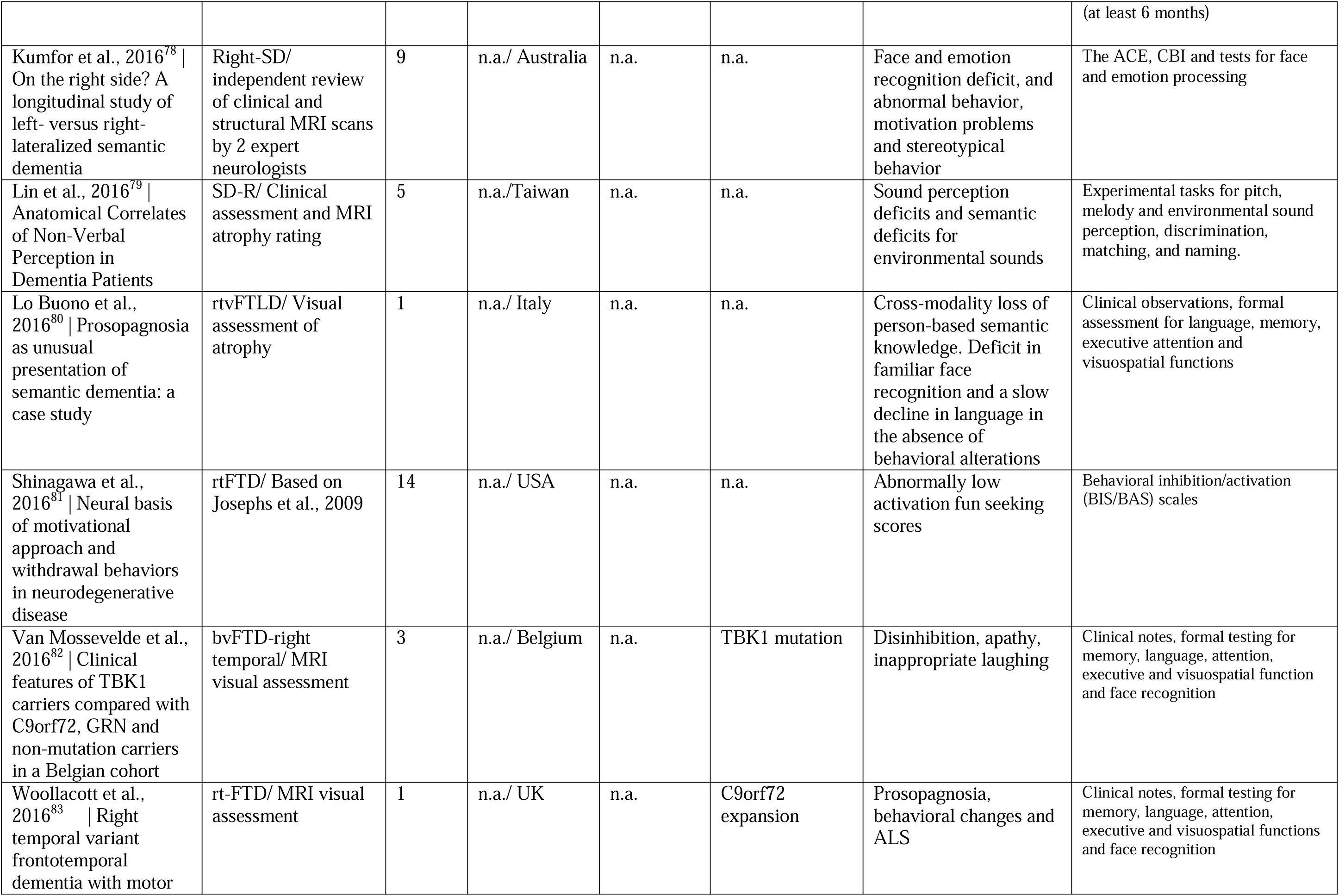
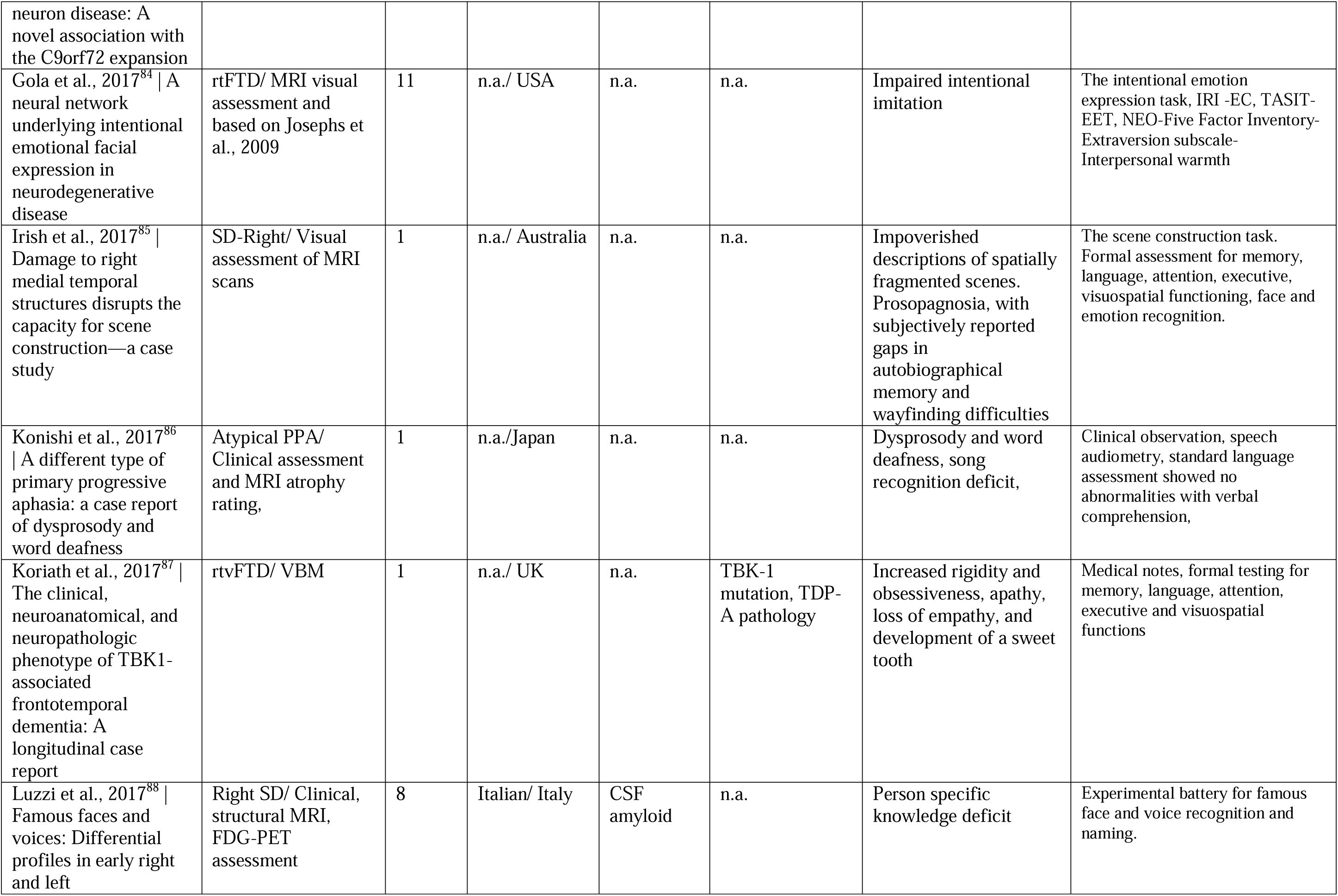
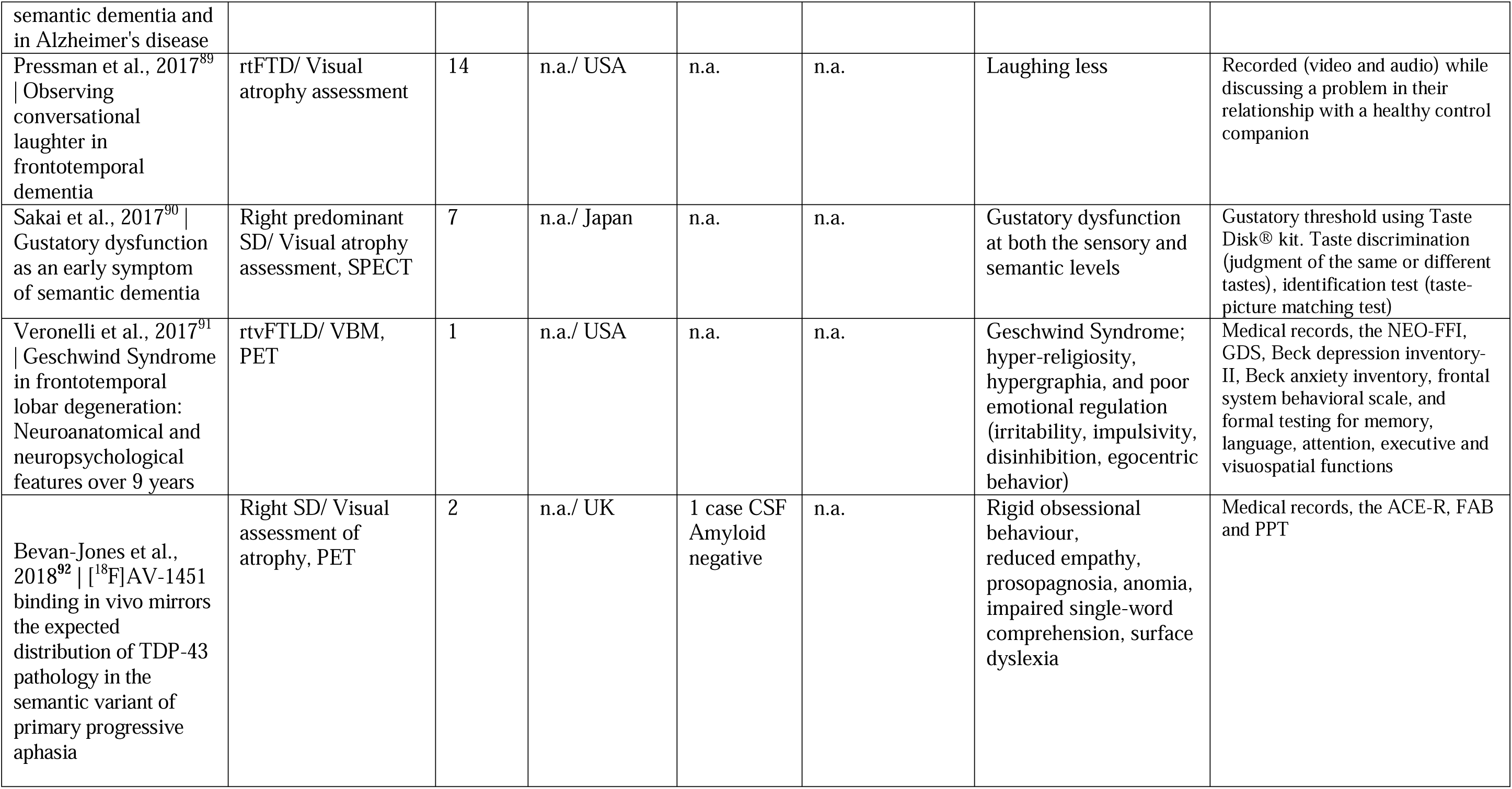
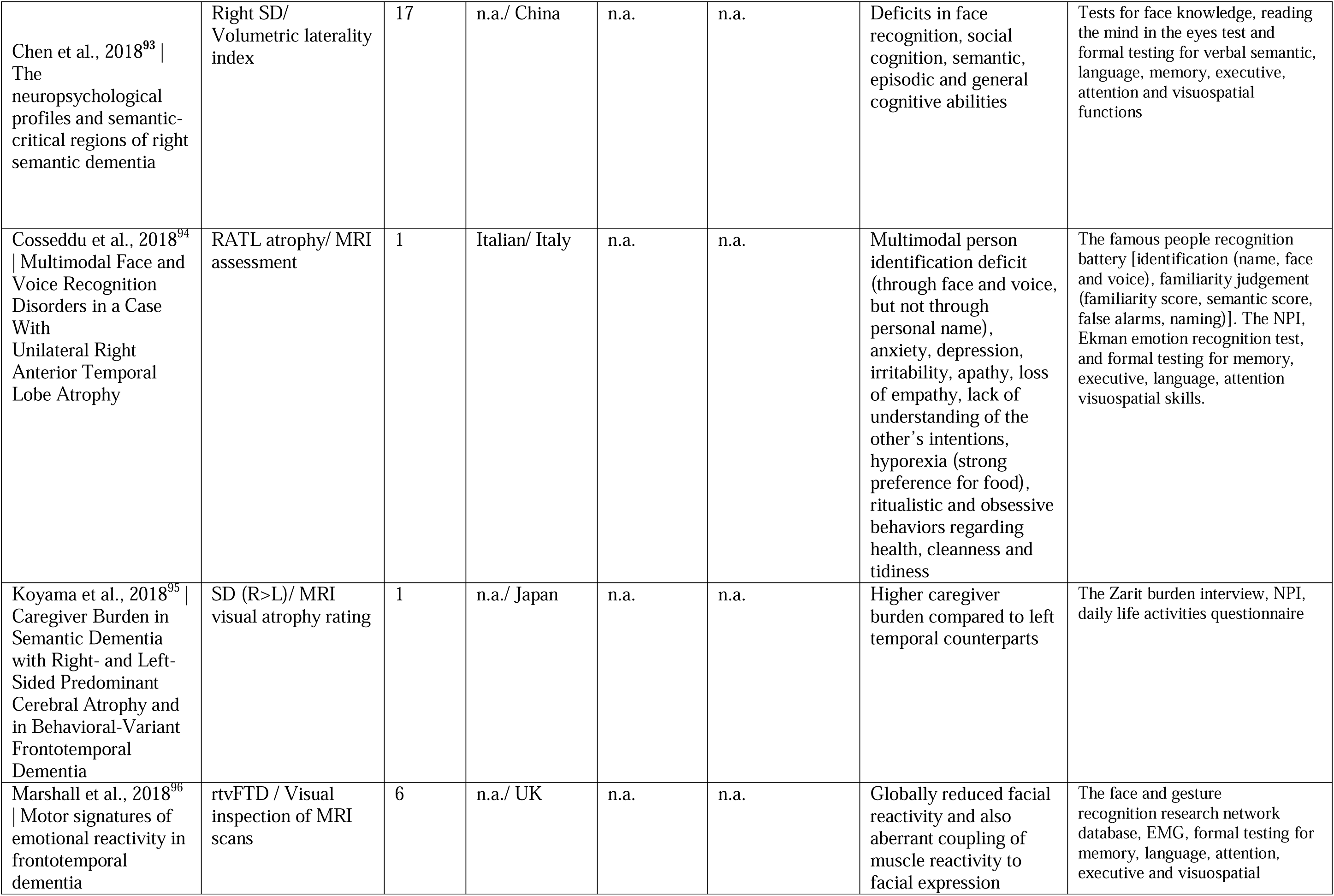
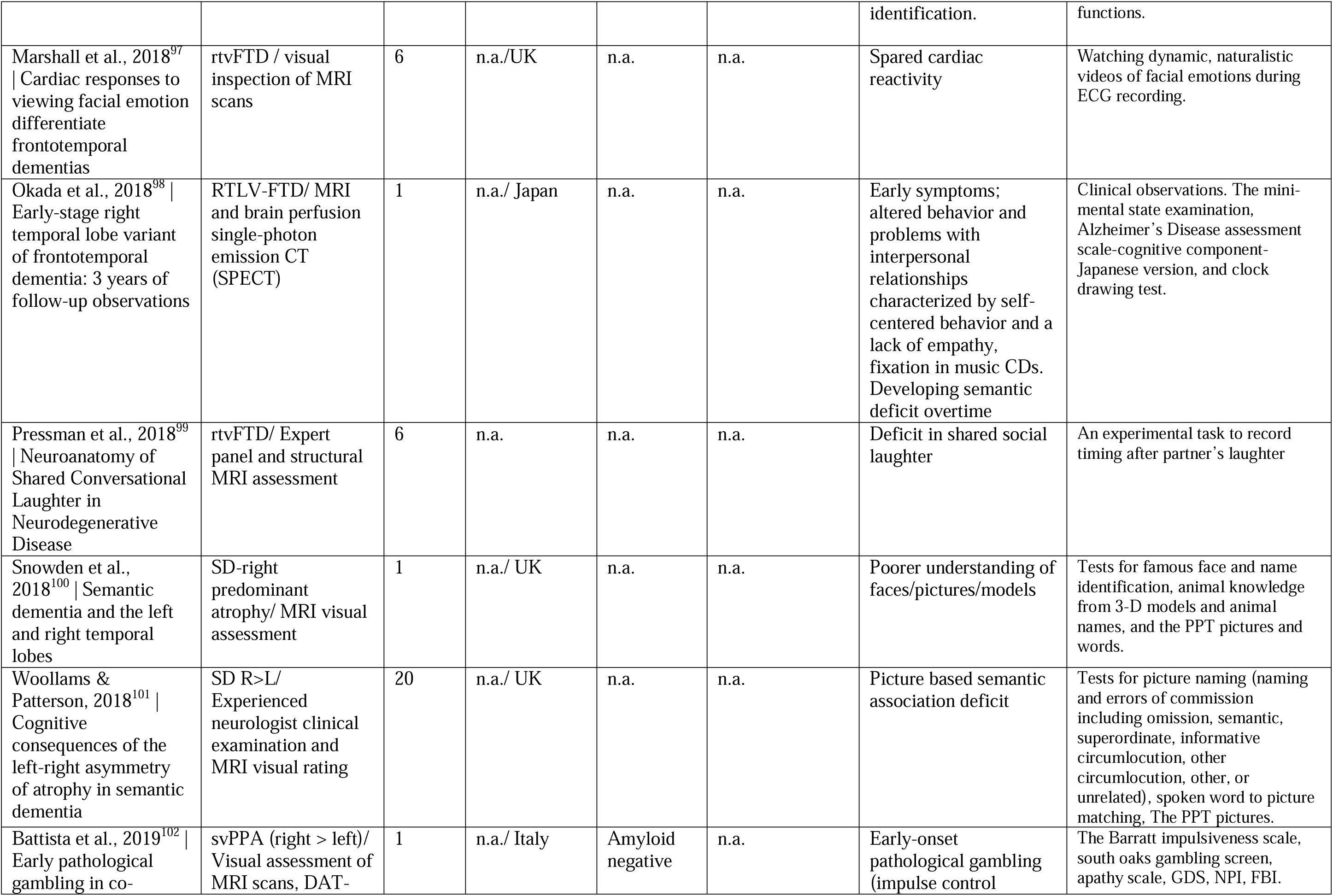
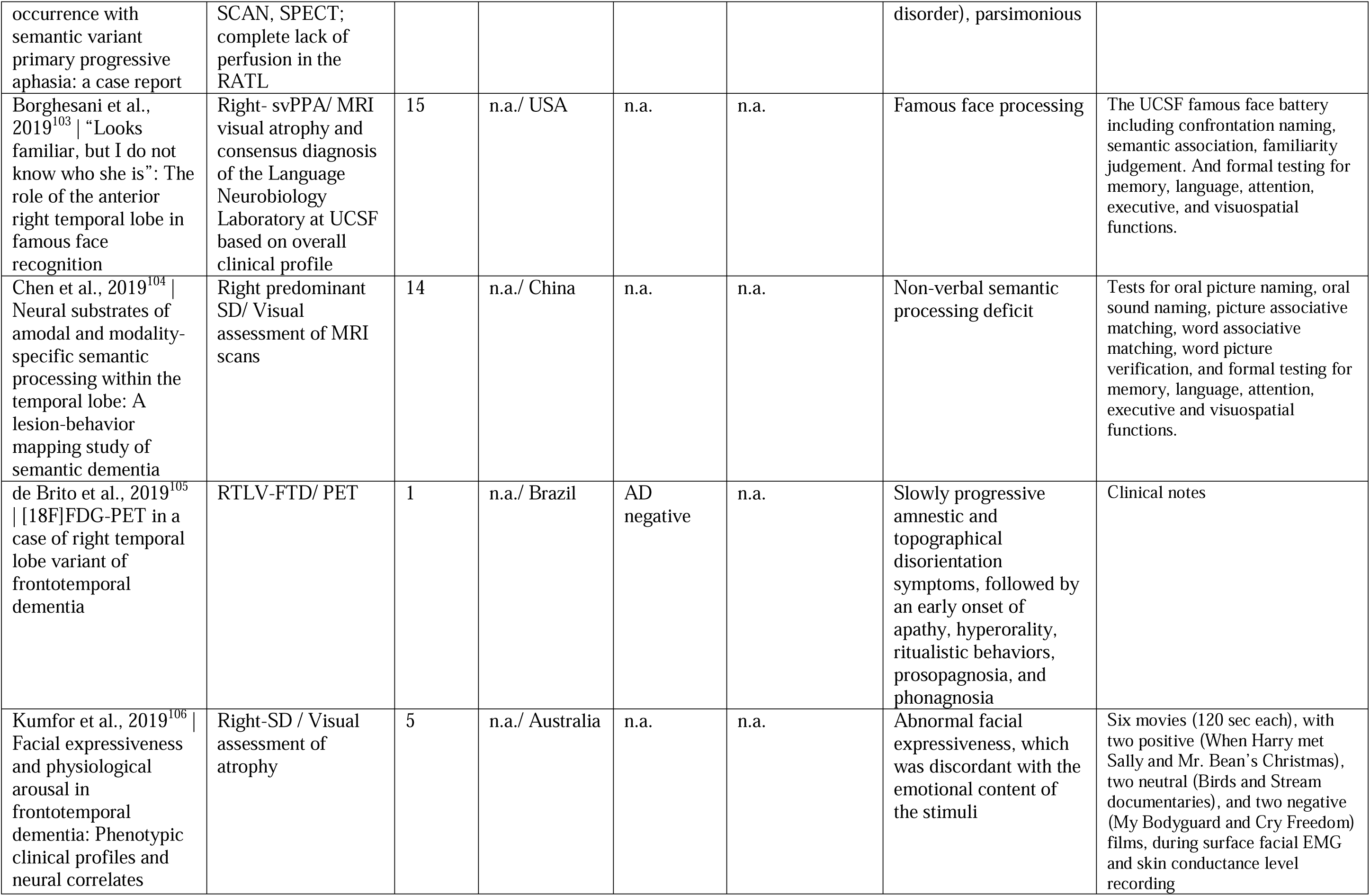
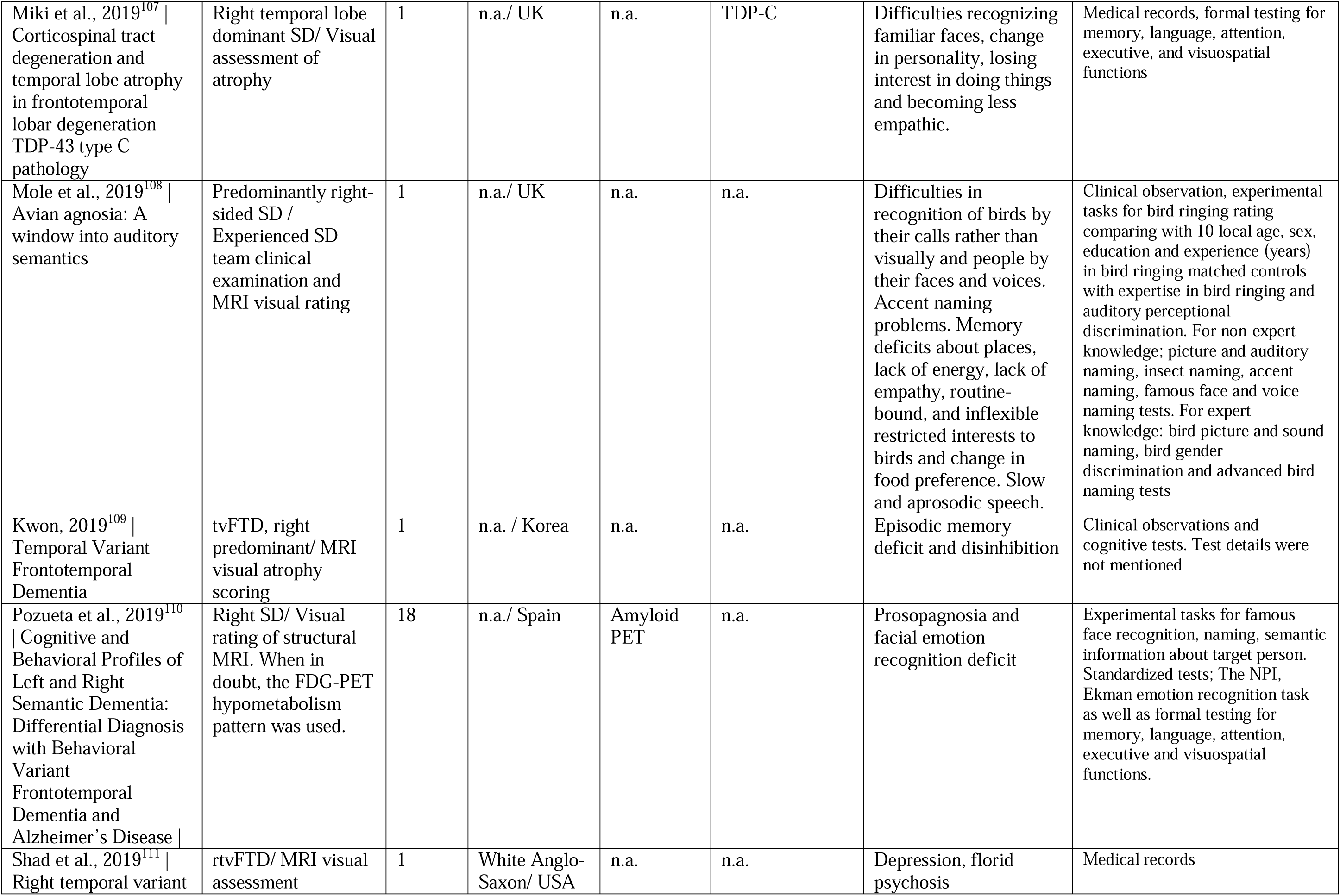
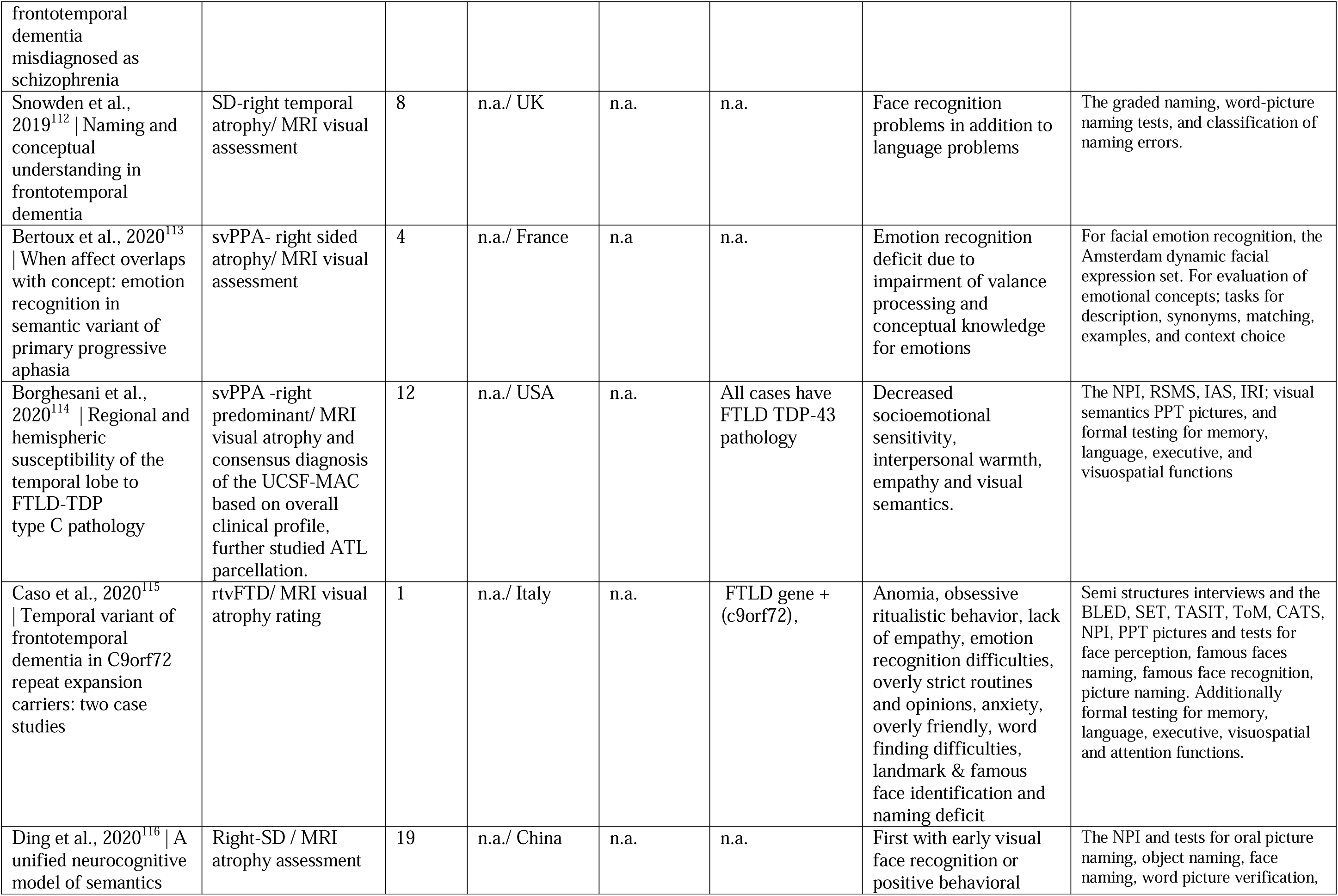
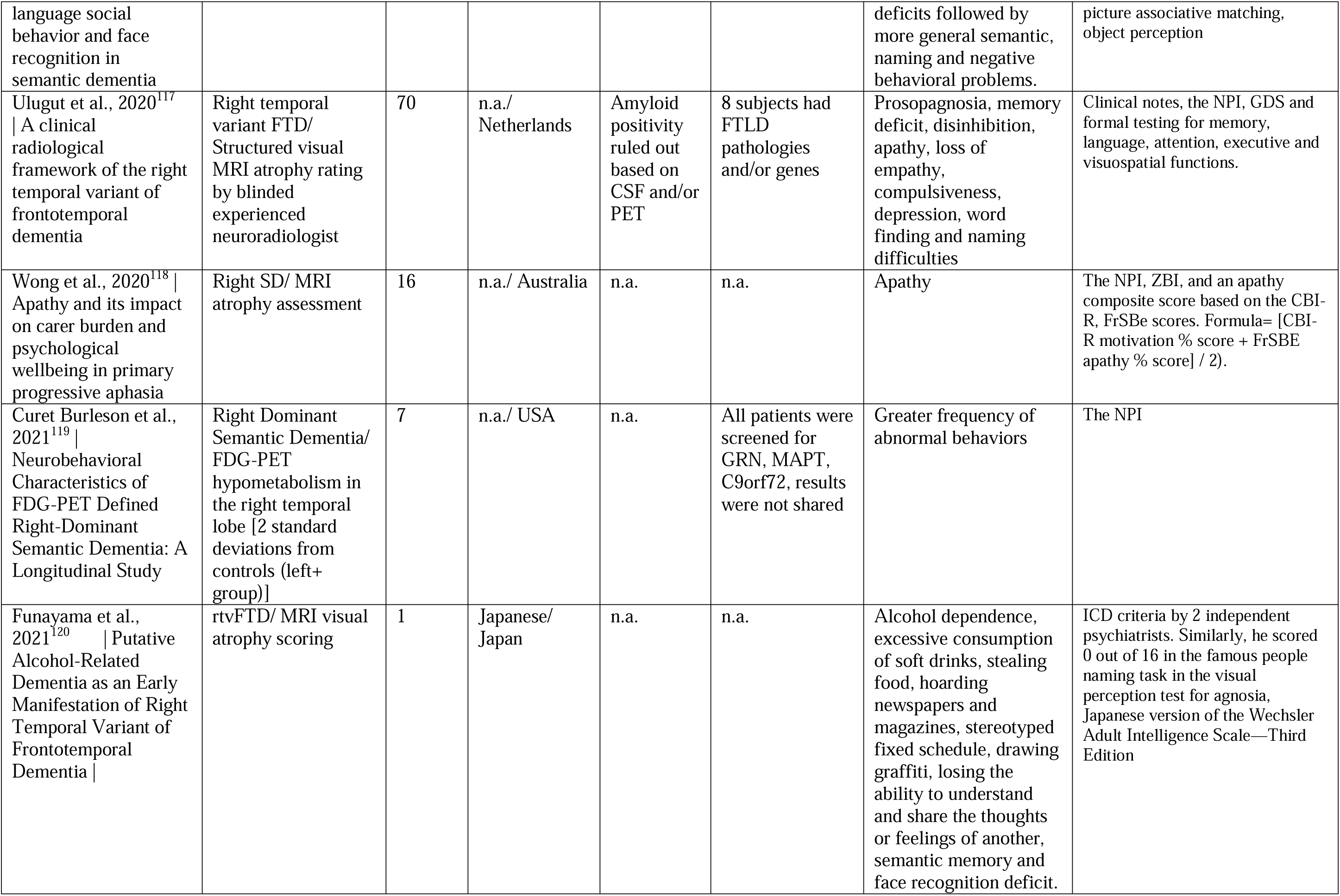
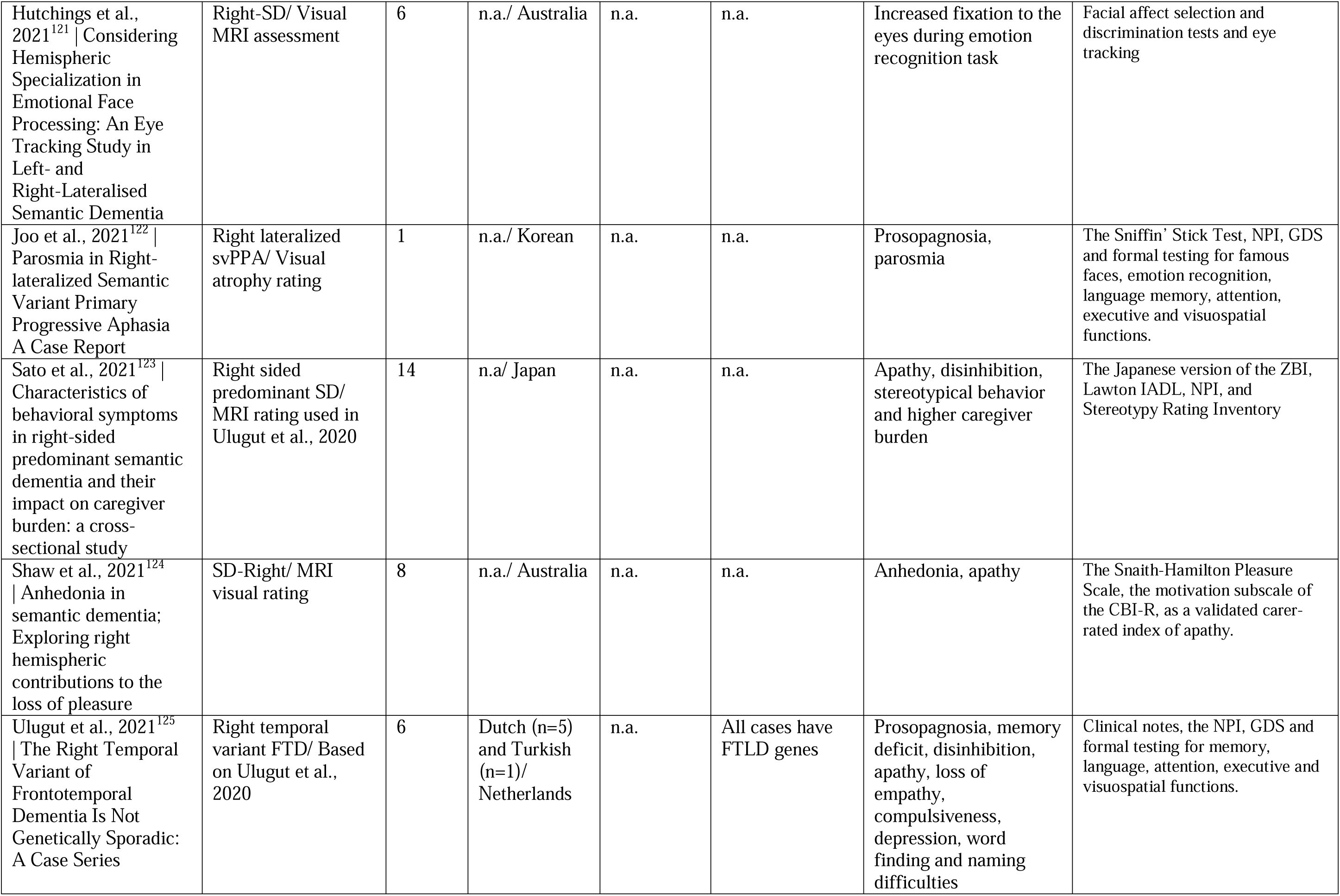
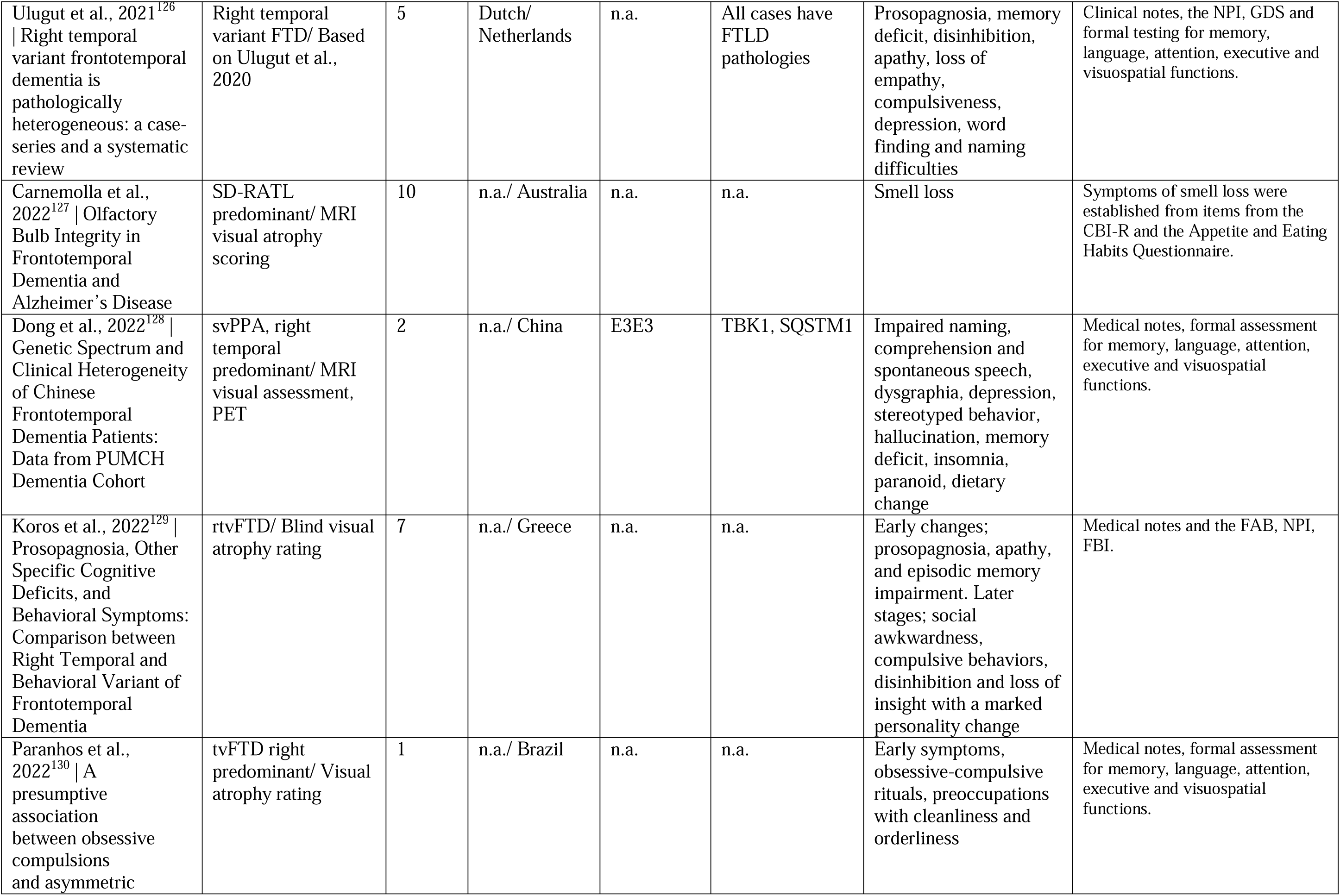
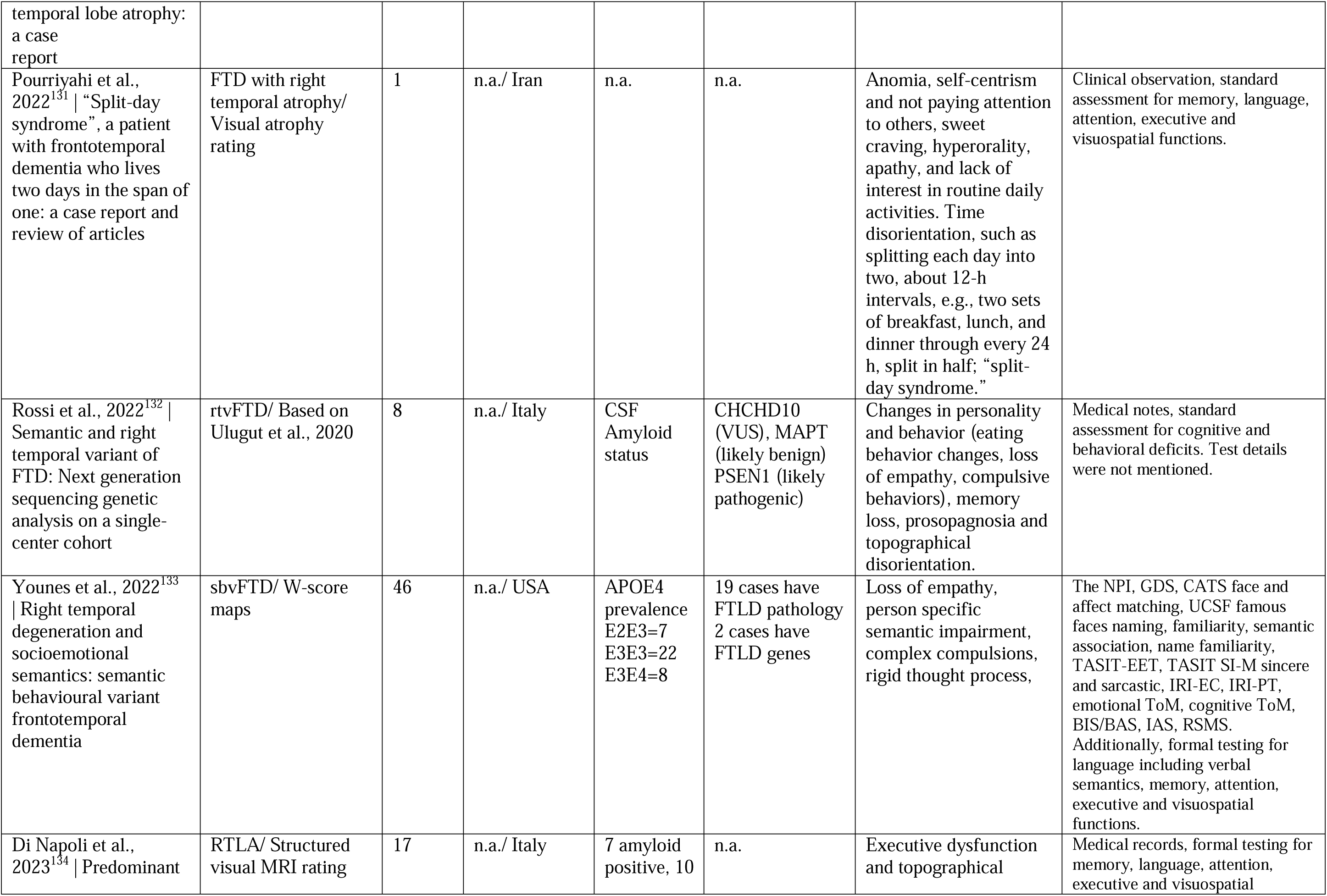
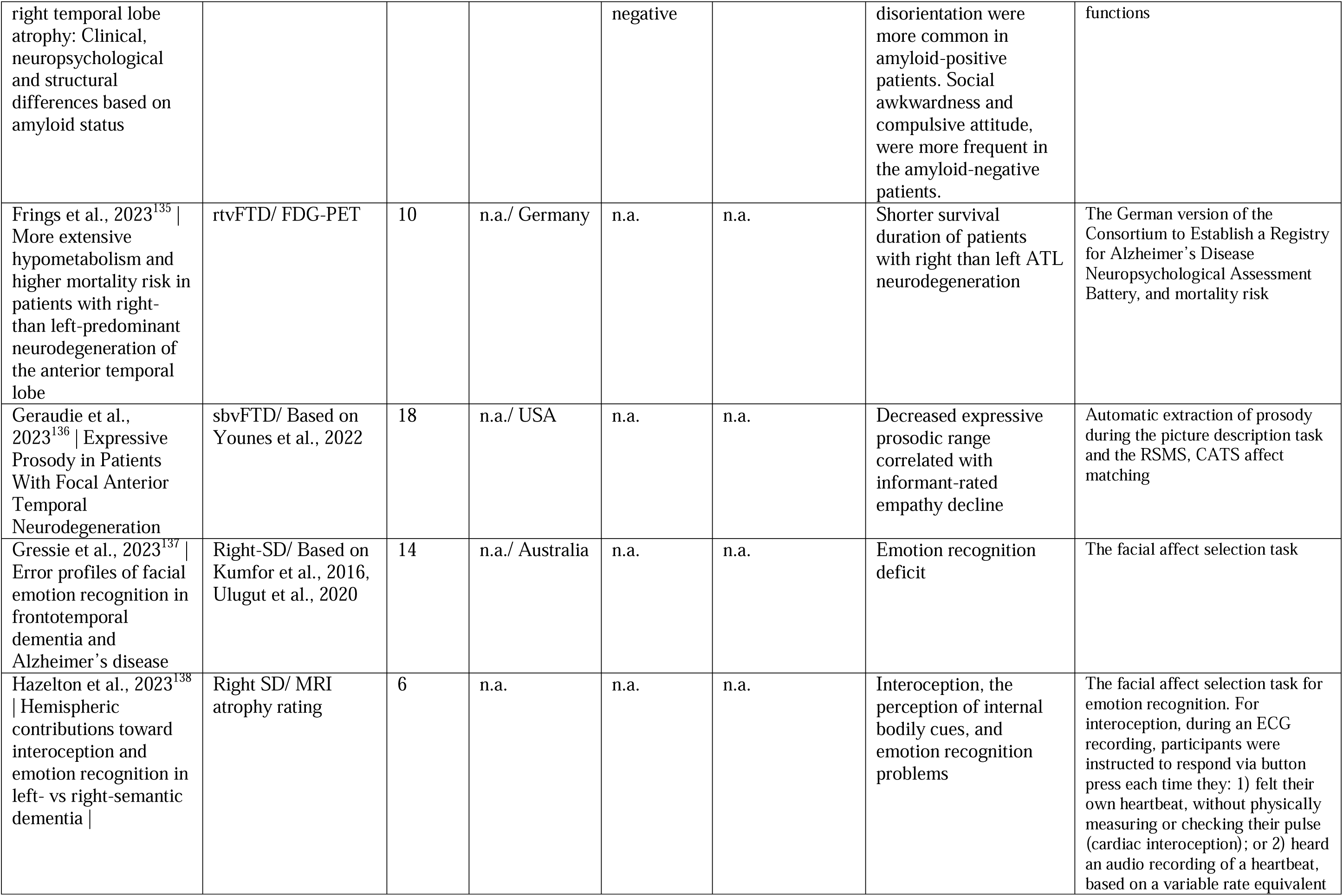
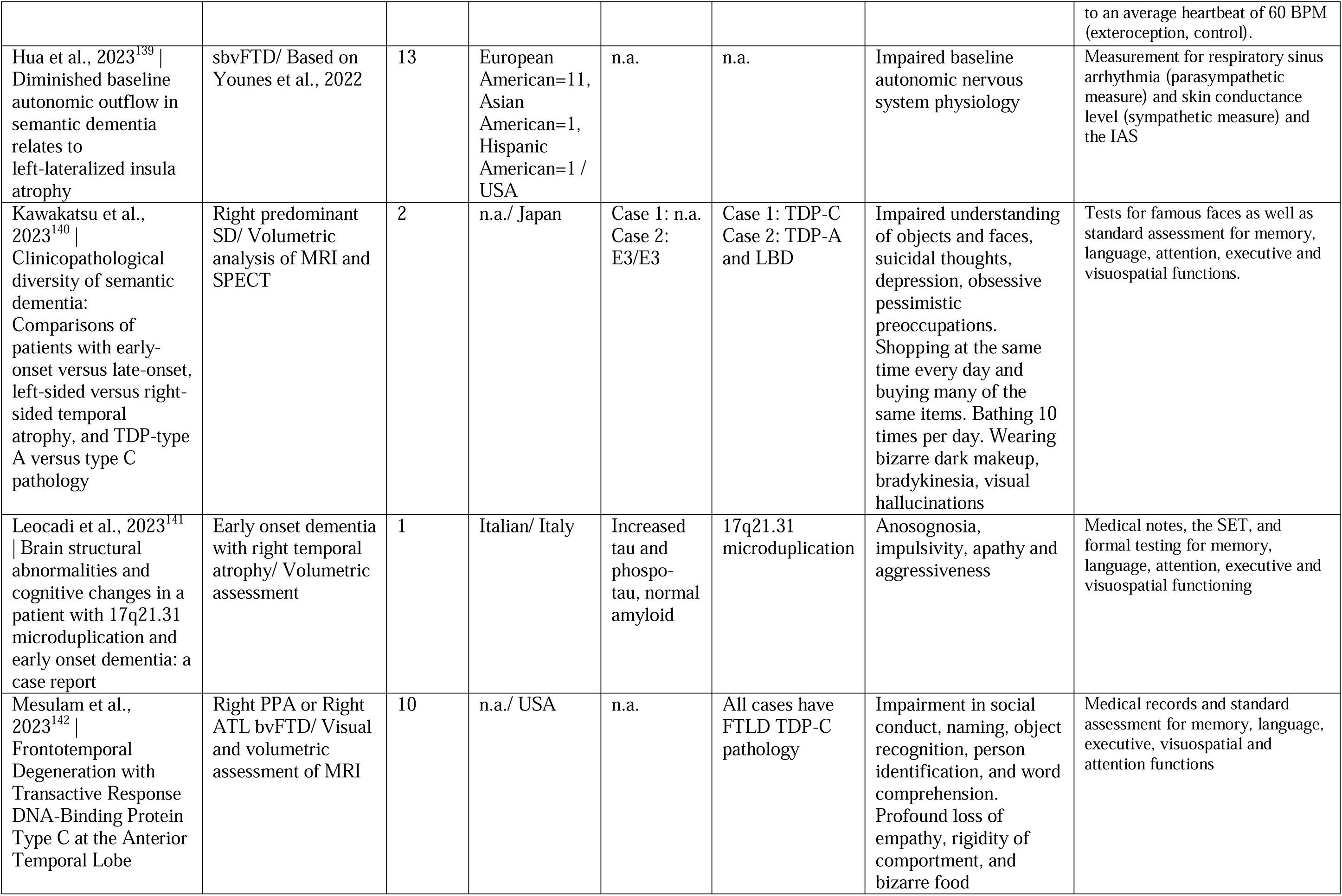
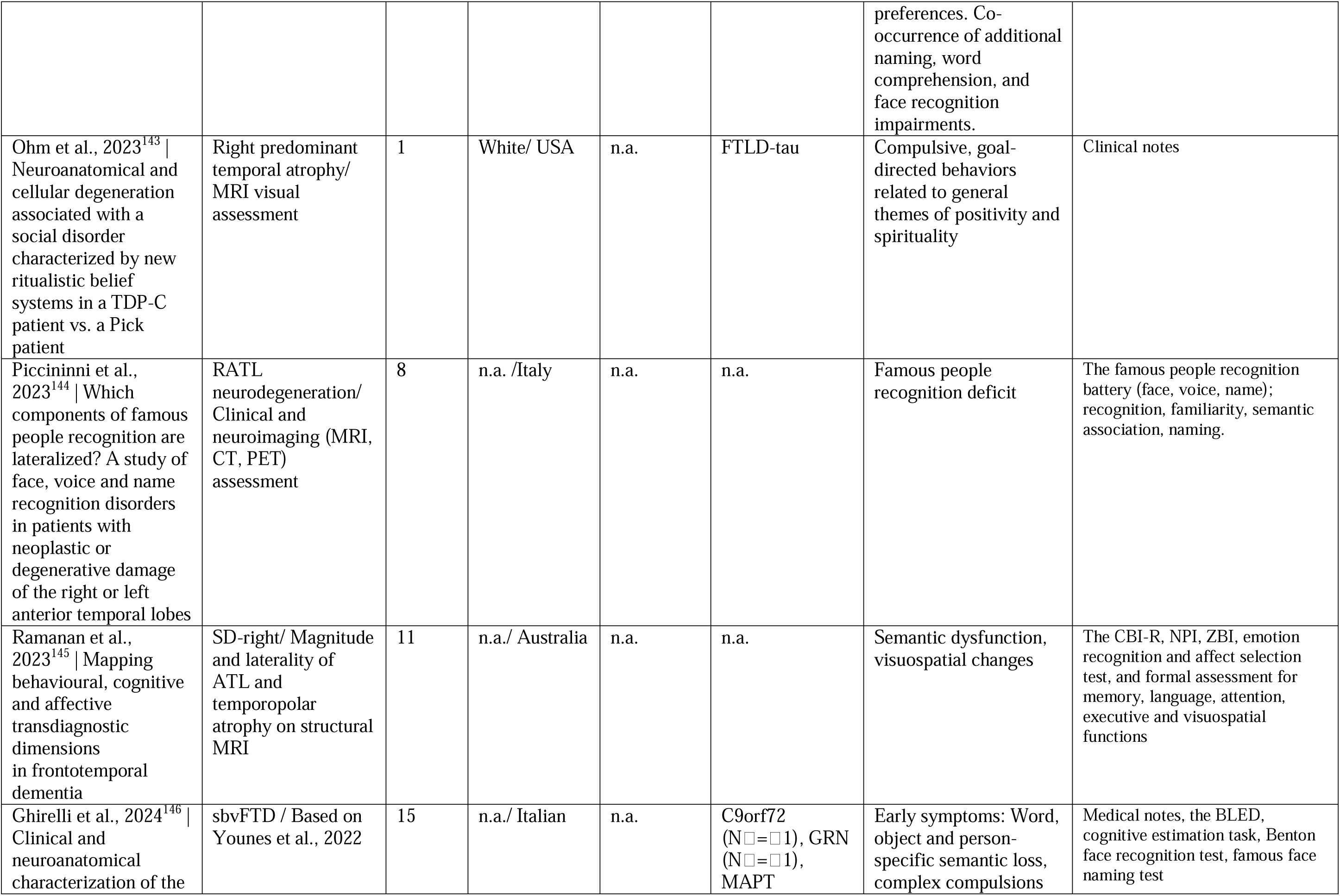
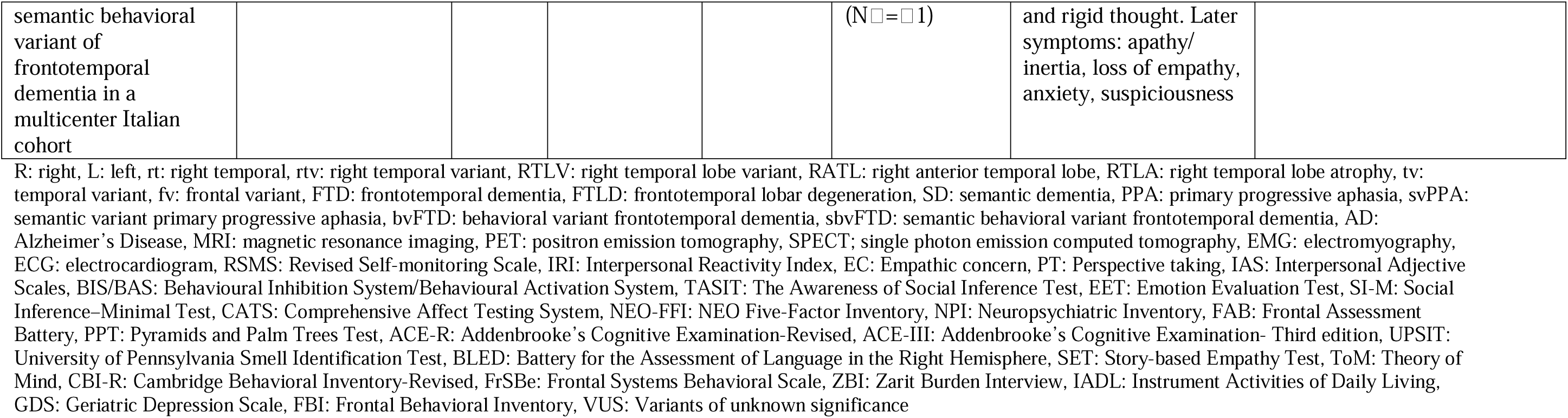
Included studies

## Notes

### Competing Interest Statement

The authors have declared no competing interest.

### Funding Statement

This project is supported by AACSF-22-849085 PI: Ulugut. Josephs and Whitwell are supported by R01-AG37491 R01-DC010367 R21-NS94684. Van den Stock is supported by KU Leuven (IDN/21/010 and C24/18/095) and the Sequoia Fund for research on ageing and mental health. Santillo is supported by the Swedish federal government under the ALF agreement. Bertoux is supported by the Fondation Planiol the Fondation France Alzheimer and Fondation Vaincre Alzheimer. Laforce; Chaire de recherche sur les aphasies primaires progressives Fondation de la Famille Lemaire. Warren is supported by the Alzheimer's Society Alzheimer's Research UK the Royal National Institute for Deaf People the NIHR UCLH Biomedical Research Centre and a Frontotemporal Dementia Research Fellowship in Memory of David Blechner (funded through The National Brain Appeal). Rankin is supported by RF1-AG029577. Rowe is funded by the Medical Research Council (MC_UU_00030/14; MR/T033371/1); the NIHR Cambridge Biomedical Research Centre (NIHR203312) and Wellcome Trust (220258). Vandenberghe receives support from the Mady Browaeys Fonds voor Onderzoek naar Frontotemporale Degeneratie.

### Author Declarations

Ethics committee/IRB od UCSF waived ethical approval for this work

