## Supplemental Material for "Clinical Recognition of Frontotemporal Dementia with Right Temporal Predominance; Consensus Recommendations of the International Working Group"

**SUPPLEMENTARY MATERIALS**

**Contents**

1. Member list of the international working group

*Page 2*

2. Establishment of the international working group

*Page 7*

3. Consensus approach

*Page 7*

4. Search terms for the systematic review

*Page 8*

5. Search strategy

*Page 9*

6. Atrophy rating guidelines

*Page 10*

7. Search results

*Page 16* **Member list of the international working group (IWG) (Alphabetical order)**

1. Augustin Ibanez, Latin American Brain Health Institute (BrainLat)
2. Alan Lerner, Case Western Reserve University
3. Alexander F Santillo, Lund University
4. Alexandre Morin, Rouen University Hospital
5. Alma Ghirelli, San Raffaele Scientific Institute
6. Andrea Arighi, Fondazione Hospital
7. Arabella Bouzigues, ICM Paris Brain Institude
8. Bedia Samanci, Istanbul University
9. Bradley Boeve, Mayo Clinic Rochester
10. Bruce Miller, UCSF Memory and Aging Center
11. Carmela Tartaglia, University of Toronto
12. Caroline Dallaire Theroux, University of Laval
13. Christopher Kobylecki, Manchester Cerebral Function Unit
14. Daniel Ohm, Penn University
15. Daniela Galimberti, University of Milan
16. David Foxe, University of Sydney
17. David Irwin, Penn University
18. David Perry, UCSF Memory and Aging Center
19. Diana Matallana, Javeriana University
20. Edoardo Gioele Spinelli, San Raffaele Scientific Institute
21. Elisa Canu, San Raffaele Scientific Institute
22. Elisabet Englund, Lund University
23. Emily Rogalski, Northwestern University
24. Emma Devenney, University of Sydney
25. Emma Rhodes, Penn University
26. Eun Joo Kim, Pusan National University
27. Federica Agosta, San Raffaele Scientific Institute
28. Francesco Di Lorenzo, Foundation Santa Lucia
29. Frederik Barkhof, Alzheimer Center Amsterdam
30. Fiona Kumfor, University of Sydney
31. Florence Pasquier, University of Lille
32. Floor Duits, Alzheimer Center Amsterdam
33. Gail Robinson, The University of Queensland
34. Giorgio Fumagalli, University of Milan
35. Giuseppe Piga, ICM Paris Brain Institute
36. Gregory Kuchcinski, University of Lille
37. Hakan Gurvit, Istanbul University
38. Halle Quang, University of Sydney
39. Harro Seelaar, Erasmus University
40. Howie Rosen, UCSF Memory and Aging Center
41. Hulya Ulugut, UCSF Memory and Aging Center
42. Ignacio Illan Gala, Hospital de la Santa Creu I Sant Pau
43. James Rowe, Cambridge University
44. Jan Van den Stock, Leuven University
45. Janine Diehl Schmid, Technical University of Munich
46. Jason Warren, Dementia Research Centre London
47. Jennifer Thompson, Manchester Cerebral Function Unit
48. Jennifer Whitwell, Mayo Clinic Rochester
49. Jessica Hazelton, University of Sydney
50. Jonathan Rohrer, Dementia Research Centre London
51. Julie Fields, Mayo Clinic Rochester
52. Julie Snowden, Manchester Cerebral Function Unit
53. Julien Lagarde, Hospital Sainte Anne
54. Jwala Narayanan, Manipal Hospital India
55. Katherine Rankin, UCSF Memory and Aging Center
56. Katya Rascovsky, Penn University
57. Keith Josephs, Mayo Clinic Rochester
58. Kristina Horne, University of Sydney
59. Kyan Younes, Stanford University
60. Leonardo Cruz de Souza, Universidade Federal de Minas Gerais
61. Leonel T. Takada, University of Sao Paulo
62. Lina Riedl, Technical University of Munich
63. Lize Jiskoot, Erasmus University
64. Lucy Russell, Dementia Research Centre London
65. Manuela Pintus, Fondazione Hospital
66. Maria Landqvist Waldo, Lund University
67. Maria Luisa Gorno Tempini, UCSF Memory and Aging Center
68. Mario F. Mendez, University of California Los Angeles
69. Marsel Mesulam, Northwestern University
70. Massimo Filippi,  San Raffaele Scientific Institute
71. Masud Husain, Oxford University
72. Mathieu Vandenbulcke, Leuven University
73. Matthew Jones, Manchester Cerebral Function Unit
74. Matthew Rouse, Cambridge University
75. Matthias L. Schroeter, Max Planck Institute
76. Maud Tastevin, University of Laval
77. Maxime Bertoux, University of Lille
78. Maxime Montembeault, McGill University
79. Mira Didic, Aix-Marseille University
80. Murat Emre, Istanbul University
81. Muireann Irish, University Sydney
82. Na-Yeon Jung, Pusan National University
83. Olivier Piguet, University of Sydney
84. Oskar Hansson, Lund University
85. Philip Scheltens, Alzheimer Center Amsterdam
86. Paulo Caramelli, Universidade Federal de Minas Gerais
87. Peter Pressman, University of Colorado
88. Raffaella Migliaccio, Pitie-Salpetriere Hospital, INSERM
89. Ratnavallie Ellajosyula, Manipal Hospital India
90. Rik Ossenkoppele, Alzheimer Center Amsterdam
91. Rik Vandenberghe, Leuven University
92. Robert Laforce, University of Laval
93. Samantha Loi, University of Melbourne
94. Sandra Weintraub, Northwestern University
95. Shalom Henderson, Cambridge University
96. Sian Thompson, Oxford University
97. Siddharth Ramanan, Cambridge University
98. Simon Ducharme, McGill University
99. Sun Min Lee, Ajou University
100. So Young Moon, Ajou University
101. Thibaud Lebouvier, University of Lille
102. Toji Miyagawa, Mayo Clinic Rochester
103. Virginia Sturm, UCSF Memory and Aging Center
104. Welmoed Krudop, Alzheimer Center Amsterdam
105. Wiesje van der Flier, Alzheimer Center Amsterdam
106. Yolande Pijnenburg, Alzheimer Center Amsterdam

**Establishment of the IWG:** HU and YP reviewed the international literature to identify the authors / centers that might have an interest in rtvFTD, based on their scientific reports and /or clinical cohort characteristics. Partners were invited via email and/or zoom meetings to collaborate in the project. Initial invitations were extended to multiple centers, and subsequent Zoom meetings were held to outline the objectives of the IWG and the specific project. This resulted in a positive reaction for collaboration from 18 centers including the United States, United Kingdom, Italy, France, Belgium, Germany, Spain, Sweden, Canada, Turkey, Brazil, and the Netherlands. The invitation remained open to allow additional centers to join subsequent round table meetings, which aimed to achieve consensus on various aspects including aims, methodology, study designs, generation of the symptom checklist for data collection, data analyses, interpretation of the collected data, terminology, recommendations, and future directions. Over time new collaborators have joined the collaboration that contributed to the round table discussions although they couldn’t provide patient data for the retrospective study. Currently the IWG contains 55 centers across the world with more than 100 early career and senior FTD researchers.

**Consensus approach:** A series of round table meetings was organized to reach consensus on aims, methodology, generation of the symptom checklist, study designs and future directions. The nominal group process technique which combines qualitative and quantitative data and encourages and enhances the participation of group members was used to deliver rapid and reliable results to have a consensus and generate ideas/solutions to be used in the project (*GALLAGHER M, HARES T, SPENCER J, BRADSHAW C, WEBB I. The Nominal Group Technique: A Research Tool for General Practice? Fam Pract. 1993 Mar 1;10(1):76–81*). All meetings were recorded and shared on safe platforms (SURFDrive). Generation of the symptom checklist, symptom definitions, interpretations, terminology, and how to abstract data from case notes were elucidated and a guidebook, open discussion chat (SURFDrive), and a dataset template were prepared based on the discussions. Following data collection, preliminary results such as sample characteristics, frequencies of the symptoms and cognitive test results were documented, and surveys were prepared to systematically collect the opinions of the collaborators. All results were shared and discussed in the round table meetings to (1) interpret and categorize the listed symptoms and (2) establish consensus on most frequent and distinctive symptoms and recommended terminology.

**Search Terms**

This paper focuses on clinical terminology and nomenclature. To improve the impact of the paper, we conducted 13 individual searches, each focusing on a distinct right anterior temporal lobe (RATL)-specific symptom, alongside the expert opinion.

**Table 1: Search terms**

| “frontotemporal dementia” OR “semantic dementia” OR "semantic variant primary progressive aphasia” OR “behavioral variant frontotemporal dementia” OR “temporal variant frontotemporal dementia” OR “frontotemporal lobar degeneration” OR “Pick’s Disease” OR “right temporal” OR “right anterior*” OR “temporal pole” | AND | “prosopagnosia” OR “associative prosopagnosia” OR “face recognition deficit” OR “person knowledge” OR “semantics for people” OR “face agnosia” OR “face blindness” OR “person identification” OR “person-specific” OR “person recognition” OR “face recognition” |
| --- | --- | --- |
|  |  | “emotion recognition” OR “emotion labeling” OR “emotion naming” OR “emotion discrimination” OR “alexithymia” OR “emotion reading” OR “emotional semantic” OR “emotion understanding” OR “emotion perception” OR “empathy” OR “lack of empathy” OR “loss of empathy” OR “emotion” OR “feeling” OR “theory of mind” OR “perspective taking” OR “mentalizing” OR “emotional caricatures” OR “misplaced empathy” OR “affect” OR “affective” |
|  |  | “social cognition” OR “social semantic” OR “paralinguistic” OR “social knowledge” OR “disinhibition” OR “socially inappropriate” OR “social norms” OR “decision making” OR “moral dilemma” OR “social behaviour” OR “social behavior” “impulsivity” “judgment” OR “lack of judgment” OR “criminal behavior” OR “crime” OR “delinquency” OR “misdemeanour” OR “misdemeanor” OR “humor” OR “humour” OR “sarcasm” OR “irony” OR “intertemporal choices” OR “delay discounting” OR “faux pas” |
|  |  | “taste” OR “gustatory agnosia” OR “flavor” OR “flavour” OR “flavor knowledge” OR “taste recognition” OR “taste identification” OR “taste discrimination” OR “food preference” |
|  |  | “sound” OR “vocal agnosia” OR “voice agnosia” OR “phonagnosia” OR “audio” OR “sound knowledge” OR “sound recognition” OR “voice recognition” OR “sound identification” OR “sound discrimination” OR “prosody” OR “tone” OR “auditory agnosia” |
|  |  | “smell” OR “odor” OR “odour” OR “olfactory agnosia” OR “smell knowledge” OR “smell recognition” OR “smell identification” OR “smell discrimination” |
|  |  | “visceral” OR “visceral agnosia” OR “interoceptive agnosia” OR “somatic” OR “psychosomatic” OR “hysteria” OR “pain” OR “temperature” OR “headache” OR “hypochondria” OR “hypochondriacal preoccupation” OR “somatoform” OR “somatoform symptoms” OR “somatoform disorder” OR “hyperalgesia” OR “itching” OR “asomatognosia” OR “alexisomia” OR “bodily sensations” OR “bodily sensation recognition” OR “bodily sensation identification” OR “bodily sensation discrimination” OR “interoceptive” OR “interoception” OR “thermoception” OR “nociception” OR “autonomic” OR “homeostatic” OR “tactile” OR “haptic” OR “somatosensory” OR “self” OR “non-self” OR “sleep” |
|  |  | “landmarks” OR “topographagnosia” OR “landmarks semantic” OR “recognition of monuments” OR “place knowledge” OR “getting lost” |
|  |  | “visual semantic” OR “visual agnosia” OR “object naming” OR “object identification” OR “object recognition” OR “non-verbal semantic” OR “semantic” |
|  |  | “memory” OR “amnesia” OR “episodic memory” OR “semantic memory” OR “autobiographical memory” OR “visual memory” OR “verbal memory” OR “social memory” OR “emotional memory” |
|  |  | “apathy” OR “motivation” OR “affective apathy” OR “social withdrawal” OR “emotional apathy” OR “social interest” OR “social apathy” OR “inertia” OR “anhedonia” OR “reward processing” OR “reward seeking” OR “abulia” OR “aboulia” OR “loss of motivation” |
|  |  | “mental rigidity” OR “mental inflexibility” OR “rigid thinking” OR “hyper-focused interests” OR “preoccupation” OR “compulsion” OR “compulsive” OR “obsession” OR “specific interest” OR “hedonic valuation” OR “reward” OR “hyper-religiosity” OR “mystic” OR “music*” OR “art*” OR “esthetic” OR “color” OR “puzzle” OR “time” OR “schedule” OR “ritualistic” OR “fixated” OR “ritual” OR “religion” OR “colour” OR “game” OR “indulge” OR “punctuality” OR “clock-watching” OR “fixed” OR “routine” OR “gambling” |
|  |  | “depression” OR “mania” OR “mood” OR “altered emotion” OR “affective dysregulation” OR “psychiatric” OR “delusion” OR “hallucination” OR “hyper” OR “impatient” OR “euphoric” OR “cross-modal” OR “synaesthetic” |

**Search strategy**

This systematic review was performed in accordance with the Preferred Reporting Items for Systematic Reviews and Meta-Analyses (PRISMA) statement. Relevant studies were retrieved using keywords in Table 1 from MEDLINE (PubMed) and Embase databases as well as other sources (see PRISMA flow charts), covering the literature until February 2024. The titles and abstracts of the citations were screened by 3 independent authors (H. Ulugut, M. Mountembault, and K. Younes) to determine their relevance for inclusion. Disagreements between authors were resolved through consensus or through the decision of a third author (Y. Pijnenburg). Full-text articles of the relevant citations were then assessed to determine whether the study meets the predefined criteria for RATL case identification (see inclusion criteria and PRISMA flow charts)

**Inclusion criteria/ RATL case identification**

FTD (Neary 1994), FTD (Neary 1998), SD (Neary 1998), FTD (McKhann 2001), bvFTD (Rascovsky 2011), PPA (Gorno Tempini 2011), FTLD (MacKenzie & Neumann 2016)

Right* temporal variant FTD, right predominant* svPPA/SD, right-sided* svPPA/SD, right variant* svPPA/SD, right temporal variant/sided/predominant* bvFTD, RATL atrophy, right temporal lobe atrophy, semantic behavioral variant FTD

**Exclude**

lvPPA, nfvPPA, left predominant* svPPA/SD, left-sided* svPPA/SD, left variant* svPPA/SD, frontal atrophy, frontal predominant FTD, frontal variant FTD, frontal variant/sided/predominant bvFTD, absence of data on predominant atrophy patterns.

*: If the patient is not right-handed and the clinical presentation supports predominance in the opposite hemisphere, the lateralization will be switched

**Atrophy Rating Guidelines:** Right anterior temporal lobe (RATL) predominant impairment is the inclusion criterion of our study. Therefore, all cases have to be examined by at least one structural neuroimaging technic. Since the specificity of PET imaging is relatively low, we considered to include only MRI or CT. We will use the visual assessment method to identify the RATL atrophy. For the mesial temporal atrophy (MTA) scoring please see below (*Scheltens et al., 1992 Atrophy of medial temporal lobes on MRI in "probable" Alzheimer's disease and normal ageing: diagnostic value and neuropsychological correlates*). For the frontotemporal atrophy scoring please see below (*Kipps et al., 2007 Clinical Significance of Lobar Atrophy in Frontotemporal Dementia: Application of an MRI Visual Rating Scale* ). Additionally you will see a protocol for anterior temporal atrophy rating (Credit: Giorgio Fumagalli  and also *Harper et al., 2015 Using visual rating to diagnose dementia: a critical evaluation of MRI atrophy scales*) The most important issue is that the frontal or the left temporal atrophy scores must be lower (at least 1 grade) than the RATL atrophy score. Patients with frontal/left temporal atrophy ≥ 3 (max score=4) (even if they had predominant RATL atrophy) will be excluded to minimize gross effects of general neurodegeneration.

1. **Anterior Temporal Rating Protocol
   1.a. SLICE SELECTION**

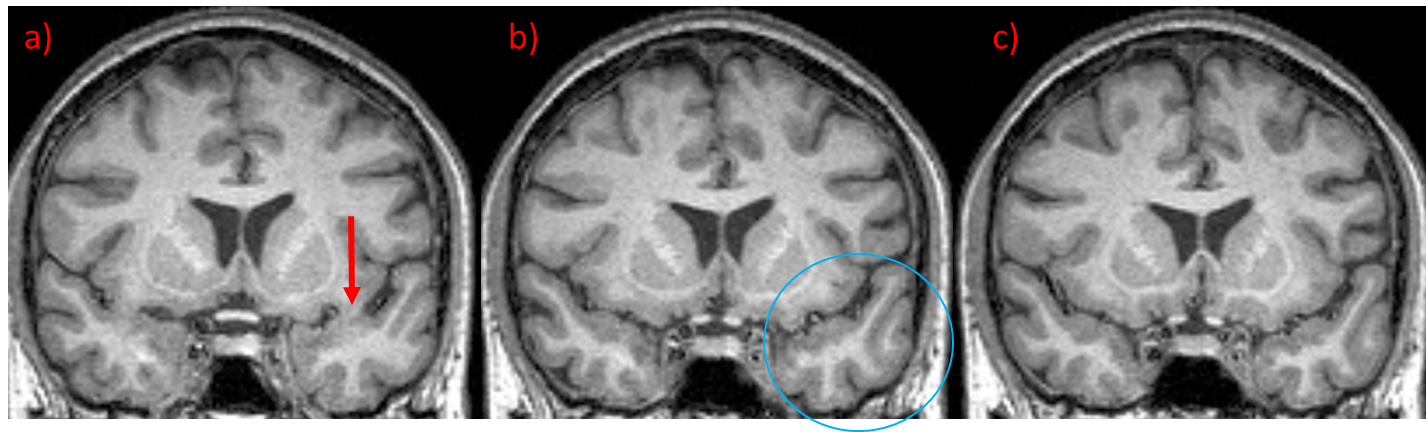
Select the first slice where the connection between the frontal and temporal lobes is NOT visible: rate temporal pole

a) Pre-rating slice: connection between the frontal and temporal lobes is still visible

b) Correct slice: NO visible connection between the frontal and temporal lobes

c) Post-rating slice

**1.b. RATING GUIDE**

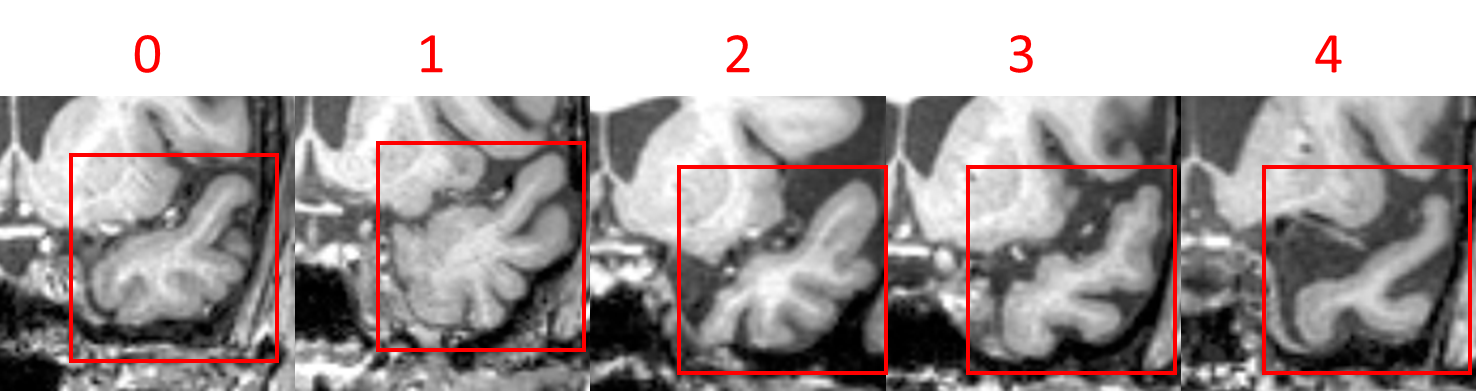

0: Normal appearances (closed sulci)

1: Small sulcal slit of anterior temporal sulci

2: Temporal sulci definitely widened

3: Triangular shape of gyri

4: Severe atrophy of Temporal pole (convex sulci)

2. **Scheltens MTA Scale**

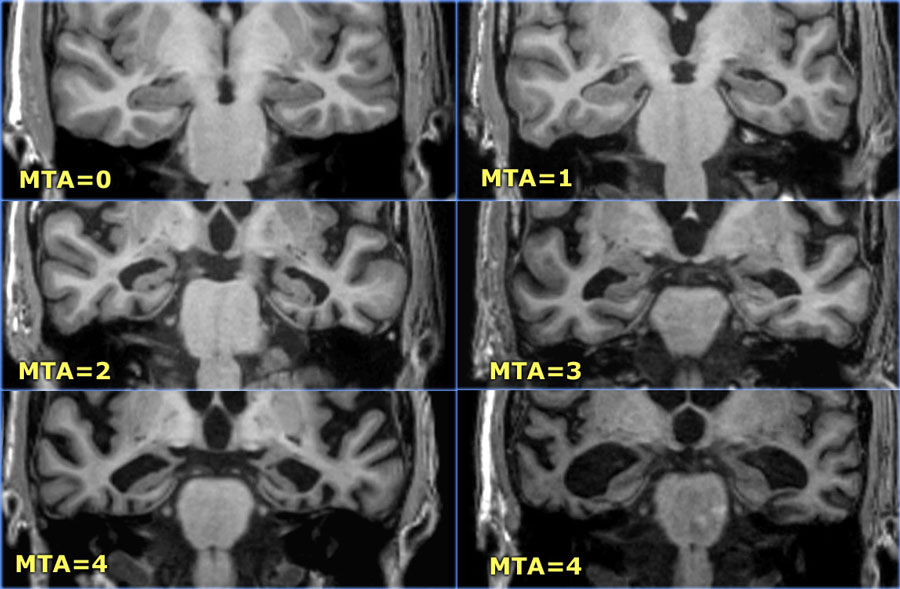

3. **Kipps Frontotemporal Scale**

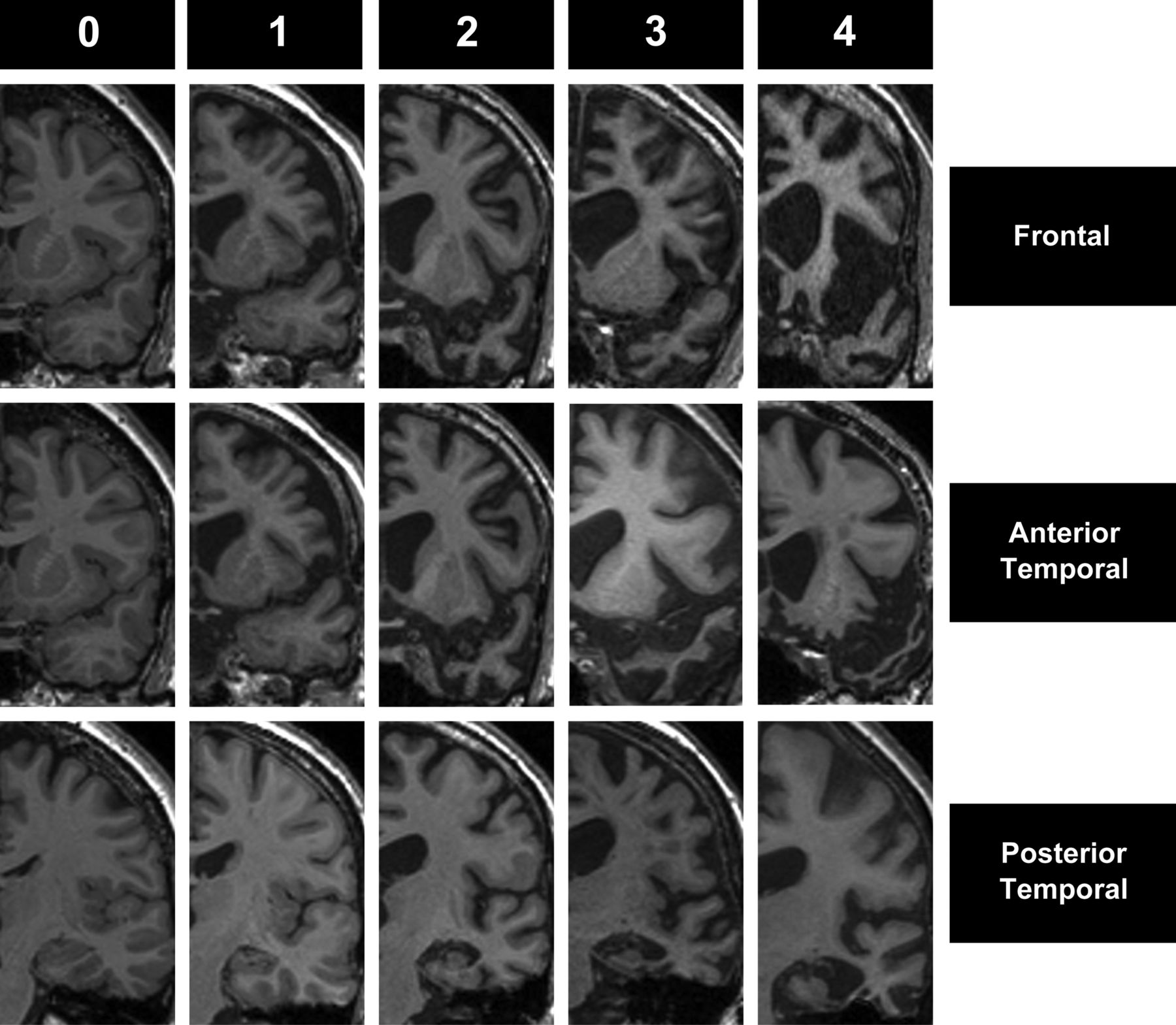

Image is from *Harper et al., 2015 Using visual rating to diagnose dementia: a critical evaluation of MRI atrophy scales*

**Search Results**

| **Category** | **Keywords related disease**  **(Table 1, column 1)** | **Keywords related symptom**  **(Table 1, column 2)** | **Overlap** | **Final Included** |
| --- | --- | --- | --- | --- |
| Person | P: 25,252  E: 50,249 | P: 36,382  E: 7,870 | P: 1,597  E:381 | 23 |
| Emotion | P: 25,252  E: 50,249 | P: 1,073,573  E: 1,416,355 | P: 2,183  E: 4,186 | 28 |
| Social Interaction | P: 25,252  E: 50,249 | P: 85,096  E: 112,956 | P: 119  E: 213 | 6 |
| Taste | P: 25,252  E: 50,249 | P: 70,181  E: 110,265 | P: 65  E: 160 | 10 |
| Sound | P: 25,252  E: 50,249 | P: 222,842  E: 349,233 | P: 482  E: 924 | 13 |
| Smell | P: 25,252  E: 50,249 | P: 46,946  E: 62,991 | P: 77  E: 140 | 2 |
| Landmarks | P: 25,252  E: 50,249 | P: 25,299  E: 30,577 | P: 99  E: 123 | 6 |
| Bodily Sensations | P: 25,252  E: 50,249 | P: 3,614,845  E: 5,139,752 | P: 2,927  E: 7,063 | 15 |
| Visual Stimuli | P: 25,252  E: 50,249 | P: 54,034  E: 68,508 | P:2,112  E: 4,148 | 32 |
| Memory | P: 25,252  E: 50,249 | P: 368,679  E: 541,720 | P: 3,765  E: 8,200 | 58 |
| Apathy | P: 25,252  E: 50,249 | P: 169,814  E: 222,722 | P: 605  E: 1,576 | 30 |
| Rigidity | P: 25,252  E: 50,249 | P: 6,081,465  E: 34,708,791 | P: 4,562  E: 36,708,791 | 24 |
| Psychiatric | P: 25,252  E: 50,249 | P: 894,783  E: 1,429,260 | P: 2,413  E: 6,305 | 21 |

P: Pubmed, E: Embase. Numbers are representing the amount of the articles after each search. Final included: The number of the articles after the exclusion of duplications, irrelevant articles, reviews, metanalyses, commentaries, full text unavailable abstracts, and studies not including RATL cases based on patient selection criteria listed above.
